# Genome-wide cross-trait analysis of vascular dementia and Alzheimer’s disease highlights novel loci and lung-brain axis

**DOI:** 10.64898/2026.02.27.26345967

**Authors:** Shan Gao, Shiyang Wu, Fengzhen Liu, Ping Zhu, Yijie He, Shuyuan Hu, Ruibai Wang, Jin Yang, Lu Zhao, Xuman Liu, Zhifa Han, Tao Wang, Yan Zhang, Kun Wang, Yan Chen, Keshen Li, Guiyou Liu

## Abstract

Until now, most genetic risk for vascular dementia (VD) remains unknown. Here, we firstly performed the largest cross-ancestry genome-wide association study meta-analysis comprising 5,886 VD and 1,027,883 controls of European, East Asian, South Asian, African, and Admixed American ancestry. We identified 37 genome-wide significant loci including *CLU* and *APOE* tagged by common variants and 35 loci tagged by rare variants, and demonstrated enrichment of VD heritability in lung and genetic association between VD and lung function traits. We further conducted a cross-trait of VD and Alzheimer’s disease, and identified 13 genome-wide significant loci including *CR1*, *BIN1*, *GRM7*, *HLA-DRA*, *TREM2*, *CLU*, *ECHDC3*, *AGBL2*, *MS4A4E*, *PICALM*, *SLC24A4*, *ABCA7*, and *APOE.* A multi-omics integrative analysis identified 619 genes. 241 genes were significantly differentially expressed in VD cells and 21 exhibited strong evidence of interaction with FDA-approved drugs. Collectively, our findings provide valuable insights into the potential underlying mechanisms of VD.

## Introduction

Vascular dementia (VD), caused by reduced blood flow to the brain, is the second most common form of dementia after Alzheimer’s disease (AD) and accounts for at least 20% of dementia cases ^1,2^. Genetic factors play important roles in the etiology of both AD and VD ^1,2^. Until now, large-scale genome-wide association study (GWAS) datasets have identified the common AD genetic variants and risk loci especially *APOE* ^3^. Unlike AD, there are only few VD GWAS and limited sample sizes including 67 cases and 5,700 controls ^4^, 84 cases and 200 controls ^5^, 373 cases and 3,289 controls ^6^, 89 cases and 3,016 controls ^6^. The Mega Vascular Cognitive Impairment and Dementia (MEGAVCID) Consortium performed a large-scale GWAS including 3,892 VD and 466,606 controls of European descent, and only identified one genetic variant rs429358 near the *APOE* reaching the genome-wide significance with *P=*2.90E-196 ^7^. Until now, the majority of genetic risk of VD remains unknown.

It is known that GWAS was designed to broadly capture the common genetic variants with a minor allele frequency (MAF) greater than 1% ^8,9^. Rare variants were not tagged by common genetic variants from genotyping arrays and imputation ^8,9^. It was largely unknown about the contribution of rare variants (MAF<1%) to human traits and diseases ^8,9^. Until recently, publicly available biobanks using whole-genome sequence offered an unprecedented opportunity to assess the effects of both common and rare genetic variants on human traits and diseases, and highlighted large effects and significant contribution of rare variants ^10–14^.

Here, we hypothesized that VD GWAS using rare variants may contribute to (1) increase the number of novel genetic variants and susceptibility loci, (2) identify the rare variants of large effects, and (3) increase the proportion of heritability. We collected four publicly available biobanks, and conducted the largest VD cross-ancestry GWAS meta-analysis to date in 5,886 patients diagnosed with VD and 1,027,883 control individuals from five ancestral populations: European, East Asian, South Asian, African, and Admixed American using genetic variants with MAF>0.01%. We further systematically characterized the genetic architecture of VD using multiple multi-omics integration approaches including gene mapping, gene-based association test, polygenic priority score (PoPS) analysis, gene set enrichment analysis, tissue enrichment analysis, transcriptome-wide association study (TWAS), colocalization analysis, summary-data-based Mendelian randomization (SMR), multi-trait analysis of GWAS (MTAG), case-control gene expression analysis, drug-gene interaction analysis, and genetic correlation analysis. An overview of the workflow is provided in Fig 1.

**Fig 1.**
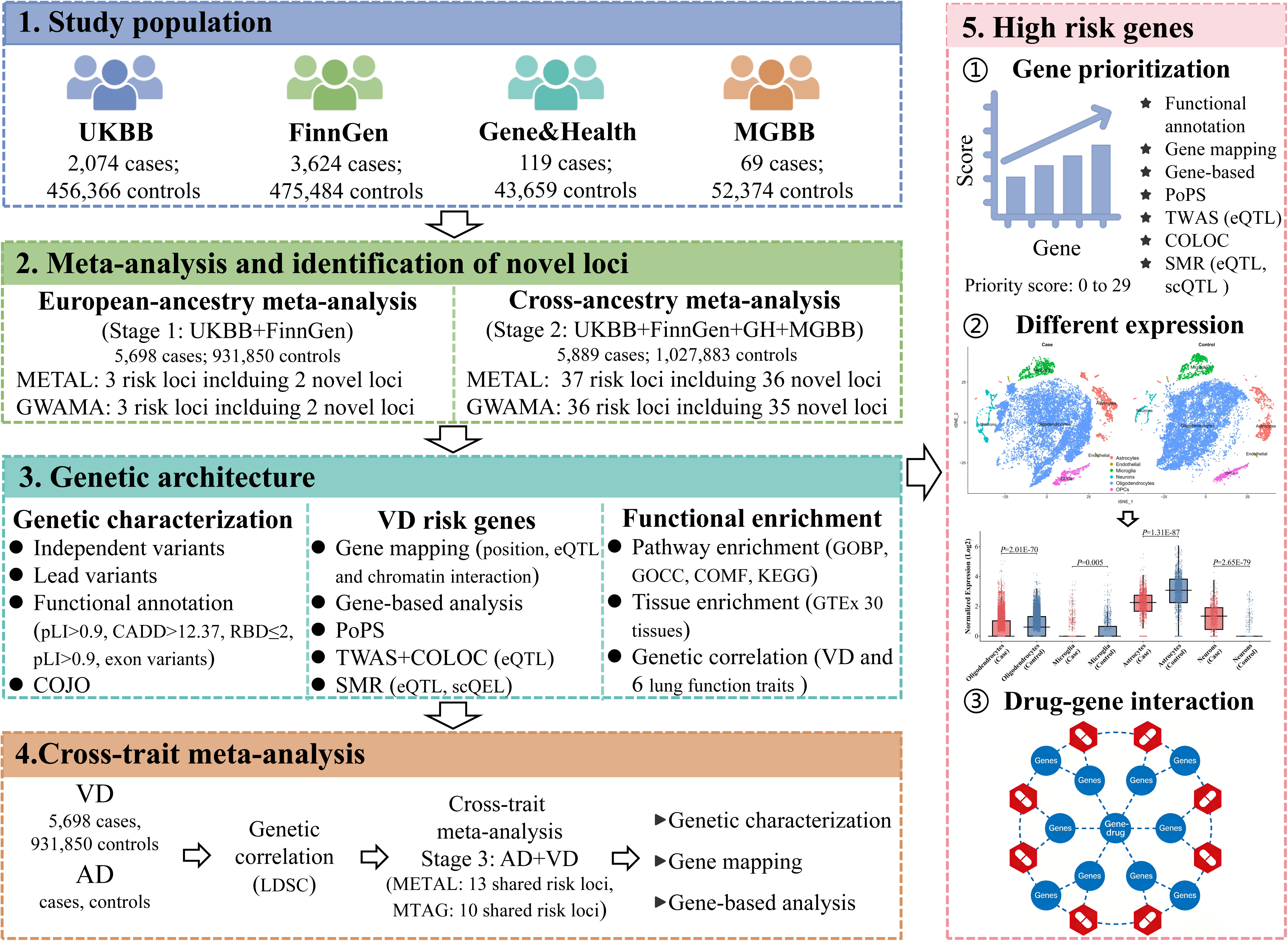
Study overview. **Part 1**. Data sources and study population. We analyzed genome-wide association study (GWAS) summary statistics from four publicly available biobanks/cohorts comprising five ancestral populations: European (EUR), East Asian (EAS), South Asian (SAS), African (AFR), and Admixed American ancestry (AMR). **Part 2**. Two-stage genetic discovery. Stage 1, European meta-analysis of UKBB and FinnGen (5,698 cases and 931,850 controls) identified 3 genome-wide significant loci (2 novel). Stage 2, Cross-ancestry meta-analysis of all four cohorts (5,889 cases and 1,027,883 controls) identified 37 significant loci. Analyses were performed with METAL and GWAMA. **Part 3**. Characterization of genetic architecture. We performed functional annotation of risk variants and prioritized candidate genes through gene mapping, gene-based association testing, polygenic priority score (PoPS) analysis, transcriptome-wide association study (TWAS) with colocalization (COLOC), and summary-data-based Mendelian randomization (SMR). Pathway, tissue enrichment, and genetic correlation with lung function traits were also assessed. **Part 4**. Cross-trait genetic analysis. A meta-analysis of VD and Alzheimer’s disease (AD) GWAS data using METAL and MTAG revealed shared genetic risk loci and shared risk genes. **Part 5**. Functional validation and drug target discovery. Genes were prioritized by integrating 29 lines of evidence. Their differential expression was validated in single-cell RNA sequencing data from VD patients, leading to the identification of potential druggable targets.

## Result

### European-specific GWAS meta-analysis (stage 1)

We selected two independent VD GWAS datasets including 5,698 patients diagnosed with VD and 931,850 control individuals of European descent from UKBB (2,074 VD and 456,366 controls) ^15^ and FinnGen R12 (3,624 VD and 475,484 controls) ^16^ (Methods, Supplementary Table 1). We conducted a large-scale GWAS meta-analysis of both datasets using a fixed-effects inverse-variance weighted (IVW) method implemented in METAL ^17^. Using linkage disequilibrium (LD) score regression (LDSC) ^18^, we estimated the single nucleotide polymorphism (SNP) based heritability on liability scale to be □^2^=6.57% and s.e.=0.0132, assuming a VD population prevalence of 1.16% ^19^. The genomic inflation factor (λ_GC_) was 1.0754 and the LDSC intercept was 1.0241 (s.e.=0.0078), which indicated little evidence of genetic inflation (Supplementary Fig. 1A).

Utilizing Functional Mapping and Annotation (FUMA) ^20^, we identified three independent genome-wide significant loci including *HTR4*, *CLU*, and *APOE*, which are tagged by rs564080066, rs7982 and rs429358, respectively (Fig. 2A, Table 1, Supplementary Fig. 2). A sensitivity GWAS meta-analysis using GWAMA (Genome-Wide Association Meta-Analysis) further confirmed all three loci (*P*<5.00E-08) ^21^ (Fig. 2A). rs564080066 is novel and rare variant with an effect allele frequency of 0.0023 in European population (Table 1). rs7982 is a proxy of rs11136000 (*r*²=0.98 and D’=0.99), which was a known genome-wide significant variant associated with AD ^22^. rs429358 is a well-known genome-wide significant variant associated with multiple dementias including VD ^7^, AD ^22^, frontotemporal dementia (FTD) ^23^, and Lewy body dementia (LBD) ^24^. Using FUMA ^20^, we identified 48 independent genetic variants (*r*^2^<0.6, Supplementary Table 2) and 21 independent lead variants (*r*^2^<0.1, Supplementary Table 3) around these three loci. A stepwise conditional analysis using GCTA conditional and joint (COJO) analysis and linkage disequilibrium (LD) information from UKBB individuals ^25^ further confirmed these three loci (Supplementary Table 4). Using positional mapping, eQTLs mapping, and chromatin interaction mapping (Supplementary Table 5, Supplementary Data 1), we identified 120 VD risk genes (Fig. 2C, Supplementary Table 6).

**Fig 2.**
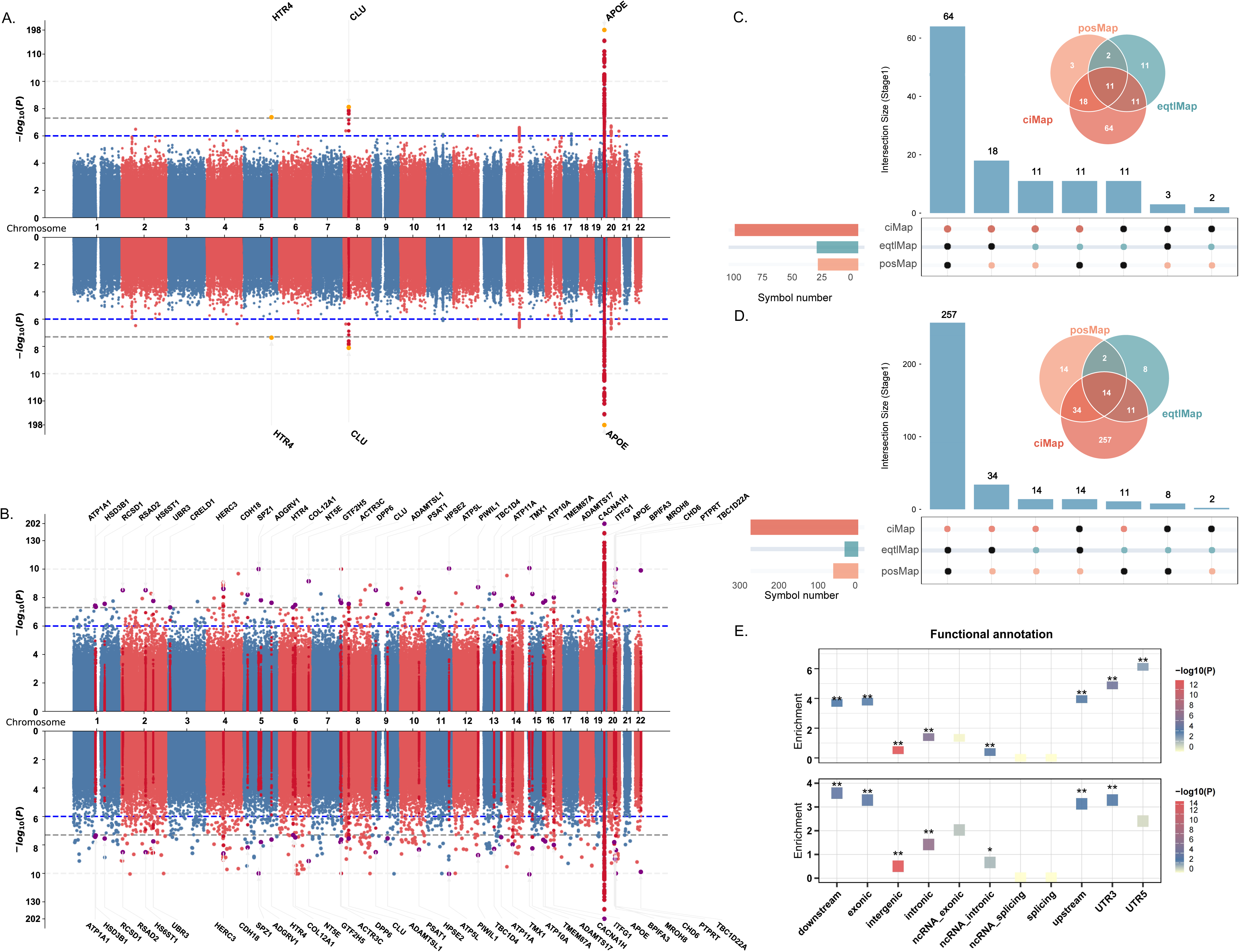
Identification and functional annotation of vascular dementia risk loci through GWAS meta-analyses. **A.** Manhattan plots from the Stage 1 (European-specific) GWAS meta-analysis performed with METAL (upwards) and GWAMA (downwards). **B.** Manhattan plots from the Stage 2 (cross-ancestry) GWAS meta-analysis performed with METAL (upwards) and GWAMA (downwards). In A and B, the horizontal axis shows the chromosomal position (chromosomes 1-22) and the vertical axis shows the significance (-log_10_ *P* value) of tested markers. *P* values are two-sided and based on an inverse variance weighted (IVW) fixed effects meta-analysis. Each dot represents a genetic variant. The threshold for genome-wide significance (*P*<5.00E-08) is indicated by a grey dotted line, and genome-wide significance loci are shown in red. *P* values are two-sided and derived from an inverse-variance-weighted fixed-effects meta-analysis. **C.** An UpSet plot and a Venn diagram illustrating the overlap of candidate risk genes identified through three independent mapping strategies (positional mapping, expression quantitative trait locus mapping, and chromatin interaction mapping) within the genome-wide significant loci from Stage 1. **D.** An UpSet plot and a Venn diagram showing the overlap of candidate risk genes identified by the same three mapping strategies within the genome-wide significant loci from Stage 2. **E.** Scatter plot summarizing the results of functional enrichment analyses for variants within the genome-wide significant loci from Stage 1 (upwards) and Stage 2 (downwards). Asterisks denote statistical significance: * *P*<0.05; ** *P*<0.05/11.

**Table 1.**
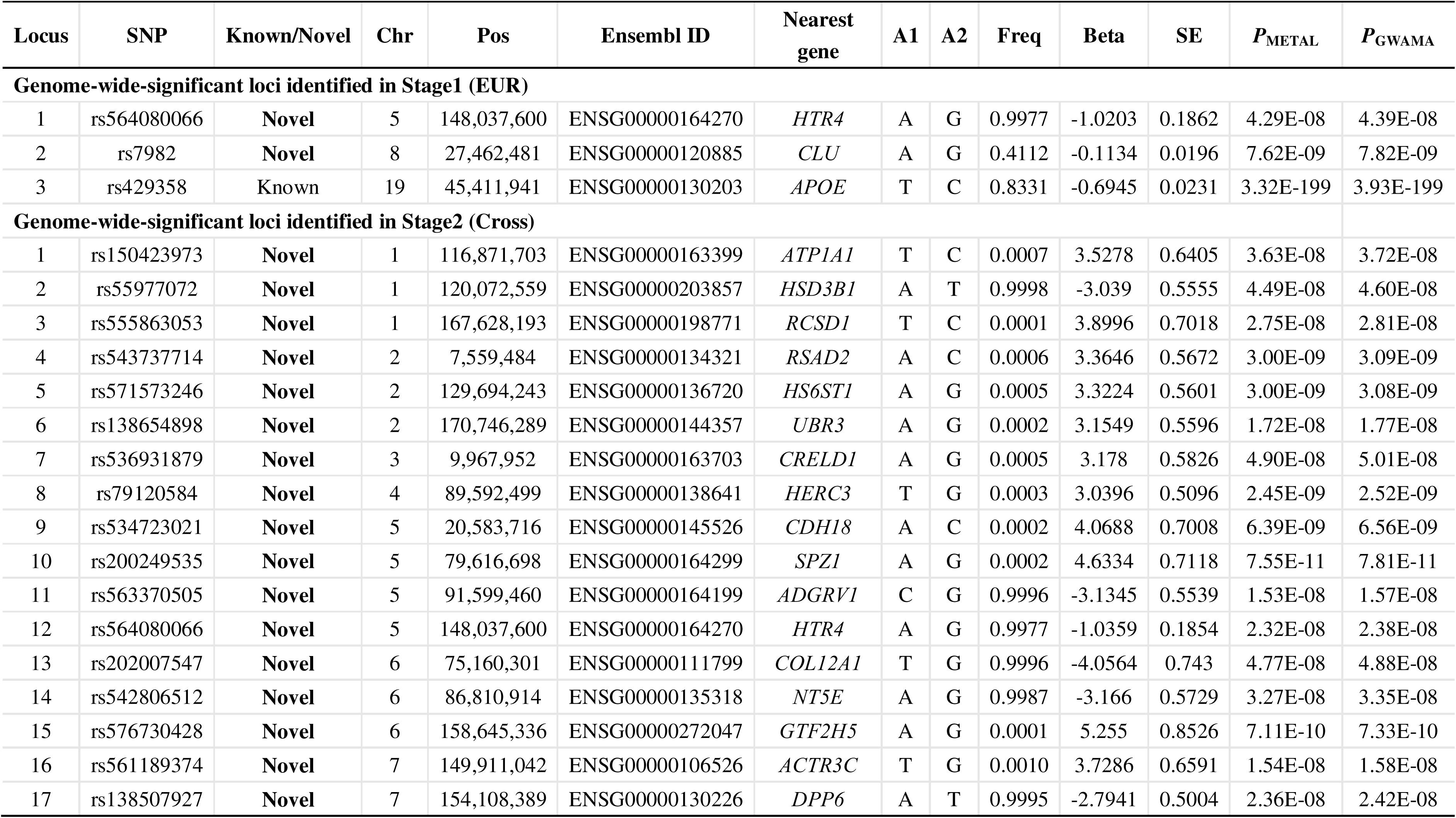
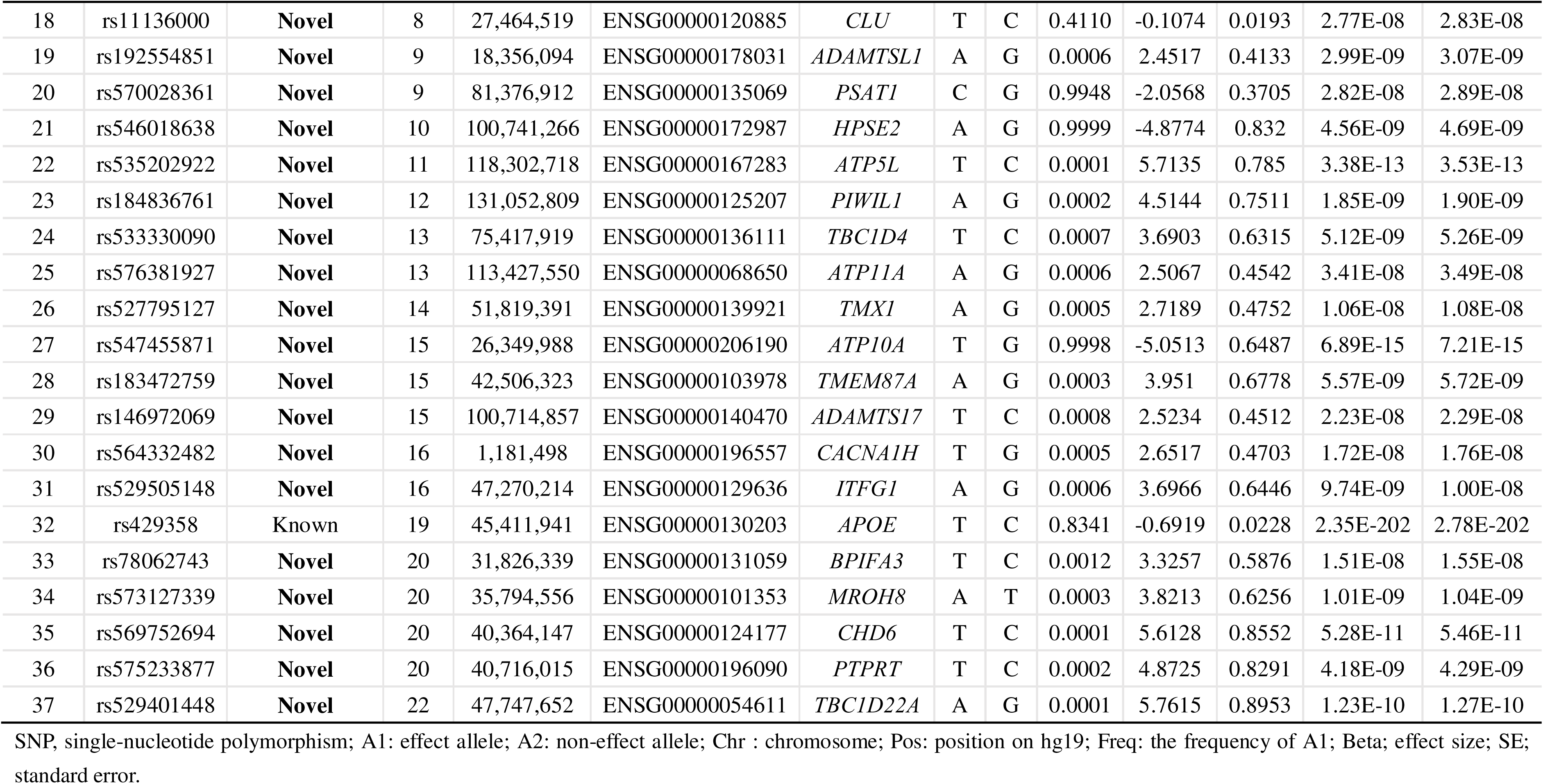
Genome-wide signifiant loci from vascular dementia GWAS meta-analysis in stage 1 and stage 2.

### Cross-ancestry GWAS meta-analysis (stage 2)

We conducted a cross-ancestry VD GWAS meta-analysis using the fixed-effects IVW meta-analysis method including 5,886 patients diagnosed with VD and 1,027,883 control individuals using four independent GWAS datasets from UKBB (European, 2,074 VD and 456,366 controls) ^15^, FinnGen R12 (European, 3,624 VD and 475,484 controls) ^16^, Genes & Health (GH, South Asian, 119 VD and 43,659 controls) ^26^, and Mass General Brigham Biobank (MGBB, European, South Asian, African, and Admixed American, 69 VD and 52,374 controls) (Methods, Supplementary Table 1)^27^. The genomic inflation factor λ_GC_=1.0741 and LDSC intercept of 1.0204 (s.e.= 0.0074) showed little evidence of genetic inflation (Supplementary Fig. 1B). The SNP-based heritability on liability scale was *h*^2^=6.47% (s.e.=0.0112) assuming the VD population prevalence of 1.16% ^19^. We revealed 37 independent genome-wide significant loci by confirming *HTR4* (rs564080066), *CLU* (rs11136000), and *APOE* (rs429358) from European-specific GWAS meta-analysis, and highlighting 34 novel loci all tagged by rare variants (Fig. 2B, Table 1, Supplementary Fig. 3). These 37 GWAS loci explained 53% of VD variance including 13.25% from loci tagged by common variants and 39.19% from loci tagged by rare variants. A sensitivity GWAS meta-analysis using GWAMA shown in Fig. 2B further confirmed 36 loci (excluding *CRELD1*) (*P*<5.00E-08) ^21^. Using FUMA, we identified 111 independent genetic variants (*r*^2^<0.6, Supplementary Table 7) and 66 independent lead variants (*r*^2^<0.1, Supplementary Table 8) around these three loci (*HTR4*, *CLU*, *APOE*). GCTA COJO analysis further confirmed these loci (Supplementary Table 9, Supplementary Data 2). Using positional mapping, eQTLs mapping, and chromatin interaction mapping, we identified 340 VD risk genes (Fig. 2D, Supplementary Table 10).

### Functional annotation

In stage 1, we annotated 388 SNPs in LD with the 48 independent significant lead variants using ANNOVAR ^28^, and found predominant enrichment in the intronic, upstream, downstream, exonic, UTR3 and UTR5 (Supplementary Tables 11-12 and Fig. 2E). Among these 388 SNPs, we identified 286 SNPs that were in LD (*r*^2^>0.6) with 48 independent significant lead variants, extracted from the 1000 genomes European reference panel (Supplementary Table 13) ^29^. rs564080066 is an intronic variant with a Combined Annotation Dependent Depletion (CADD) score of 3.382, indicating a moderate potential of regulatory impact (Supplementary Table 13) ^30^. rs7982 is an exonic variant with CADD score of 1.763, indicating a moderate potential of regulatory impact (Supplementary Table 13). rs429358 is an exonic variant with a high CADD score of 12.64, suggesting a potential of deleterious effects (Supplementary Table 13). In stage 2, we annotated 381 SNPs in LD with the 89 independent lead variants using ANNOVAR ^28^, and revealed predominant enrichment in intronic, downstream, exonic, upstream, ncRNA_exonic and UTR3 genomic categories (Fig. 2E, Supplementary Tables 14-15). Among these 381 SNPs, 278 SNPs were in LD (*r*^2^>0.6) with 89 independent significant lead variants, extracted from the 1000 genomes ALL reference panel (Supplementary Table 16) ^29^.

### Gene-based association test, gene set and tissue enrichment analyses

It is noted all subsequent analyses were performed using the stage 1 GWAS summary statistics. We conducted a gene-based association test, gene set enrichment analysis, and tissue enrichment analysis of stage 1 VD GWAS meta-analysis summary data using MAGMA ^31^. Gene-based association test identified 18 statistically significant genes with the Benjamini-Hochberg FDR-corrected *P*<0.05 (Fig. 3A, Supplementary Table 17). 15 genes are located within the *APOE* region including *APOC1*, *APOE*, *APOC4*, *NECTIN2*, *TOMM40*, *RELB*, *CEACAM16*, *PVR*, *CLPTM1*, *IGSF23*, *CEACAM19*, *EXOC3L2*, *KLC3*, *ZNF226*, *ZNF227*. 3 genes are located outside the *APOE* region including *CLU*, *CALCRL*, and *WNK1* (Supplementary Table 17). In brief, gene-based association test not only verified two GWAS loci *APOE* and *CLU*, but also highlighted two additional novel genes *CALCRL* and *WNK1* (Supplementary Table 17).

**Fig 3.**
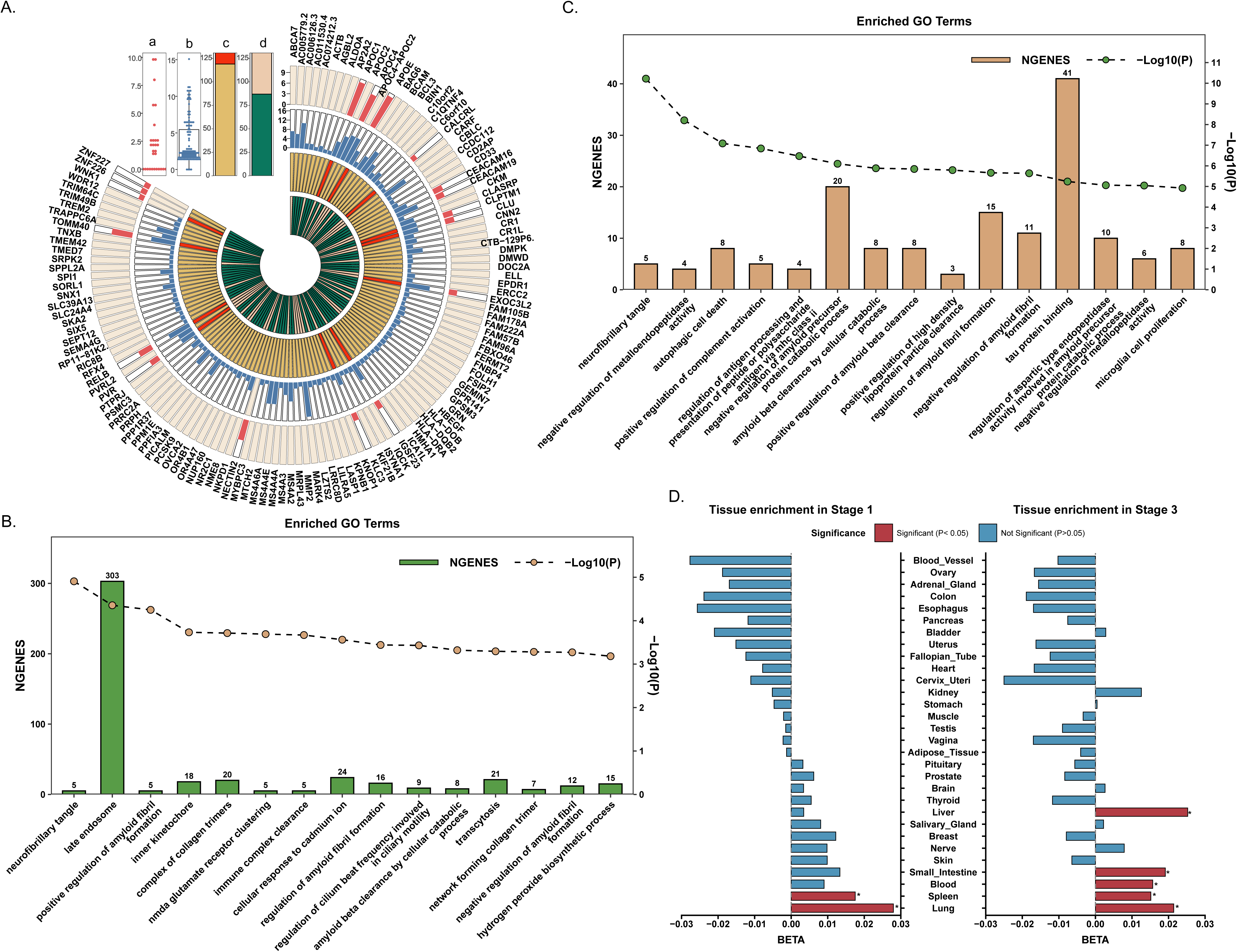
Gene-based association test, polygenic priority score analysis, gene-set enrichment analysis, and tissue enrichment analysis. **A.** Multi-track circular plot synthesizes the result of gene-based association and polygenic priority score (PoPS) analyses from Stage 1 and Stage 3. The concentric tracks, from outermost to innermost, sequentially display: the union of genes mapped to significant loci (FDR<0.05) from either stage; genes identified by gene-based association analysis in Stage 1 (the y-axis represents -log_10_ adjusted *P*-value), genes identified by gene-based association analysis in Stage 3 (the y-axis represents -log_10_ adjusted *P*-value), the top 10% of PoPS in Stage 1 (genes within this top 10% PoPS rank are filled in red), and top 10% of PoPS in Stage 3 (genes within this top 10% PoPS rank are filled in prink). **B.** Dual-axis bar chart presents the significant results (FDR-adjusted *P*<0.05) of gene set enrichment analysis performed on Stage 1 using MAGMA. **C.** Dual-axis bar chart presents the significant results (FDR-adjusted *P*<0.05) of gene set enrichment analysis performed on Stage 3 using MAGMA. In B and C,the x-axis lists the names of the significant Gene Ontology (GO) terms. The left y-axis indicates the number of enriched genes (NGENES) within each term, represented by the bars. The right y-axis indicates the statistical significance level (-log_10_ *P*-value) of the enrichment for each term, represented by the line plot with points. **D.** Butterfly bar chart presents the results of tissue-specific enrichment analysis for Stage 1 and Stage 3 VD risk loci, performed using MAGMA based on the GTEx dataset. The y-axis lists the names of 30 tissues from GTEx. The horizontal position (x-axis) of each bar represents the enrichment beta value. Bars extending to the left depict enrichment results for Stage 1, while bars extending to the right depict results for Stage 3. For each stage, tissues with statistically significant enrichment (FDR-adjusted *P*<0.05) are filled in red, and non-significant tissues are filled in blue.

Gene set enrichment analysis identified two statistically significant gene ontology (GO) cellular components with the Benjamini-Hochberg FDR-corrected *P*<0.05 including neurofibrillary tangle (GO:0097418, *P*=1.24E-05, and FDR=1.29E-02), and late endosome (GO:0005770, *P*=4.42E-05, and FDR=2.30E-02) (Fig. 3B, Supplementary Table 18). Meanwhile, 5 GO biological processes including positive regulation of amyloid fibril formation (GO:1905908, *P*=5.63E-05, and FDR=1.96E-01), NMDA glutamate receptor clustering (GO:0097114, *P*=2.04E-04, and FDR=3.19E-01), regulation of amyloid fibril formation (GO:1905906, *P*=3.62E-0504, and FDR=3.88E-01), amyloid-beta clearance by cellular catabolic process (GO:0150094, *P*=4.78E-04, and FDR=4.00E-01), negative regulation of amyloid fibril formation (GO:1905907, *P*=5.35E-04, and FDR=4.00E-01) showed suggestive association with VD (Supplementary Table 18). Tissue enrichment analysis showed evidence of enrichment in GTEx v8 lung (*P*=4.60E-03) and spleen (*P*=1.46E-02), but not in brain tissues or blood (*P*=7.66E-02) (Fig. 3D, Supplementary Table 19).

### Genetic association between VD and lung function traits

Tissue enrichment analysis showed that the VD heritability was mainly enriched in GTEx v8 lung (*P*=4.60E-03) (Supplementary Table 20). Here, we further investigated the genetic correlation between VD (stage 1) and 6 lung function traits including forced expiratory volume in 1-second (FEV1), forced vital capacity (FVC), FEV1/FVC, peak expiratory flow (PEF), asthma and COPD using LDSC (Supplementary Table 20) ^18^. We identified that VD showed statistically significant negative genetic correlation with FEV1 (*rg*=-0.1695, *P*=5.20E-03) and FVC (*rg*=-0.1678, *P*=3.10E-03) using a Bonferroni-corrected statistical significance threshold of 0.05/6. VD was suggestively associated with PEF (*rg*=-0.1528, *P*=1.26E-02), and asthma (*rg*=0.1965, *P*=4.81E-02) (Supplementary Table 21). These findings showed that impaired lung function was associated with a higher risk of VD.

### Polygenic Priority Score

Polygenic Priority Score (PoPS) is a new method that learns trait-relevant gene features to pinpoint the most likely causal genes at GWAS loci ^32^. We computed the polygenic priority score by combining the stage 1 VD GWAS meta-analysis summary statistics with biological pathways, gene expression and protein–protein interaction data ^32^. Among 18 statistically significant genes from gene-based association test, *APOE*, *APOC1*, *CLU*, *RELB*, *WNK1*, *PVR*, *IGSF23*, *CALCRL*, *EXOC3L2*, *TOMM40*, *CLPTM1*, and *KLC3* were ranked the top 10% of the polygenic priority scores (range: 0.37–4.22), underscoring their likely functional relevance and potential roles in the pathogenesis of VD. *APOE*, *CLU*, *WNK1*, and *CALCRL* were ranked with the highest, 4^th^, 42^nd^ and 125^th^ polygenic priority scores, respectively (Fig. 3A, Supplementary Table 22).

### Transcriptome-wide association study and colocalization analysis

We performed a TWAS to identify the genes whose expression levels are implicated in the pathogenesis of VD ^33^. Here, we integrated the stage 1 VD GWAS meta-analysis dataset with the gene expression data from Genotype-Tissue Expression (GTEx) ^34^, Young Finns Study (YFS) ^35^, and brain eQTLs datasets from CommonMind Consortium (CMC) ^36^, respectively. We identified 25 transcriptome-wide significant VD genes with the Benjamini-Hochberg FDR-corrected *P*<0.05 including 17 genes around the *APOE* region and 8 genes outside the *APOE* region including *CLU* (novel GWAS locus and novel gene from gene-based association test), *WNK1* (novel gene from gene-based association test), *SLC17A4*, *PARD3*, *AFF1*, *FBXW8*, *WDR27*, and *WWOX* (Fig. 4A-B, Supplementary Table 23). These genes exhibited multiple lines of evidence with VD including *CLU* (2 tissues), *WNK1* (2 tissues), *SLC17A4* (4 tissues), *PARD3* (aorta artery), *AFF1* (6 tissues), *FBXW8* (24 tissues), *WDR27* (41 tissues), and *WWOX* (24 tissues) (Fig. 4A-B, Supplementary Table 23). *CALCRL*, the novel gene from gene-based association test, also showed evidence of association with VD in 9 tissues including aorta artery, coronary artery, tibial artery, cerebellum, tibial nerve, and whole blood. Bayesian colocalization analysis highlighted that *CLU*, *WWOX*, and *NKPD1* (within the *APOE* region) showed evidence of colocalization with PPH_4_>0.70 (Fig. 4A-B, Supplementary Table 24).

**Fig 4.**
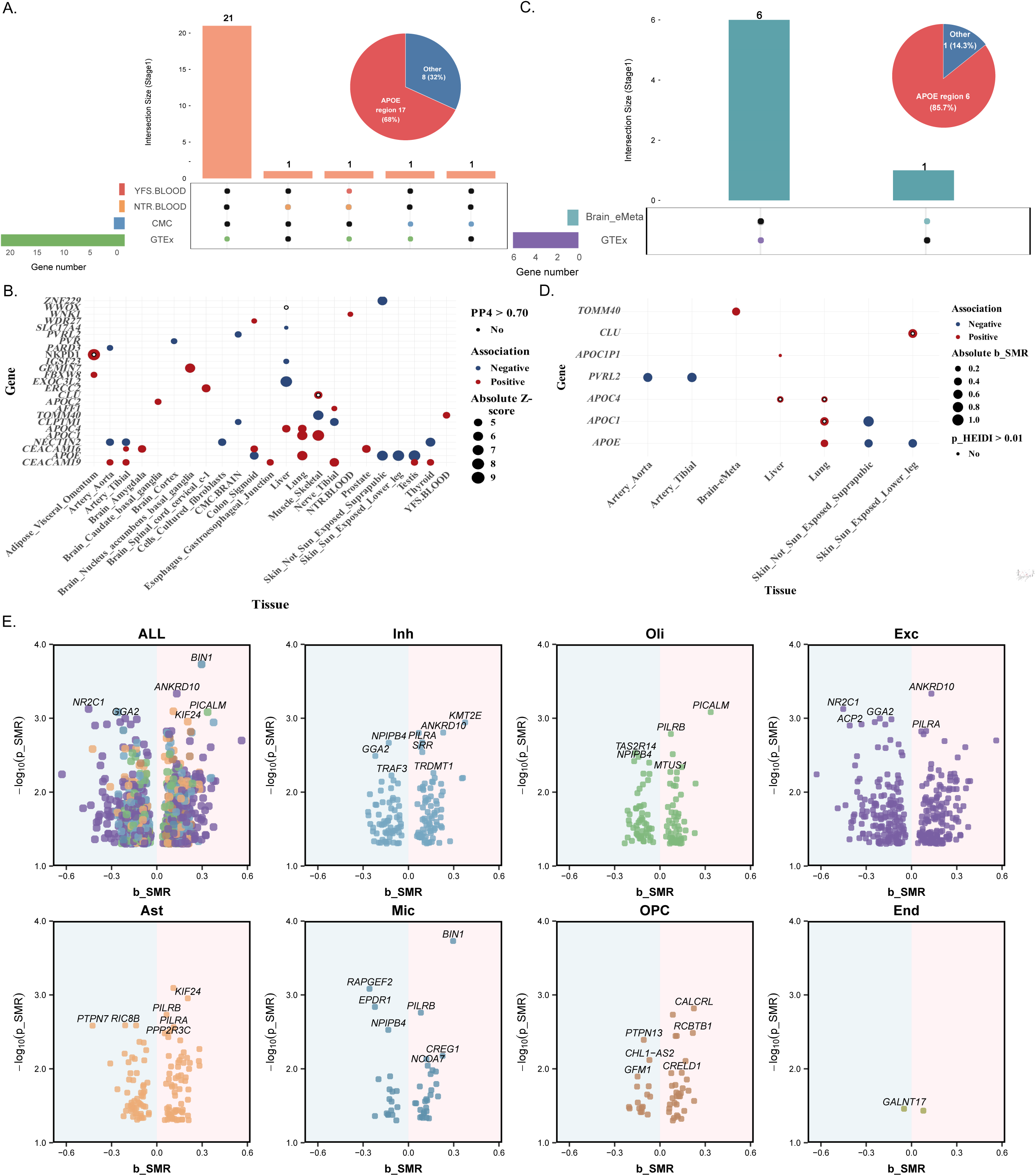
Integrative analysis of GWAS and multi-omics xQTLs in Stage 1. **A.** Integrative analysis of VD Stage 1 and eQTLs datasets in relevant tissues using TWAS. The size of each dot represents the absolute z-score for each gene, and the dots are colored according to the direction of the effect. White dots indicate the associations with PPH4>0.70 in coloc analysis. **B.** Upset plot of genes identified via TWAS using expression weights data from 51 eQTLs datasets. **C.** Integrative analysis of VD Stage 1 and eQTLs datasets in relevant tissues using SMR. The size of each dot represents the beta for each gene, and the dots are colored according to the direction of the effect. White dots indicate the associations with *P*_HEIDI_>0.01. **D.** Upset plot of genes identified via SMR using expression weights data from eQTLs datasets. **E.** Grouped volcano plot validates genes identified via SMR using expression weights data from sc-qtl dataset. The seven panels correspond to seven major cell types. In each panel, the x-axis represents effect (b_SMR) and the y-axis represents the significance (-log_10_ *p*_SMR). Each point represents a single gene within the specified cell type, visually confirming its dysregulation in the VD.

### Summary-data-based Mendelian randomization

We conducted a SMR ^37^ to identify genes putatively causally associated with VD by integrating the stage 1 VD GWAS meta-analysis summary data with multiple eQTLs datasets from GTEx (v8 54 tissues) ^34^, eQTLGen (whole blood) ^38^, BrainMeta v2 (brain) ^39^, and brain single-nucleus eQTLs datasets ^40^. Using bulk tissue eQTLs datasets, we revealed 3 statistically significant genes including *APOC4*, *APOC1*, and *CLU* with the Benjamini-Hochberg FDR-corrected *P*<0.05 and HEIDI (heterogeneity in dependent instruments) test *P*>0.01 (Fig. 4C-D, Supplementary Table 25). Collectively, both SMR and TWAS provide consistent findings about the involvement of *APOE* and *CLU* in VD. Using single-nucleus eQTLs datasets, SMR highlighted *PICALM* as the only statistically significant gene with Benjamini-Hochberg FDR-corrected *P*<0.05 in microglia. In brief, genetically increased *PICALM* expression in microglia was associated with significantly decreased risk of VD with beta=-0.18, *P*_SMR_=4.52E-05, FDR=0.03, and *P*_HEIDI_=0.87 (Fig. 4E, Supplementary Table 26). Meanwhile, *PICALM* was suggestively associated with the risk of VD in oligodendrocyte (beta=0.34, *P*_SMR_=8.25E-04, and *P*_HEIDI_=0.14) and excitatory neuron (beta=-0.63, *P*_SMR_=5.74E-03, and *P*_HEIDI_=0.52) (Fig. 4E, Supplementary Table 26).

### Cross-trait meta-analysis of VD and AD (stage 3)

We first estimated the genetic correlation between VD (stage 1) and AD (European) ^22^ using LDSC ^18^, and found a statistically significant positive genetic correlation with *r*g=0.4469 and *P*=3.75E-14, which suggested that it was appropriate and reliable to combine both GWAS datasets. We further conducted a cross-trait meta-analysis of VD (stage 1) and AD GWAS datasets using the fixed-effects IVW method implemented in METAL ^17^, and identified 13 independent genome-wide significant loci including *CR1*, *BIN1*, *GRM7*, *HLA-DRA*, *TREM2*, *CLU*, *ECHDC3*, *AGBL2*, *MS4A4E*, *PICALM*, *SLC24A4*, *ABCA7*, and *APOE* (Fig. 5A-C, Supplementary Table 27). Subsequently, we performed a sensitivity cross-trait meta-analysis of AD and VD GWAS datasets using multi-trait analysis of GWAS (MTAG) ^41^. MTAG identified 10 genome-wide significant loci for AD and 11 loci for VD, of which 10 loci were shared between the two traits. Interestingly, all these 10 shared loci were known loci identified using METAL, further confirming the robustness and pleiotropic nature of these genetic signals (Fig. 5D, Supplementary Table 28). Using FUMA ^20^, we further delineated 213 independent genetic variants (*r*^2^< 0.6; Supplementary Table 29) and 72 independent lead variants (*r*^2^< 0.1; Supplementary Table 30), and annotated 2,213 SNPs in LD with the 213 independent significant lead variants across these loci. Through positional mapping, eQTL mapping, and chromatin□interaction mapping, we ultimately identified 353 candidate risk genes (Supplementary Tables 31-33).

**Fig 5.**
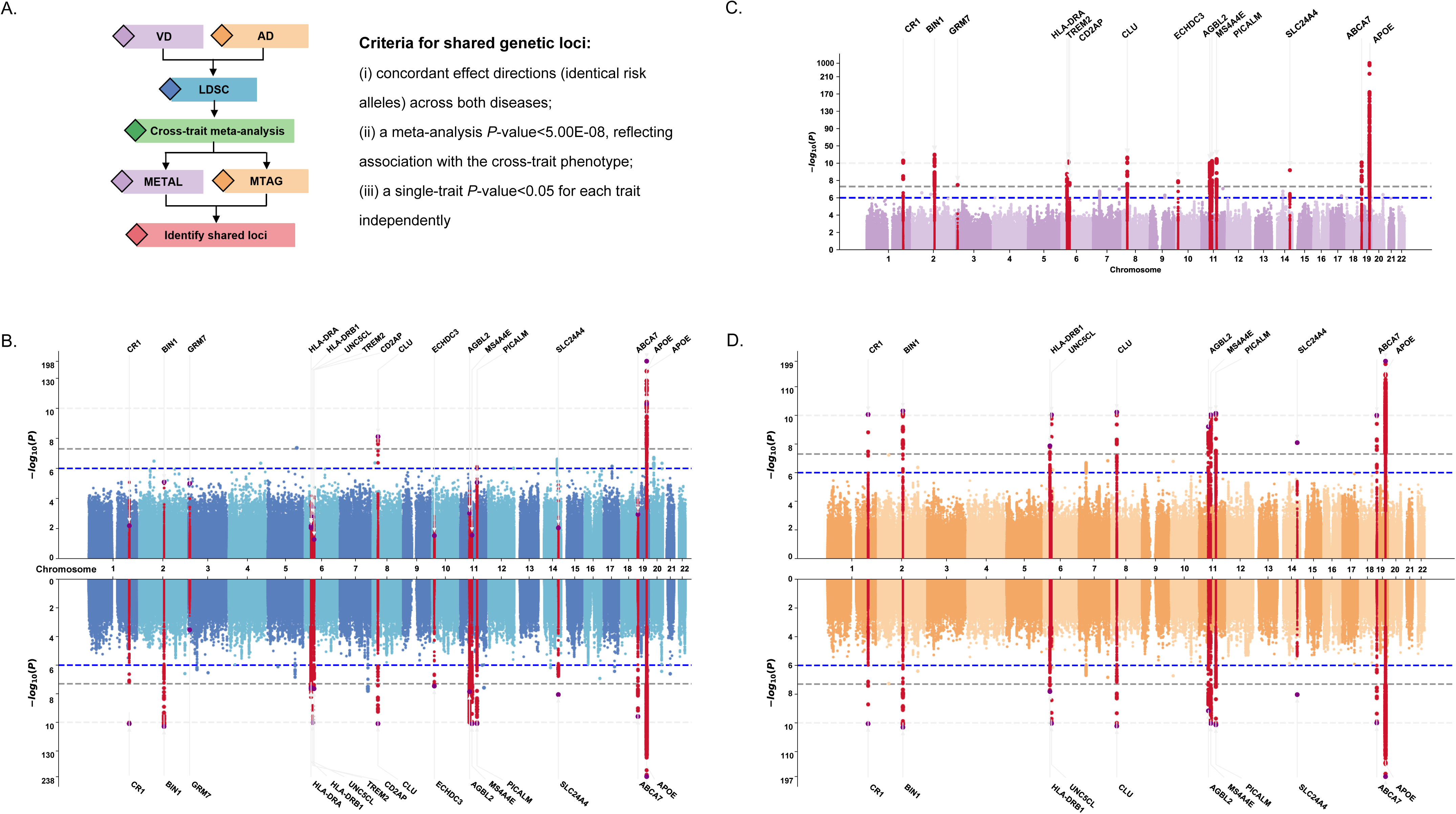
Identification of shared risk loci through multi-trait analysis. **A.** Schematic diagram of cross-trait meta-analysis for identifying shared genetic loci in AD and VD. **B.** Manhattan plots from the VD (upwards) and AD GWAS (downwards) meta-analysis. **C.** Manhattan plots from the VD-AD (cross-trait) GWAS meta-analysis performed with METAL. **D.** Manhattan plots from the VD (upwards) and AD GWAS (downwards) meta-analysis performed with MTAG. In B, C,and D, the horizontal axis shows the chromosomal position (chromosomes 1-22) and the vertical axis shows the significance (-log_10_ *P* value) of tested markers. *P* values are two-sided and based on an inverse variance weighted (IVW) fixed effects meta-analysis. Each dot represents a genetic variant. The threshold for genome-wide significance (*P*<5.00E-08) is indicated by a grey dotted line, and genome-wide significance loci are shown in red. *P* values are two-sided and derived from an inverse-variance-weighted fixed-effects meta-analysis.

We conducted the gene-based association test, gene set enrichment analysis, and tissue enrichment analysis of the cross-trait GWAS meta-analysis summary data using MAGMA ^31^. Gene-based association test identified 127 statistically significant genes with the Benjamini-Hochberg FDR-corrected *P*<0.05 (Fig. 3A, Supplementary Table 34). 44 of these 127 genes were ranked within the top 10% of polygenic priority scores (range: 0.37–4.22), indicating their high posterior probability of being functionally relevant in VD (Fig 3A, Supplementary Table 35) ^32^. *APOE* and *CLU* were ranked with the highest and 4^th^ polygenic priority scores, respectively (Supplementary Table 35).

Gene set enrichment analysis identified 49 statistically significant GO pathways with the Benjamini-Hochberg FDR-corrected *P*<0.05 including neurofibrillary tangle (GO:0097418, *P*=5.99E-11, and FDR=1.02E-06), negative regulation of endopeptidase activity (GO:0010951), autophagic cell death (GO:0048102), positive regulation of complement activation (GO:0045917), regulation of antigen processing and presentation of peptide or polysaccharide antigen via MHC class II (GO:0002580), negative regulation of amyloid precursor protein catabolic process (GO:1902992), amyloid-beta clearance by cellular catabolic process (GO:0150094, *P*=1.31E-06, and FDR=2.69E-03), positive regulation of amyloid-beta clearance (GO:1900223), positive regulation of high-density lipoprotein particle clearance (GO:0010983), and regulation of amyloid fibril formation (GO:1905906, *P*=2.18E-06, and FDR=3.24E-03), as the top 10 significant signals (Fig. 3C, Supplementary Table 36). Tissue enrichment analysis identified evidence of enrichment of VD heritability in GTEx v8 liver (*P*=1.08E-04), blood (*P*=7.56E-03), lung (*P*=2.68E-02), spleen (*P*=3.23E-02), and small intestine (*P*=4.54E-02), but not in human brain tissues (Fig. 3D, Supplementary Table 37).

### Gene prioritization

To identify the potential causal genes, we integrated evidence from 29 complementary approaches: GWAS significance (*P*<5.00E-08), gene mapping (position mapping, chromatin interaction mapping, or eQTLs mapping), variant annotation (exonic SNPs, CADD/RDB/pLI), gene-based association test (*P_FDR_*<0.05), PoPS (top 10%), TWAS (*P_FDR_*<0.05), colocalization (*PPH_4_*>0.70), SMR (eQTLs/sc-QTLs, *P_FDR_*<0.05), drug-gene interaction, gene expression in brain cells (|Log_2_FC|>0.5 and *P_FDR_*<0.05). We calculated the priority scores ranging from 1 to 29 (Supplementary Table 38). Finally, 14 of the 619 protein-coding genes were supported by at least 15 lines of evidence, including *CLU* and other 13 genes around the *APOE* region (Fig. 6).

**Fig 6.**
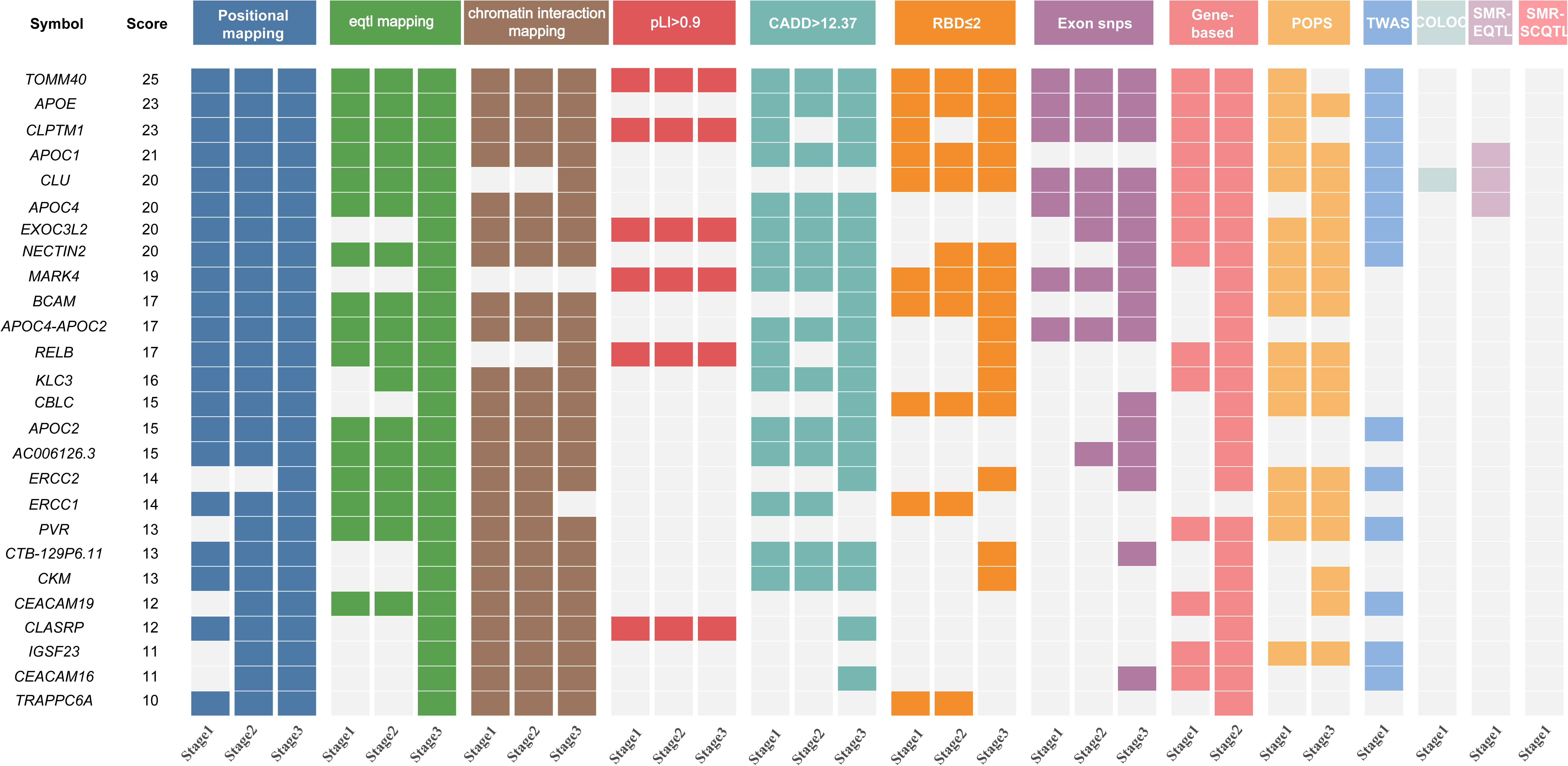
Summary of evidence for vascular dementia priority genes (priority score≥10). A total of 29 lines of evidence were used for gene prioritization, and each column represents a type of supportive evidence. The left table displays the gene symbol and the total score across all evidence categories. The right heatmap groups and colors evidence categories according to their respective domains. Only genes with a priority score≥10 are shown in this figure, and the full results can be found in Supplementary Table 36. pLI, probability of being loss-of-function intolerant; CADD, combined annotation-dependent depletion; RDB, RegulomeDB; PoPS, polygenic priority score; TWAS, transcriptome-wide association study; COLOC, colocalization;,SMR, summary-data-based Mendelian randomization.

### Differential gene expression analysis in human brain cells

We identified 619 protein-coding genes using VD GWAS (stage 1 and stage 2), gene mapping (stage 1 and stage 2), gene-based association test (stage 1), TWAS (stage 1), SMR (stage 1), cross-trait GWAS meta-analysis, gene mapping (stage 3), and gene-based association test (stage 3) (Supplementary Table 38). To investigate the differential expression of VD genes in human brain cells, we analyzed the single-nucleus RNA-sequencing (snRNA-seq) data in human periventricular white matter from 5 VD patients with lesion, 5 VD patients adjacent to the lesion, and 5 normal control subjects (GEO accession: GSE213897, Methods) ^42^, 4 VD patients and 4 age and sex-matched healthy controls (GEO accession: GSE282111, Methods) ^43^. All single cells in GSE282111 were classified into 30 clusters and 6 primary cell types astrocytes, endothelial cells, microglia, neurons, oligodendrocytes and oligodendrocyte precursor cells (OPCs) (Fig. 7A-B). 89 and 201 genes including 49 shared genes exhibited significantly differential expression in VD patients compared to normal controls in at least one cell type with |Log_2_fold change (FC)|>0.5 and *P_FDR_*<0.05 in GSE213897 and GSE282111, respectively (Fig. 7C-D, Supplementary Tables 39-40). Briefly, we found significantly differential expression of 24 VD GWAS loci (*DPP6*, *ITFG1*, *RCSD1*, *CHD6*, *PTPRT*, *CLU*, *HPSE2, ADAMTS17*, *ATP11A*, *TBC1D22A*, *TBC1D4*, *ADAMTSL1*, *ADGRV1*, *APOE*, *CDH18*, *PSAT1*, *HERC3*, *ATP1A1*, *CRELD1*, *COL12A1*, *HS6ST1*, *MROH8*, *TMEM87A*, *ATP10A*) and 9 cross-trait GWAS loci (*SLC24A4*, *CD2AP*, *PICALM*, *CLU*, *HLA-DRA*, *APOE*, *BIN1*, *ABCA7*, *TREM2*, *MS4A4E*) (Fig. 7E-Q, Supplementary Tables 39-40, Supplementary Fig. 4). Meanwhile, *CALCRL* from gene-based association test, *WWOX*, *PARD3*, *AFF1* and *FBXW8* from TWAS also showed significantly differential expression.

**Fig 7.**
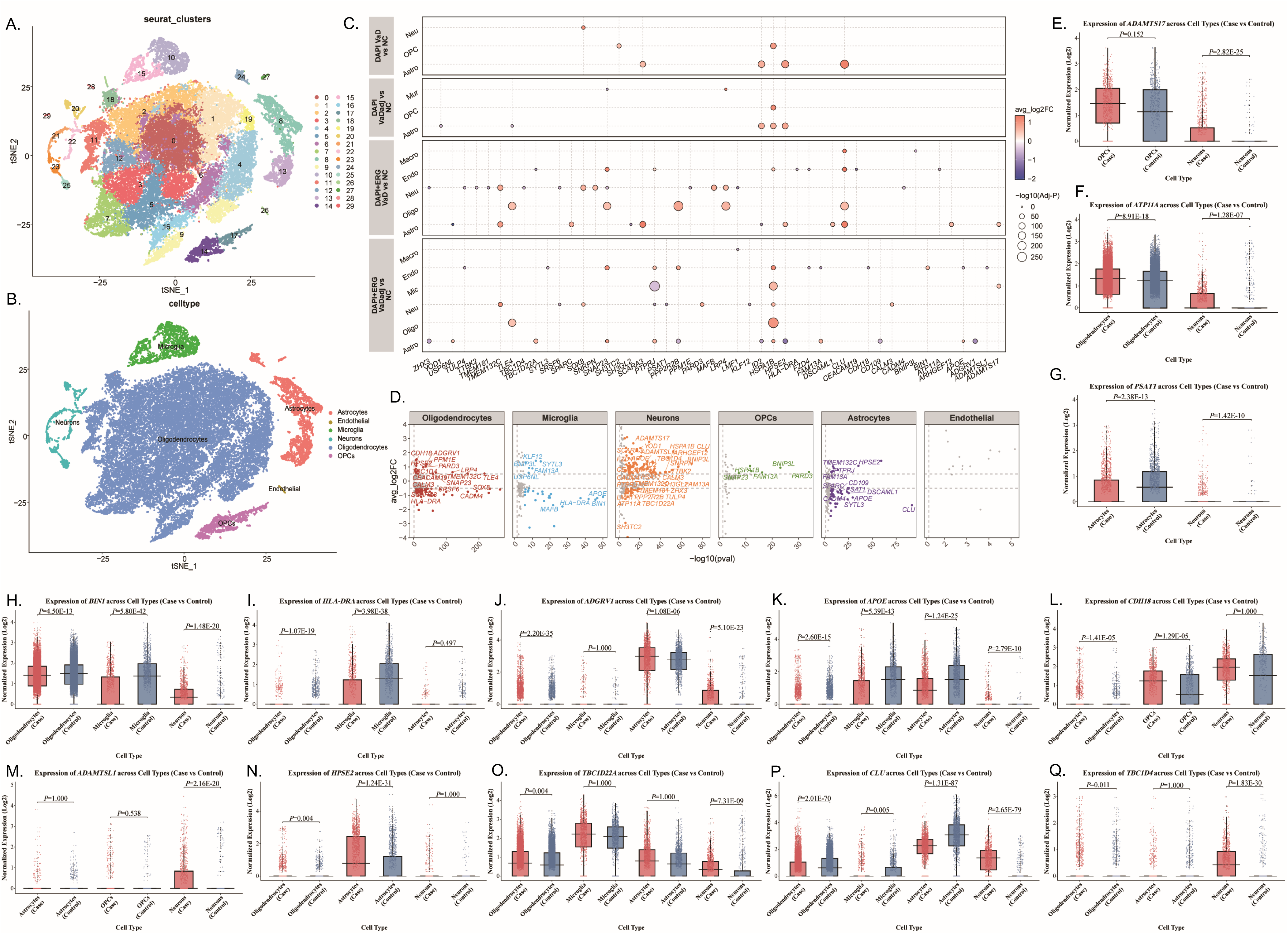
Single-cell validation of differentially expressed genes across vascular dementia. **A-B.** The UMAP plot illustrates the comprehensive annotation of VD and control samples into 6 distinct cell types in GSE282111, with each color representing a specific cell type. **C.** Grouped scatter plot validates the differential expression of 49 genes identified as shared between the GSE213897 and GSE282111 datasets, across distinct cell types within the GSE213897 single-cell RNA sequencing data. Panels correspond to four independent disease-control comparisons: DAPI VaD vs. NC, DAPI VaDadj vs. NC, DAPI+ERG VaD vs. NC, and DAPI+ERG VaDadj vs. NC. In each panel, the y-axis denotes individual cell types, while the x-axis represents the gene symbol. Each point signifies the expression change of a single gene within a specific cell type. **D.** Grouped volcano plot validates the differential expression of the 49 genes shared between the GSE213897 and GSE282111 datasets, across various cell types in the GSE282111 single-cell RNA sequencing data. The six panels correspond to six major cell types. In each panel, the x-axis represents the statistical significance of expression change (-log10 adjusted *P-*value), and the y-axis represents the magnitude and direction of change (log_2_ fold change). Each point represents a single gene within the specified cell type, visually confirming its dysregulation in the VD. **E.** Box plot of *ADAMTS17* from differential expression analysis using GSE282111; **F.** Box plot of *ATP11A* from differential expression analysis using GSE282111; **G.** Box plot of *PSAT1* from differential expression analysis using GSE282111; **H.** Box plot of *BIN1* from differential expression analysis using GSE282111; **I.** Box plot of *HLA-DRA* from differential expression analysis using GSE282111; **J.** Box plot of *ADGRV1* from differential expression analysis using GSE282111; **K.** Box plot of *APOE* from differential expression analysis using GSE282111; **L.** Box plot of *CDH18* from differential expression analysis using GSE282111; **M.** Box plot of *ADAMTSL1* from differential expression analysis using GSE282111; **N.** Box plot of *HPSE2* from differential expression analysis using GSE282111; **O.** Box plot of *TBC1D22A* from differential expression analysis using GSE282111; **P.** Box plot of *CLU* from differential expression analysis using GSE282111; **Q.** Box plot of *TBC1D4* from differential expression analysis using GSE282111;

### Drug-gene interaction analysis

To investigate whether these significantly differentially expressed genes are potential therapeutic targets, we examined the interactions between these genes and known drugs using the Drug-Gene Interaction Database 5.0 (DGIdb, https://dgidb.org) ^44^. Among 241 significantly differentially expressed genes, 21 showed strong evidence of interaction with U.S. Food and Drug Administration (FDA) approved drugs with interaction score>1, which highlighted the potential clinical utility of these genes (Supplementary Table 41). *CDH18*, the VD GWAS significant locus, indicated the strongest interaction (interaction score=26.25) with Thiamine (vitamin B1). *ATP1A1*, the VD GWAS significant locus, indicated the strong interaction with Almitrine (interaction score=10.50). *HSD3B1*, the VD GWAS significant locus, indicated the strong interaction with Trilostane (interaction score=8.75). *NT5E*, the VD GWAS significant locus, indicated the strong interaction with Tinidazole (interaction score=5.53), which is used to treat infections caused by protozoa. *ATP10A*, the VD GWAS significant locus, indicated the strong interaction with Duloxetine (interaction score=20.2), which is used to treat depression and anxiety. *SLC24A4*, the cross-trait GWAS significant locus, indicated the strong interaction with salbutamol (interaction score=8.08), which was approved to treat asthma and chronic obstructive pulmonary disease (COPD).

## Discussion

Until now, *APOE* is the only genome-wide significant locus identified by recent VD GWAS from MEGAVCID consortium ^7^. Here, we performed the largest VD GWAS meta-analysis to date in 1,033,769 individuals including 5,886 VD patients and 1,027,883 controls from five ancestral populations: European, East Asian, South Asian, African, and Admixed American. We identified 37 independent genome-wide significant loci including *APOE* and *CLU* tagged by common variants and 35 loci tagged by rare variants, which explained 53% of VD variance. Gene-based association test, TWAS, SMR and gene set enrichment analysis identified statistically significant 18, 25, 4 genes and 2 pathways, respectively. Cross-trait meta-analysis of VD and AD identified 13 independent genome-wide significant loci, 127 statistically significant genes, 49 statistically significant pathways. These findings provide novel insights into the genetic basis of VD and new leads for the molecular mechanisms underlying VD. Until now, the exact roles of *APOE* and *CLU* in VD remain unclear, however their roles have been investigated in AD. *APOE* is a well-known genome-wide significant locus associated with multiple dementias including AD ^22^, FTD ^23^, and LBD ^24,45,46^. *CLU*, which codes clusterin protein, was a known genome-wide significant locus for AD ^22^. It is known that VD is caused by reduced blood flow to the brain, which damages and eventually kills brain cells ^2^. Loss of clusterin shifts amyloid deposition to the cerebrovasculature, and promotes cerebrovascular cerebral amyloid angiopathy (CAA), which is a neurological condition where amyloid-beta protein deposits in the walls of cerebral blood vessels ^47,48^. *CLU* ameliorates diabetic atherosclerosis by inhibiting the release of inflammatory factors and macrophage pyroptosis ^49^.

In addition to *APOE* and *CLU*, we identified *CALCRL* and *WNK1* as two additional novel VD genes using gene-based association test. *CALCRL* (calcitonin receptor like receptor) is a major G-protein-coupled neuropeptide receptor for both adrenomedullin and calcitonin gene-related peptide (CGRP), which contribute to widen blood vessels, allowing more blood to flow through ^50^. TWAS revealed that genetically decreased arterial expression of *CALCRL* and higher abundance in blood were associated with increased white matter hyperintensities (WMH), a MRI marker of cerebral small vessel disease (CSVD) that is the most common pathology underlying VD ^51,52^. *CALCRL* was a known genome-wide significant locus for ischemic stroke ^53^. Mendelian randomization showed that higher expression of CALCRL in the brain tissues was linked to larger WMH burden and AD risk ^54^. *WNK1* plays an important role in regulating blood pressure and vasoconstriction ^55,56^. Inactivation of mouse *Wnk1* in mature neurons leads to axon degeneration in the adult brain, and *WNK1* may have neuroprotective role in kinds of neurodegenerative diseases ^57^.

*PARD3*, TWAS significant gene in aorta artery, regulates the trafficking and processing of amyloid precursor protein ^58^. Loss of Par3 promotes dendritic spine neoteny and enhances learning and memory ^59^. Hippocampal atrophy is a recognized biological marker of AD ^60^. *FBXW8*, TWAS significant gene in 24 tissues, is a known genome-wide significant locus for hippocampal volume ^60^. *WWOX*, TWAS significant gene in 24 tissues, is a known genome-wide significant locus for AD ^61^. *WDR27*, TWAS significant gene in 41 tissues, is associated with a higher genetic risk for AD and related dementia ^62^. The combination of *WDR27* variants *UNC93A* variants can impair the function of the neurovascular unit (which includes brain blood vessels) and contribute to the development of dementia ^62^.

*ATP1A1*, a novel VD GWAS significant locus tagged by rare variant, is associated with thrombosis and hypertension. *ATP1A1* haplodeficiency or inhibition significantly inhibited thrombosis and sensitized clopidogrel’s anti-thrombotic effect ^63^. *ATP1A1* somatic mutations can lead to aldosterone-producing adenomas, causing excess aldosterone and high blood pressure ^64^. *ATP1A1* genetic variant was associated with essential hypertension ^65^. An *ATP1A1*-related long non-coding RNA, *ATP1A1-AS1*, is implicated in the development of intracranial aneurysms by promoting smooth muscle cells phenotype switching and apoptosis ^66^. *CDH18*, the VD GWAS significant locus tagged by rare variant, regulates differentiation towards vascular smooth muscle cells ^67^, and shows the most significantly upregulated protein expression in the good prognosis group of moyamoya disease, a rare cerebrovascular disorder ^68^. *ATP10A*, the novel VD GWAS significant locus tagged by rare variant, showed a positive association with TDP-43 protein level, the accumulation of which in the central nervous system is a hallmark of frontotemporal lobar degeneration and amyotrophic lateral sclerosis ^69^. *ATP10A* indicated lower expression in AD endothelial cells ^70^. Gene-set enrichment analysis of VD GWAS highlighted the involvement of neurofibrillary tangle (GO:0097418), late endosome (GO:0005770), positive regulation of amyloid fibril formation (GO:1905908), NMDA glutamate receptor clustering (GO:0097114), regulation of amyloid fibril formation (GO:1905906), amyloid-beta clearance by cellular catabolic process (GO:0150094), negative regulation of amyloid fibril formation (GO:1905907). It is known that AD is characterised by both amyloid-β plaques and neurofibrillary tangles, which suggests that VD may coexist with AD. Interestingly, our genetic correlation analysis supported the statistically significant positive genetic correlation between VD and AD with *rg*=0.4469 and *P*=3.75E-14. In fact, population study demonstrated that amyloid-β plaques and neurofibrillary tangles have the strongest association with dementia including AD, VD, or both ^71^, and midlife vascular risk factors was significantly associated with later-life elevated amyloid-β plaques ^72^. Meanwhile, endosome dysfunction was involved in AD and other neurodegenerative diseases, and targeting endosome may be a strategy for treatment ^73^.

We further conducted a cross-trait GWAs meta-analysis of VD and AD, and identified 13 loci, 127 genes, and 53 pathways. It is noted that *GRM7* is the only novel cross-trait GWAS locus that was not previously reported to be associated with VD or AD ^22^. The subcellular-resolution spatial transcriptome atlas of the human prefrontal cortex revealed the increase of *GRM7* expression in the severe AD group compared to the moderate AD group ^74^. Biallelic *GRM7* variants cause epilepsy, microcephaly, and cerebral atrophy ^75^. *GRM7* prevents glutamate release from pre-synaptic vesicles ^76^. *GRM7* variants predict the risk of schizophrenia and antipsychotic effect of seven common drugs ^77^.

Using scRNA-seq data, we demonstrated significantly differential expression of 241 VD genes including 24 VD GWAS loci and 9 cross-trait GWAS loci especially *CLU*, *CDH18*, *ATP1A1*, and *SLC24A4*. Interestingly, evidence supported the dysregulation of these genes in other neurological diseases. *CLU* was significantly downregulated in dementia with Lewy bodies (DLB) microglia (Log_2_FC=-3.76 and *P_FDR_*=2.87E-08), and OPC (Log_2_FC=-1.42 and *P_FDR_*=3.08E-02), downregulated in Parkinson’s disease with dementia (PDD) excitatory neuron (Log_2_FC=-1.34 and *P_FDR_*=1.30E-27), microglia (Log_2_FC=-1.36 and *P_FDR_*=4.12E-02), and OPC (Log_2_FC=-1.16 and *P_FDR_*=7.60E-04), upregulated in Parkinson’s disease (PD) excitatory neuron (Log_2_FC=0.91 and *P_FDR_*=5.85E-116), inhibitory neuron (Log_2_FC=0.75 and *P_FDR_*=8.03E-24), oligodendrocyte (Log_2_FC=1.15 and *P_FDR_*=3.86E-29) ^78^, dopaminergic neuron (Log_2_FC=-1.58 and *P_FDR_*=1.12E-03), OPC (Log_2_FC=-1.12 and *P_FDR_*=3.40.E-02), and pericyte (Log_2_FC=-4.57 and *P_FDR_*=1.08.E-10) ^79^. *CDH18* was significantly upregulated in DLB excitatory neuron (Log_2_FC=1.37 and *P_FDR_*=3.26E-08), and PDD excitatory neuron (Log_2_FC=0.94 and *P_FDR_*=3.00E-09), but downregulated in PD excitatory neuron (Log_2_FC=-0.96 and *P_FDR_*=8.64E-15) ^78^. *ATP1A1* was significantly downregulated in DLB (astrocyte, excitatory neuron, inhibitory neuron, microglia, oligodendrocyte, OPC, vascular), PDD (astrocyte, excitatory neuron) ^78^, and dopaminergic neuron (Log_2_FC=-2.6 and *P_FDR_*=1.49.E-02)^79^. *SLC24A4* was significantly downregulated in DLB excitatory neuron (Log_2_FC=-0.66 and *P_FDR_*=4.20E-04) ^78^. Meanwhile *APOE* was significantly upregulated in dementia with DLB astrocyte (Log_2_FC=1.12 and *P_FDR_*=2.42E-38) ^78^, and PD microglia (Log_2_FC=1.748 and *P_FDR_*=2.76.E-02) ^79^.

Drug-gene interaction analysis highlights *APOE*, *CDH18*, *ATP1A1*, *HSD3B1*, *NT5E*, *ATP10A*, and *SLC24A4*, as the potential therapeutic targets for VD (Supplementary Table 13). *APOE* is the target of 37 drugs especially FDA approved drugs for the treatment of AD including lecanemab ^80^, donepezil ^81^, rivastigmine ^82^, and galantamine ^83^. Importantly, randomized controlled trials (RCTs) demonstrated that donepezil and galantamine effectively improved cognition in VD patients with good safety and tolerability ^84–86^. *CDH18* indicated the strongest interaction (interaction score=26.25) with thiamine (vitamin B1). Two cross-sectional observational studies using data from the National Health and Nutrition Examination Survey (NHANES) showed that the increase in dietary intake of vitamin B1 contribute to better cognitive function in individuals aged over 60, and a decreased risk of stroke in older individuals ^87,88^. In cognitively healthy and older Chinese individuals, there is a J-shaped association between dietary vitamin B1 intake and cognitive decline ^89^. These findings highlight the potential importance of adequate dietary vitamin B1 intake to prevent cognitive decline and stroke in the aging population.

*ATP1A1* shows the strong interaction with almitrine (interaction score=10.50), which was approved to treat chronic obstructive lung disease. A meta-analysis of three RCTs showed that Duxil (a combination of almitrine and raubasine) significantly improved the cognitive function in VD patients measured by MMSE ^90^. *ATP10A* showed a strong interaction with Duloxetine (interaction score=20.2), a type of antidepressant medicine to treat depression and anxiety ^91^. Recent study identified duloxetine as a highly potent selective competitive inhibitor of butyrylcholinesterase, which was involved in the regulation of the nervous system that affects memory and cognition, and may have positive effects on memory and cognitive functions in the elderly ^91^. A RCT demonstrated that duloxetine was effective to treat patients with moderate to severe central post-stroke pain ^92^. *SLC24A4* indicated the strong interaction with salbutamol (interaction score=8.08), which was approved to treat asthma and COPD. Interestingly, salbutamol is effective at reducing the accumulation and rate of the tau protein formation ^93^. Our current findings broaden the potential therapeutic scope of the available drugs, and may offer potential as a new treatment for VD.

Tissue enrichment analysis showed evidence of enrichment of VD heritability in lung and spleen, and VD+AD heritability in liver, blood, lung, spleen, and small intestine. In fact, large-scale GWAS showed that AD heritability was enriched in whole blood, spleen and lung ^94^. Population-based cohort studies from the Atherosclerosis Risk in Communities (ARIC) study ^95–97^, the Rotterdam study ^98^, the Rush Memory and Aging Project (MAP) ^99^, Swedish National Study on Aging and Care in Kungsholmen (SNAC□K, 2001-2004 to 2016-2019) ^100^, and Health and Retirement study ^101^ collectively demonstrated that poor lung function increased the risk of both mild cognitive impairment (MCI) and dementia, accelerated progression from MCI to dementia, associated with AD pathology and cerebral vascular disease pathology, brain microvascular damage and global brain atrophy. Impaired lung function, defined as peak expiratory flow<80% predicted, was associated with a higher risk of dementia (HR=1.74, 95% CI:1.34-2.25) ^101^. Compared to those without impaired lung function, individuals with impaired lung function had 0.10 SD higher NfL and 0.09 SD higher p-Tau 181, which mediated 7.3% and 5% of the total effect of impaired lung function on dementia ^101^. A prospective cohort study of 431,834 non-demented individuals from the UK Biobank indicated that lung function decrease was associated with increased risk for all-cause dementia, AD and VD ^102^.

Together, our large-scale GWAS meta-analysis and integrative analysis uncovered novel VD loci, genes and pathways. Our current findings demonstrated that the use of both common and rare genetic variants in large-scale VD GWAS could (1) enhance the ability to identify new loci, (2) identify rare variants of large effects, and (3) increase the proportion of VD heritability. These genetic findings provide valuable insights into the potential underlying mechanisms of VD and inform some potential clinically actionable drugs for the treatment of VD, which deserve further investigation.

## Materials and Methods

### VD GWAS datasets

We selected four independent GWAS datasets from UKBB (European, 2,074 VD and 456,366 controls) ^15^, FinnGen R12 (European, 3,624 VD and 475,484 controls) ^16^, GH (South Asian, 119 VD and 43,659 controls) ^26^, and MGBB (European, South Asian, African, and Admixed American, 69 VD and 52,374 controls) ^27^.

UKBB cohort represents a population-based, longitudinal study of over 500,000 volunteers aged 40-70 years, recruited from England, Scotland, and Wales between 2006 and 2010 ^103^. Extensive participant data were gathered through surveys, interviews, physiological assessments, and genetic profiling ^103^. VD GWAS included 2,074 VD and 456,366 controls of non-Finnish European ancestry about 16,477,695 variants with position information from the Genome Reference Consortium Human Build 37 (GRCh37) ^15^. VD were diagnosed using the available medical records, which were ascertained from Hospital Episode Statistics and recorded as International Classification of Disease version 10 (ICD-10) codes ^15^.

FinnGen study encompasses six regional and three nationwide Finnish biobanks, with participants’ health outcomes meticulously tracked through linkages to the comprehensive national health registries, spanning from birth to death ^16^. Using ICD-9 and ICD-10 codes, 3,624 individuals were identified as VD cases, and 475,484 individuals were identified as controls in FinnGen R12 ^16^. FinnGen VD GWAS included 21,306,039 genetic variants with position information from GRCh38 ^16^. Here, we converted the position information from GRCh38 to GRCh37. After a rigorous quality control process to eliminate mismatched variants between GRCh38 and GRCh37, we obtained 13,092,808 genetic variants for meta-analysis.

GH is a community-based population genomics and health study comprising about 50,000 British individuals of South Asian ancestry (British Bangladeshi and British Pakistani) recruited in the United Kingdom including East London and Bradford ^26^. GH identified 119 VD cases and 43,659 controls using ICD-10, and included 37,686,810 genetic variants with position information from GRCh38 ^26^.

MGBB was established based on Mass General Brigham, an integrated healthcare system based in the Greater Boston area of Massachusetts, annually serves 1.5 million patients. MGBB currently included 142,238 participants from European, South Asian, African, and Admixed American ancestries with 69 VD and 52,305 controls ^27^.

### GWAS meta-analysis

GWAS meta-analysis was performed using the fixed-effects IVW implemented by METAL, weighted by effect size and standard error (SE), with genomic control correction ^17^. In Stage 1, a GWAS meta-analysis was performed in participants of European ancestry from UKBB and FinnGen R12. In stage 2, a GWAS meta-analysis was performed in participants of European, South Asian, African, and Admixed American ancestries from UKBB, FinnGen R12, GH and MGBB. In addition to METAL, a sensitivity GWAS meta-analysis using GWAMA (v2.2.2) was performed ^21^. To assess the genomic inflation, we calculated the genomic inflation factor (λ_GC_) using LDSC (v1.0.1) ^18^. We defined the statistically significant SNPs using the genome-wide significant threshold (*P*<5.00E-08).

### Identification of genetic risk loci

To identify genetic risk loci from the GWAS meta□analysis and perform functional annotation, we used FUMA v1.5.2 with ancestry-specific reference panels: the 1000 Genomes Phase 3 European panel for stage 1 European-specific GWAS meta-analysis, and the 1000 Genomes Phase 3 ALL panel for stage 2 cross-ancestry GWAS meta-analysis ^20^. Initially, FUMA identified SNPs with a significance level of *P*<5.00E-08 and LD threshold *r*^2^<0.6 as the independent significant SNPs using LD-based clumping ^20^. The independent genetic risk loci were characterized by considering all SNPs in LD (*r*^2^≥0.6) with one of the independent significant SNPs within a region of 250 kilobase (kb) ^20^. Within each genetic risk locus, FUMA further distinguished the lead SNPs, which are a subset of the independent significant SNPs in LD with each other (*r*^2^<0.1) ^20^. Each locus was represented by the lead SNP with the most significant *P* value. Genetic loci were classified as novel if they were beyond a 1,000 kb from previously recognized loci ^20^. The nearest gene corresponding to the lead SNPs in each locus was annotated using the get_nearest_gene () function from the gwasRtools package (https://github.com/lcpilling/gwasRtools).

### Conditional analysis

To identify independent genetic signals at each genomic locus, we performed a step-wise conditional analysis of the VD GWAS meta-analysis summary statistics using GCTA (v 1.94.1) COJO analysis with the --cojo-slct function ^25^. The parameters set for this function included a significance threshold of *P*=5.00E-08, a distance of 10,000 kb, and a co-linearity threshold of 0.9. LD information was from the 1000 Genomes European reference panel ^25^.

### Heritability analysis

We used LDSC (v1.0.1) to calculate the SNP-based heritability of VD GWAS meta-analysis summary statistics ^18^. LDSC is a computationally efficient tool that leverages GWAS summary statistics to estimate the SNP-based heritability and genetic correlation among multiple genetic traits, while accounting for potential sample overlap ^18^. The SNP-based heritability measures the proportion of phenotypic variance explained by the additive effects of all common SNPs (i.e, the proportion of variance in disease liability due to genetic factors) ^18^. LD scores are from the 1000 genomes phase 3 European reference panel (https://data.broadinstitute.org/alkesgroup/) using a population prevalence estimate 1.16% ^19^.

### Functional annotation

FUMA offers a comprehensive annotation framework by integrating diverse external data sources including ANNOVAR ^28^, CADD ^30^, RegulomeDB ^104^ and 15-core chromatin state ^105,106^. Here, we conducted a functional annotation of all genome-wide significant SNPs (or independent significant SNPs) and their tagged SNPs with LD *r*^2^≥0.6 using FUMA v1.5.2 and 1000 Genomes European reference panel ^20^.

### Gene mapping

We assigned the genome-wide significant loci to specific genes using positional mapping, eQTLs mapping, and chromatin interaction mapping implemented in FUMA v1.5.2 ^20^. Positional mapping pinpointed protein-coding genes located within a 10 kb range of significant SNPs (either genome-wide significant or independently significant) ^20^. eQTLs mapping assigned the significant SNPs to their corresponding protein-coding genes using significant eQTLs (*P_FDR_*<0.05) ^20^. eQTLs datasets are from the eQTLGen ^38^, Blood eQTL ^107^, BIOS QTL ^108^, GTEx v8 ^109^, PsychENCODE^110^, xQTLServer ^36^, CMC ^111^, eQTL catalogue ^112^ and BRAINEAC ^113^.

### Gene-based association test, gene set and tissue enrichment analyses

We performed the gene-based association test, gene set enrichment analysis, and tissue enrichment analysis of stage 1 VD GWAS meta-analysis summary data using MAGMA v1.08 ^31^. MAGMA aggregates SNP-level association statistics into gene scores by mapping all SNPs from VD GWAS meta-analysis to 17,903 protein-coding genes using the SNP-wise mean model, genomic location and boundary information from human reference genome build 37, and the ancestry-matched LD information from the 1000 Genomes Project phase 3 reference panel ^31^. Finally, a gene-based association score was calculated by the aggregate of all SNPs inside each gene ^31^. MAGMA performed a gene set enrichment analysis through competitive analysis to identify the genes in a gene set that are more strongly associated with the phenotype of interest than other genes ^31^. Here, we focused on 16,228 GO terms including biological processes, cellular components and molecular functions from the Molecular Signatures Database (MSigDB) (v7.0, version 2025.1.Hs) ^114^. MAGMA determined whether VD heritability is enriched in specific tissues by integrating the stage 1 VD GWAS meta-analysis summary data with gene expression data from 30 GTEx v8 tissues ^115^. BH-adjusted *P*<0.05 was considered statistically significant for gene-based association test, gene set enrichment analysis and tissue enrichment analysis.

### Genetic association between VD and lung function traits

Using LDSC default parameters and the precomputed LD scores from the 1000 Genomes European reference panel ^18^, we performed a genetic association analysis to investigate the genetic correlation between VD (stage 1) and 6 lung function traits including FEV1, FVC, FEV1/FVC, PEF, asthma and COPD using large scale GWAS summary datasets from UKBB ^15^, as provided in Supplementary Table 18. We defined the statistically significant genetic association using the Bonferroni-corrected threshold *P*<8.33E-03 (0.05/6).

### Polygenic Priority Score

To pinpoint the most likely causal genes at VD GWAS loci, we calculated the polygenic priority score using PoPS (v.0.2) and the gene-level results from MAGMA analysis of stage 1 VD GWAS meta-analysis summary statistics ^32^. PoPS computes gene-level z-scores from GWAS summary statistics with an LD reference panel using MAGMA, and learns trait-relevant gene features from cell-type specific gene expression, biological pathways, and protein-protein interactions ^32^. In each genome-wide significant locus, genes within ±1 Mb of the lead variant were assigned a polygenic priority score. Within each region, genes ranking in the top 10% of the polygenic priority scores indicate a higher probability of being causal for VD.

### Transcriptome-wide association study

We performed a TWAS to identify the genes whose expression levels are significantly associated with VD using FUSION v3 ^33^. TWAS leverages the correlation between genotype and expression to identify eQTLs that modulate gene expression and are associated with the phenotype of interest ^33^. Using FUSION v3, we integrated the VD GWAS meta-analysis dataset with the gene expression data from GTEx v8 in human tissues and cell types, a whole blood eQTLs dataset from YFS (*N*=1,264) ^35^ and a brain dorsolateral prefrontal cortex eQTLs dataset from CMC (*N*=452) ^111^. A Benjamini & Hochberg FDR-corrected threshold of 0.05 was considered statistically significant for each dataset (*P_FDR_*<0.05).

### Colocalization analysis

Using COLOC implemented in FUSION v3, we performed a Bayesian colocalization analysis to identify a subset of TWAS significant genes that had the same single variant associated with both VD and gene expression with a high posterior probability (PP) ^116^. Basically, COLOC calculated five kinds of PPs ^116^. The PP for the null hypothesis (PP.H_0_): neither trait has a genetic association in the region, PP for the first alternative hypothesis (PP.H_1_): only trait 1 has a genetic association in the region, PP for the second alternative hypothesis (PP.H_2_): only trait 2 has a genetic association in the region, PP for the third alternative hypothesis (PP.H_3_): both traits are associated, but with different causal variants, PP for the fourth alternative hypothesis (PP.H_4_): both traits are associated and share a single causal variant ^116^. A PP.H_4_>0.70 indicated evidence of colocalization ^117^.

### Summary-data-based Mendelian randomization

SMR is a complementary method of TWAS to verify the causal role of TWAS genes ^37^. Here, we used SMR v1.3.1 to integrate the VD GWAS meta-analysis dataset with multiple large-scale eQTLs datasets from GTEx tissues ^34^, a large blood eQTLs dataset from the eQTLGen consortium including 31,684 human blood samples ^38^, and a brain eQTLs dataset including 2,865 human brain cortex samples ^39^. We defined the statistically significant SMR genes using *P_FDR_*<0.05 and the heterogeneity in dependent instruments (HEIDI) test for pleiotropy>0.01 ^118^.

### Cross-trait meta-analysis of VD and AD

We selected the largest clinically diagnosed AD GWAS in individuals of European ancestry from the International Genomics of Alzheimer’s Project (IGAP) stage 1 (*n*=63,926) including a total of 9,456,058 common variants and 2,024,574 rare variants ^22^. Using LDSC default parameters and the precomputed LD scores from the 1000 Genomes European reference panel ^18^, we first estimated the genetic correlation between VD (stage 1) and AD ^22^. We further conducted a cross-trait meta-analysis of VD and AD GWAS datasets using a fixed-effects IVW method implemented by METAL ^17^. Meanwhile, we performed a sensitivity cross-trait meta-analysis of AD and VD GWAS datasets using MTAG (1.0.8) ^41^. MTAG extends the standard single-trait GWAS by jointly analyzing multiple genetically related traits, thereby increasing the statistical power to detect pleiotropic loci while accounting for sample overlap ^41^. To identify robust cross-trait associations, our analysis focused on SNPs exhibiting: (i) concordant effect directions (identical risk alleles) across both diseases; (ii) a meta-analysis *P*-value<5.00E-08, reflecting association with the cross-trait phenotype; and (iii) a single-trait *P*-value<0.05 for each trait independently. Independent genomic loci were then defined using FUMA. We also conducted the gene-based association test, gene set enrichment analysis, and tissue enrichment analysis of the cross-trait GWAS meta-analysis summary data using MAGMA ^31^.

### Gene prioritization

To prioritize the most probable causative genes for VD, we integrated 29 lines of evidence including GWAS significance (*P*<5.00E-08), gene mapping (position mapping, eQTLs mapping, and chromatin interaction mapping), variant annotation (exonic SNPs, CADD/RDB/pLI), gene level association test (MAGMA, *P_FDR_*<0.05), PoPS (top 10%), TWAS (*P_FDR_*<0.05), colocalization analysis (COLOC, *PPH_4_*>0.70), SMR (eQTLs/sc-eQTLs, *P_FDR_*<0.05). For each gene, the prioritization score is the sum of multiple lines of evidence by counting “0” if the gene was ‘not prioritized’ and as “1” if the gene was ‘prioritized’. This approach ensured that genes with stronger cumulative evidence were more likely to be causally associated with VD, and reflected a higher degree of confidence in the VD etiology.

### Differential gene expression analysis in human brain cells

We performed a differential gene expression analysis of VD risk genes using snRNA-seq data in human periventricular white matter from 5 VD patients with lesion, 5 VD patients adjacent to the lesion, and 5 normal control subjects (GEO accession: GSE213897) ^42^, 4 VD patients and 4 age and sex-matched healthy controls (GEO accession: GSE282111) ^43^. Using “Seurat” package, we preprocessed and transformed the raw scRNA-seq data, excluding cells with fewer than three genes and fewer than 50 unique features counted per cell. Subsequently, we used the NormalizeData and ScaleData functions to normalize and scale the RNA transcripts per million (TPM). Using “SingleR” package, we annotated the cell types, which were classified into 6 primary cell types including astrocytes, endothelial cells, microglia, neurons, oligodendrocytes and oligodendrocyte precursor cells (OPCs), as well as 30 clusters ^42^. Finally, we conducted the differential expression analysis using Wilcoxon rank sum test. In a specific cell type, we defined the significantly differential expression in VD patients compared to normal controls with |Log_2_FC|>0.5 and *P_FDR_*<0.05.

### Drug-gene interaction analysis

To assess whether VD risk genes could serve as potential therapeutic targets, we performed a drug-gene interaction analysis using DGIdb v5.0 ^44^. DGIdb v5.0 is an online database that integrates information from drug–gene interaction databases (accessed December 2024) ^44^. DGIdb contains over 10,000 genes and 20,000 drugs involved in nearly 70,000 drug-gene interactions or belonging to one of 43 potentially druggable gene categories ^44^. A interaction score is used to rank results in an interaction search result set ^44^.

## Supporting information

Supplemental figures will be used for the link to the file on the preprint site.

Supplemental tables will be used for the link to the file on the preprint site.

Supplemental data1 will be used for the link to the file on the preprint site.

Supplemental data2 will be used for the link to the file on the preprint site.

## Data Availability

https://figshare.com/s/b095198f60ee24954ebe

## Data availability

All relevant data underlying the findings are fully available without restriction. GWAS summary statistics of UK Biobank is available at https://www.ebi.ac.uk/gwas (GCST90473240); GWAS summary statistics of FinnGen is available at https://r12.finngen.fi/; The Mass General Brigham Biobank (MGBB) GWAS summary statistics is available at https://api.kpndataregistry.org/api/d/BPEif3; Genes & Health (GH) GWAS summary statistics is available at https://www.genesandhealth.org/research/gwas-data-downloads; The 1000 Genomes Phase 3 ancestry-specific LD reference are obtained from the FUMA website at https://fuma.ctglab.nl; TWAS weights are available at http://gusevlab.org/projects/fusion/; eQTL summary datasets used for SMR are available at https://yanglab.westlake.edu.cn/software/smr/#eQTLsummarydata; Single cell gene expression datasets are available at GSE213897 (https://www.ncbi.nlm.nih.gov/geo/query/acc.cgi?acc=GSE213897) and GSE282111 (https://www.ncbi.nlm.nih.gov/geo/query/acc.cgi?acc=GSE282111). AD GWAS summary statistics is available at https://www.niagads.org/igap-rv-summary-stats-kunkle-p-value-data, GWAS summary statistics of forced expiratory volume in 1-second (FEV1) is available at https://pan-ukb-us-east-1.s3.amazonaws.com/sumstats_flat_files_tabix/continuous-20150-both_sexes-irnt.tsv.bgz.tbi, GWAS summary statistics of forced vital capacity (FVC) is available at https://pan-ukb-us-east-1.s3.amazonaws.com/sumstats_flat_files_tabix/continuous-20151-both_sexes-irnt.tsv.bgz.tbi, GWAS summary statistics of FEV1/FVC is available at https://pan-ukb-us-east-1.s3.amazonaws.com/sumstats_flat_files_tabix/continuous-FEV1FVC-both_sexes-irnt.tsv.bgz.tbi, GWAS summary statistics of peak expiratory flow (PEF) is available at https://pan-ukb-us-east-1.s3.amazonaws.com/sumstats_flat_files_tabix/continuous-3064-both_sexes-irnt.tsv.bgz.tbi, GWAS summary statistics of asthma is available at https://pan-ukb-us-east-1.s3.amazonaws.com/sumstats_flat_files_tabix/phecode-495-both_sexes.tsv.bgz.tbi, GWAS summary statistics of COPD is available at https://pan-ukb-us-east-1.s3.amazonaws.com/sumstats_flat_files_tabix/categorical-22130-both_sexes-22130.tsv.bgz.tbi.

## Code availability

No custom code was used in this study. As detailed in the Methods section, we used previously released software to generate the analysis and cite them throughout the manuscript references. METAL (March 25, 2011 release) is available at https://csg.sph.umich.edu/abecasis/Metal/; GWAMA (v2.2.2) is available at https://genomics.ut.ee/en/tools; FUMA (1.5.2) is available at https://fuma.ctglab.nl/; LocusZoom is available at http://locuszoom.sph.umich.edu/; MTAG (v1.0.8) is available at https://github-wiki-see.page/m/JonJala/mtag/; GCTA-COJO (v1.94.1) is available at https://yanglab.westlake.edu.cn/software/gcta/; MAGMA (v1.08) is available at https://cncr.nl/research/magma/; LDSC is available at https://github.com/bulik/ldsc; PoPS (v0.2) is available at https://github.com/FinucaneLab/pops; FUSION (v3) is available at http://gusevlab.org/projects/fusion/; SMR (v1.3.1) is available at https://yanglab.westlake.edu.cn/software/smr/; DGIdb (v5.0): https://beta.dgidb.org/.

## Declarations

### Ethics approval and consent to participate

This article involves human individuals from prior investigations. All individuals provided informed consent in all of the corresponding original investigations, as reported in the Materials and methods. Our analysis is based on publicly available, large-scale datasets rather than individual-level data. Thus, ethical approval was not sought.

### Consent for publication

Not applicable.

### Availability of data and materials

All relevant data are within the paper. The authors confirm that all data underlying the findings are either fully available without restriction through consortia websites, or may be made available from consortia upon request.

### Authors’ contributions

The authors declare no competing interests. G.Y.L. conceived and initiated the project. G.Y.L. and G.S analyzed the data and wrote the first draft of the manuscript. All authors contributed to the interpretation of the results and critical revision of the manuscript for important intellectual content and approved the final version of the manuscript.

