## Supplemental figures will be used for the link to the file on the preprint site. for "Genome-wide cross-trait analysis of vascular dementia and Alzheimer’s disease highlights novel loci and lung-brain axis"

### Supplementary Figures

**Supplementary Figure 1.** Quantile-quantile (Q-Q) plots for the GWAS meta-analysis of vascular dementia.

**Supplementary Figure 2.** Regional association plots for three significant locus from the European-specific GWAS meta-analysis of vascular dementia (Stage 1).

**Supplementary Figure 3.** Regional association plots for three significant locus from the Cross-ancestry GWAS meta-analysis of vascular dementia (Stage 2).

**Supplementary Figure 4.** Box plot of genes from differential expression analysis using GSE282111.

**Supplementary Figure 1. Quantile-quantile (Q-Q) plots for the GWAS meta-analysis of vascular dementia.** A) Q-Q plot for association with vascular dementia in Stage 1, Genomic inflation factor ($\lambda_{GC}$) = 1.0754, B) Q-Q plot for association with vascular dementia in Stage 2, $\lambda_{GC}$= 1.0742. The horizontal axis shows -log10 *p* values expected under the null distribution. The vertical axis shows observed -log10 *p* values. The blue framed line shows the 95% confidence interval.

**A.** **B.**

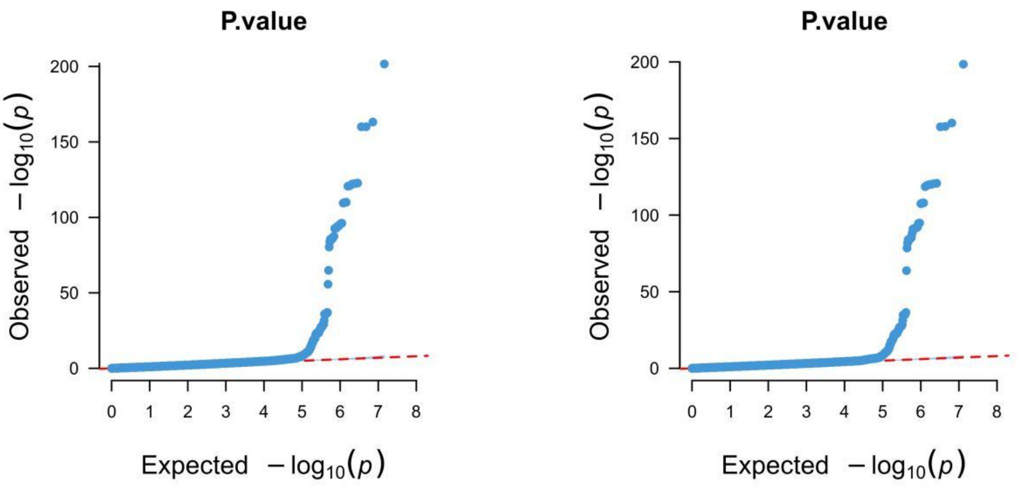

**Supplementary Figure 2. Regional association plots for three significant locus from the European-specific GWAS meta-analysis of vascular dementia (Stage 1).** A) rs564080066 is located on *HTR4*, B) rs7982 is located on *CLU*, C) rs429358 is located on *APOE.* The x-axis represents the shows the chromosomal position of SNPs (hg19). The y-axis represents -log10 p-values from two-sided z-tests for meta-analysis effect estimates. LD estimates of surrounding SNPs with the labeled index SNP (LD r2values estimated based on the 1000 Genomes Phase 3 European panel) is indicated by color (color bar on side of plot indicates color coding of r2). Local estimates of recombination rate are indicated in light blue (legend on vertical axis at right). Gene names, strands, and boundaries are shown in the box below the regional plot.

A.

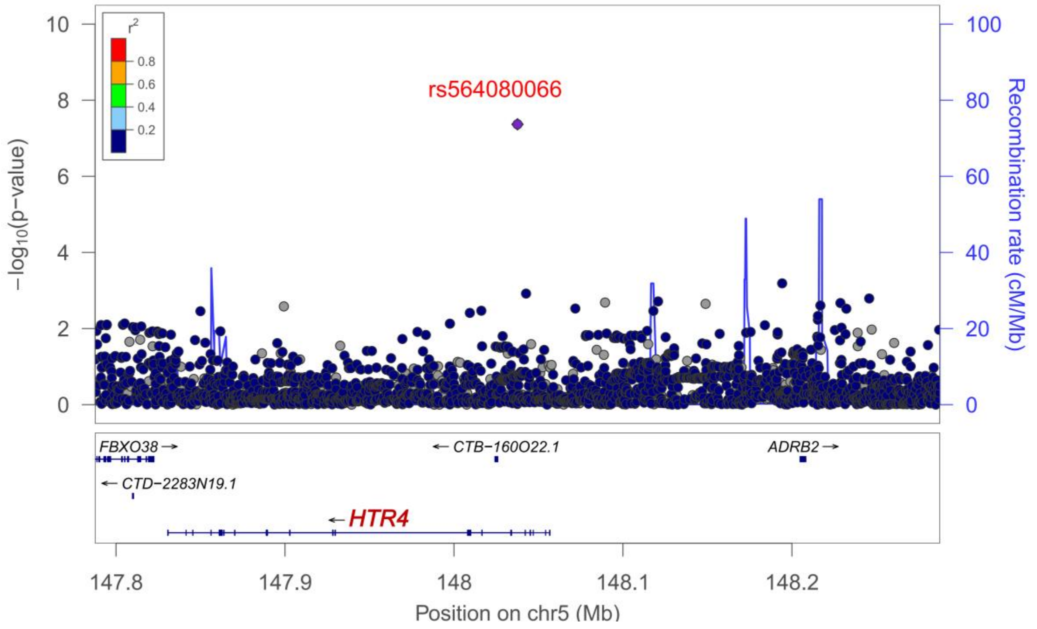


B.

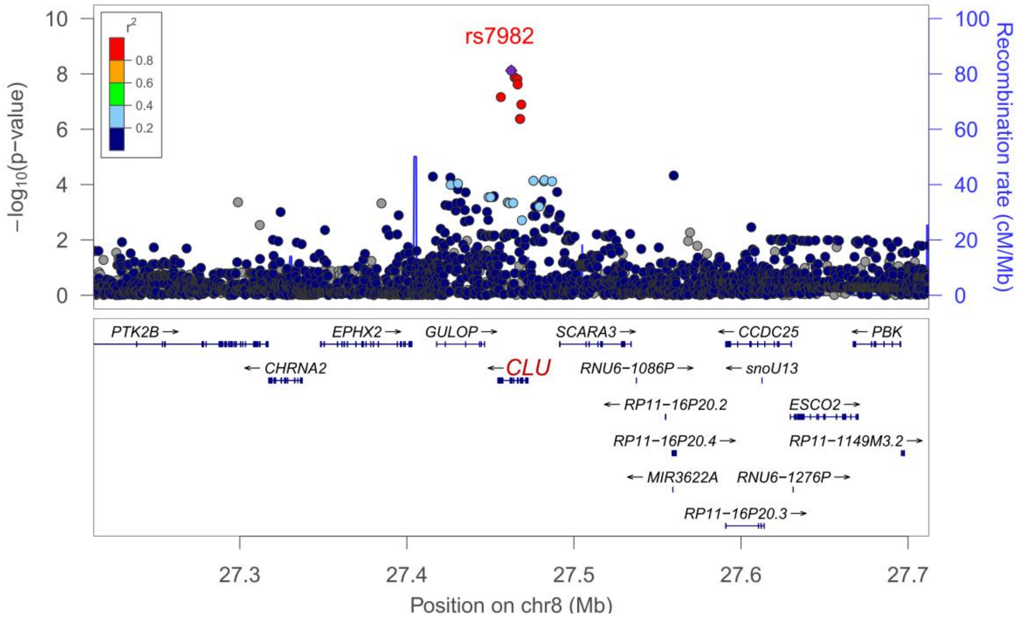

C.

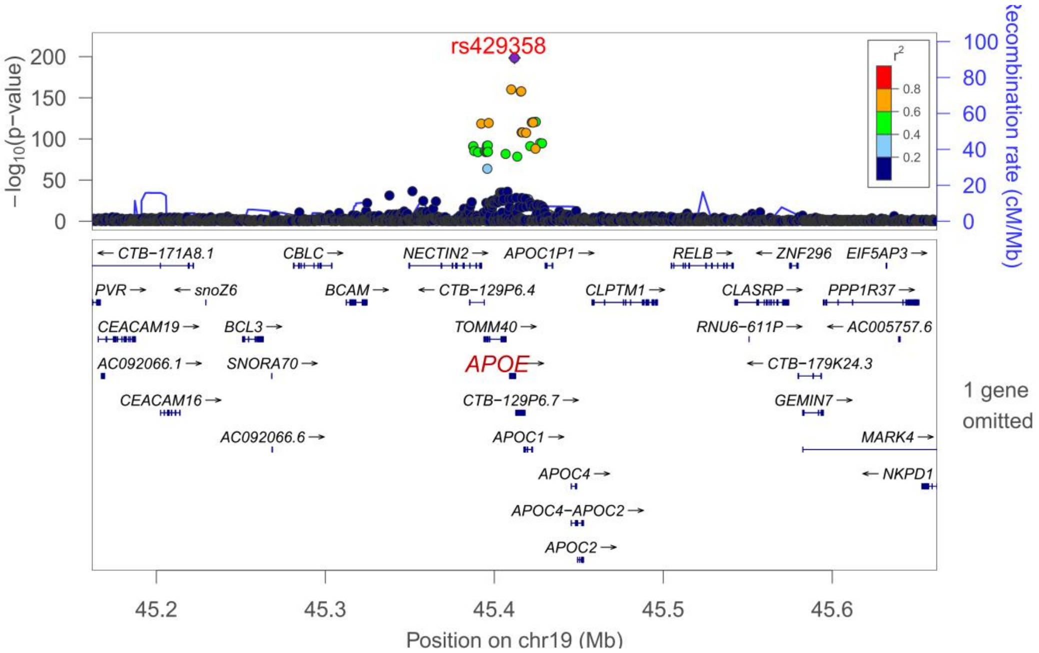

rs429358
−log10(p−value)
Recombination rate (cM/Mb)
r2
0.8
0.6
0.4
0.2
CTB−171A8.1
CBLC
NECTIN2
APOC1P1
RELB
ZNF296
EIF5AP3
PVR
snoZ6
BCAM
CTB−129P6.4
CLPTM1
CLASRP
PPP1R37
CEACAM19
BCL3
TOMM40
RNU6−611P
AC005757.6
AC092066.1
SNORA70
APOE
CTB−179K24.3
CEACAM16
CTB−129P6.7
GEMIN7
AC092066.6
APOC1
MARK4
APOC4
NKPD1
APOC4−APOC2
APOC2
1 gene omitted
45.2
45.3
45.4
45.5
45.6
Position on chr19 (Mb)

**Supplementary Figure 3. Regional association plots for thirty-seven significant locus from the Cross-ancestry GWAS meta-analysis of vascular dementia (Stage 2).** A) rs150423973 is located on *ATP1A1*, B) rs55977072 is located on *HSD3B1*, C) rs555863053 is located on *RCSD1*, D) rs543737714 is located on *RSAD2*, E) rs571573246 is located on *HS6ST1*, F) rs138654898 is located on *UBR3*, G) rs536931879 is located on *CRELD1*, H) rs79120584 is located on *HERC3*, I) rs534723021 is located on *CDH18*, J) rs200249535 is located on *SPZ1*, K) rs563370505 is located on *ADGRV1*, L) rs564080066 is located on *HTR4*, M) rs202007547 is located on *COL12A1*, N) rs542806512 is located on *NT5E*, O) rs576730428 is located on *GTF2H5*, P) rs561189374 is located on *ACTR3C*, Q) rs138507927 is located on *DPP6*, R) rs11136000 is located on *CLU*, S) rs192554851 is located on *ADAMTSL1*, T) rs570028361 is located on *PSAT1*, U) rs546018638 is located on *HPSE2*, V) rs535202922 is located on *ATP5L*, W) rs184836761 is located on *PIWIL1*, X) rs533330090 is located on *TBC1D4*, Y) rs576381927 is located on *ATP11A*, Z) rs527795127 is located on *TMX1*, AA) rs547455871 is located on *ATP10A*, AB) rs183472759 is located on *TMEM87A*, AC) rs146972069 is located on *ADAMTS17*, AD) rs564332482 is located on *CACNA1H*, AE) rs529505148 is located on *ITFG1*, AF) rs429358 is located on *APOE*, AG) rs78062743 is located on *BPIFA3*, AH) rs573127339 is located on *MROH8*, AI) rs569752694 is located on *CHD6*, AJ) rs575233877 is located on *PTPRT*, AK) rs529401448 is located on *TBC1D22A*.

**A.**

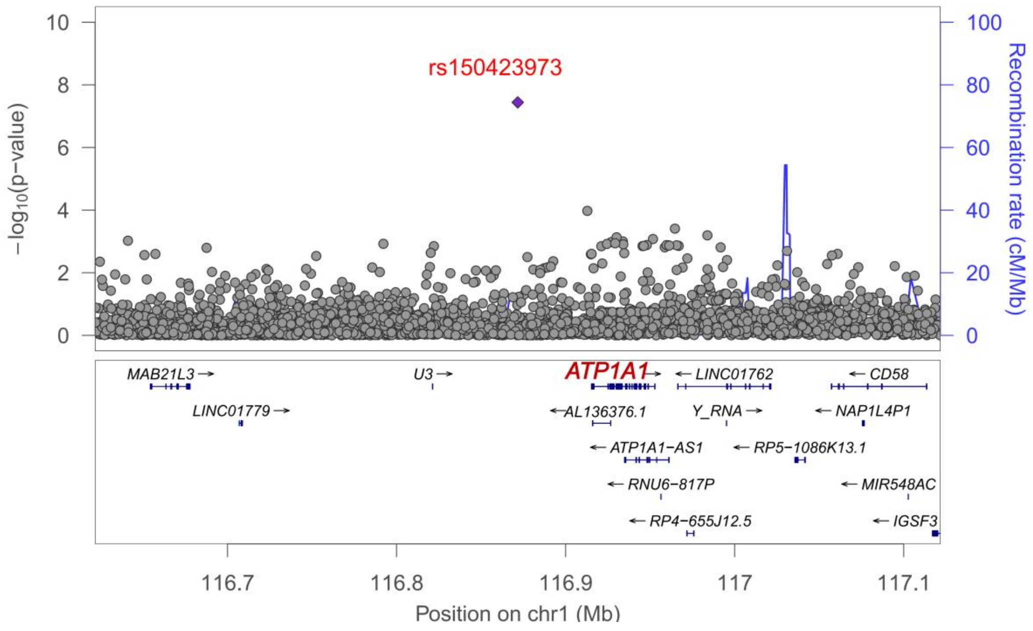

**B.**

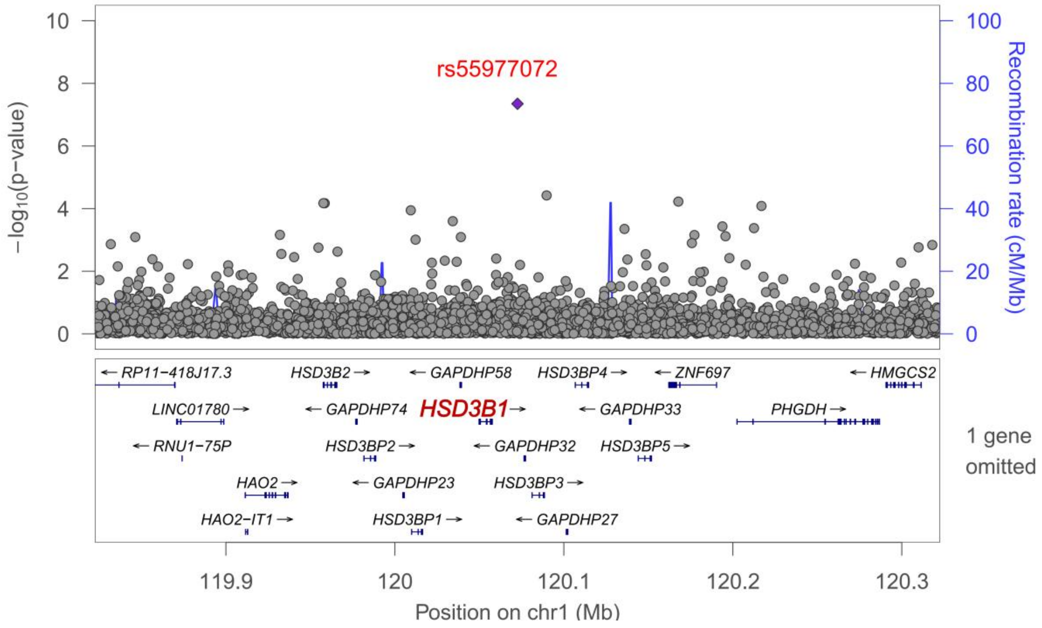
rs55977072
−log10(p−value)
Recombination rate (cM/Mb)
RP11−418J17.3
HSD3B2
GAPDHP58
HSD3BP4
ZNF697
HMGCS2
LINC01780
GAPDHP74
HSD3B1
GAPDHP33
PHGDH
RNU1−75P
HSD3BP2
GAPDHP32
HSD3BP5
HAO2
GAPDHP23
HSD3BP3
HAO2−IT1
HSD3BP1
GAPDHP27
1 gene omitted
119.9
120
120.1
120.2
120.3
Position on chr1 (Mb)

**C.**

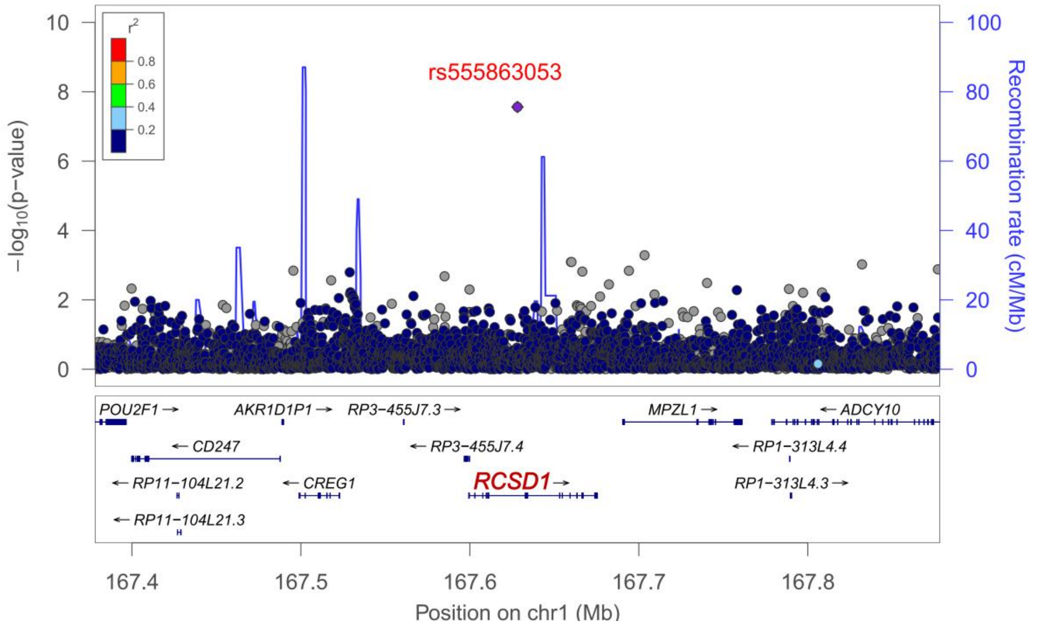
r2
0.8
0.6
0.4
0.2
rs555863053
−log10(p−value)
Recombination rate (cM/Mb)
POU2F1
AKR1D1P1
RP3−455J7.3
MPZL1
ADCY10
CD247
RP3−455J7.4
RP1−313L4.4
RP11−104L21.2
CREG1
RCSD1
RP1−313L4.3
RP11−104L21.3
167.4
167.5
167.6
167.7
167.8
Position on chr1 (Mb)

**D.**

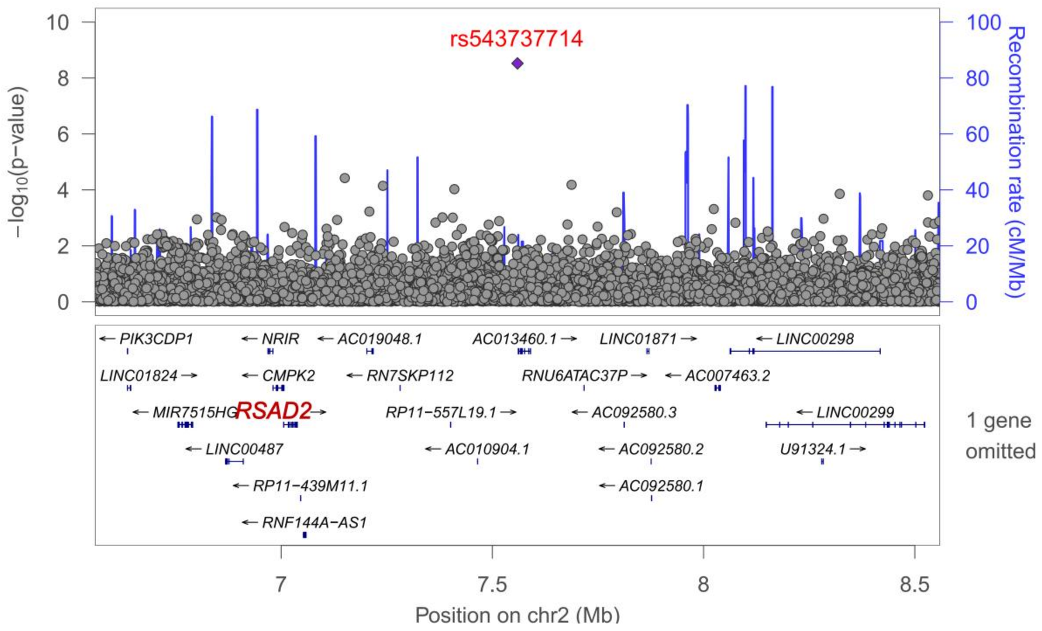

rs543737714
−log10(p−value)
Recombination rate (cM/Mb)
PIK3CDP1
NRIR
AC019048.1
AC013460.1
LINC01871
LINC00298
LINC01824
CMPK2
RN7SKP112
RNU6ATAC37P
AC007463.2
MIR7515HG
RSAD2
RP11−557L19.1
AC092580.3
LINC00299
LINC00487
AC010904.1
AC092580.2
U91324.1
RP11−439M11.1
AC092580.1
RNF144A−AS1
1 gene omitted
Position on chr2 (Mb)


**E.**

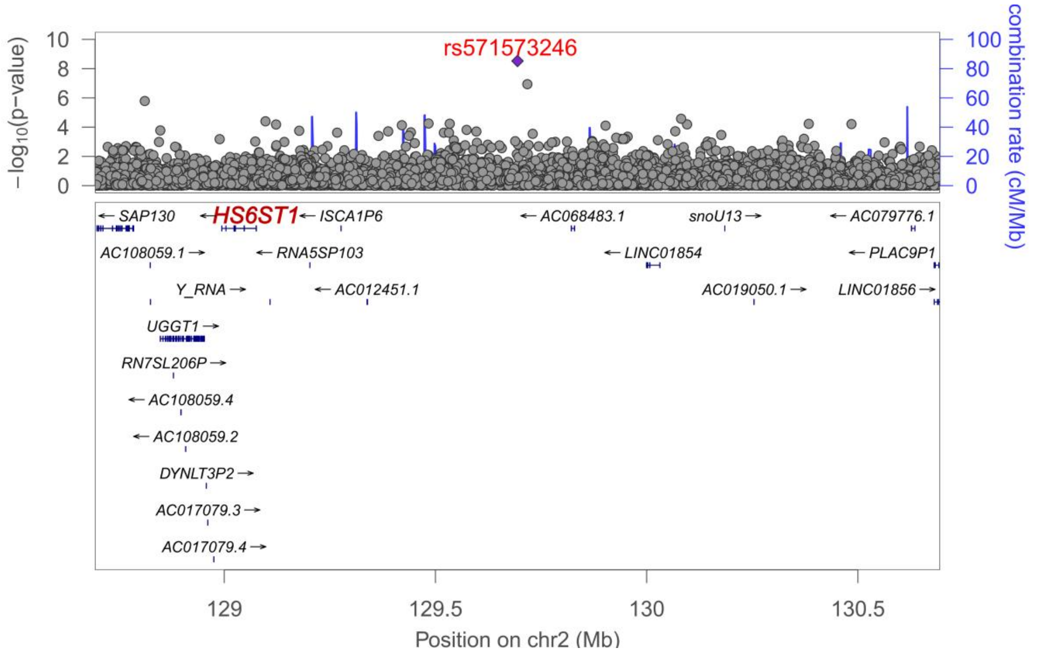

rs571573246
−log10(p−value)
combination rate (cM/Mb)
SAP130
HS6ST1
ISCA1P6
AC068483.1
snoU13
AC079776.1
AC108059.1
RNA5SP103
LINC01854
PLAC9P1
Y_RNA
AC012451.1
AC019050.1
LINC01856
UGGT1
RN7SL206P
AC108059.4
AC108059.2
DYNLT3P2
AC017079.3
AC017079.4
Position on chr2 (Mb)

**F.**

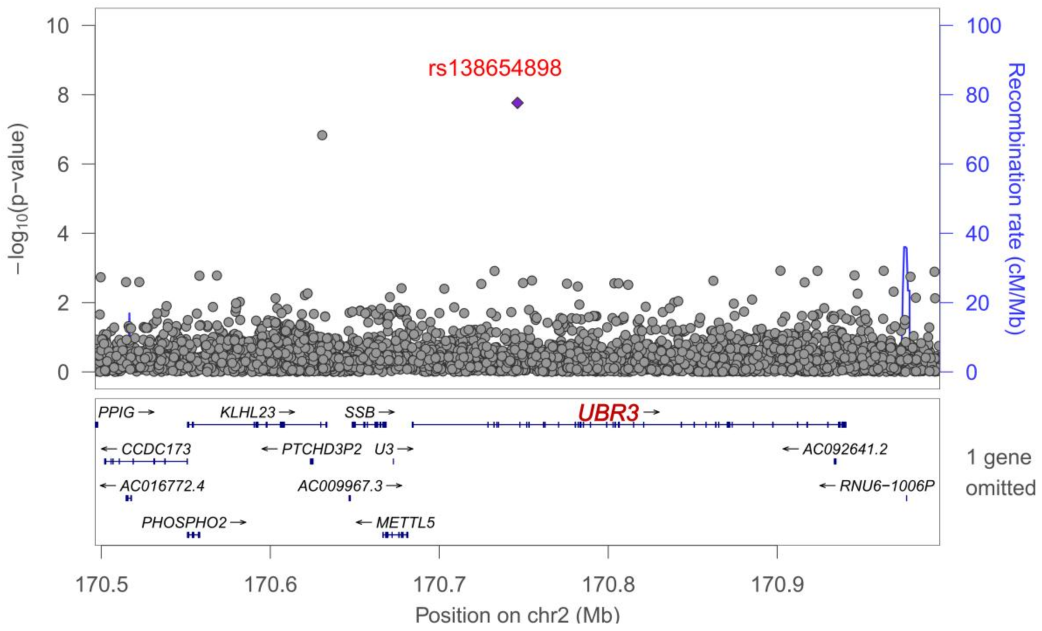

rs138654898
−log10(p−value)
Recombination rate (cM/Mb)
PPIG
KLHL23
SSB
UBR3
CCDC173
PTCHD3P2
U3
AC092641.2
AC016772.4
AC009967.3
RNU6−1006P
PHOSPHO2
METTL5
1 gene omitted
170.5
170.6
170.7
170.8
170.9
Position on chr2 (Mb)


**G.**

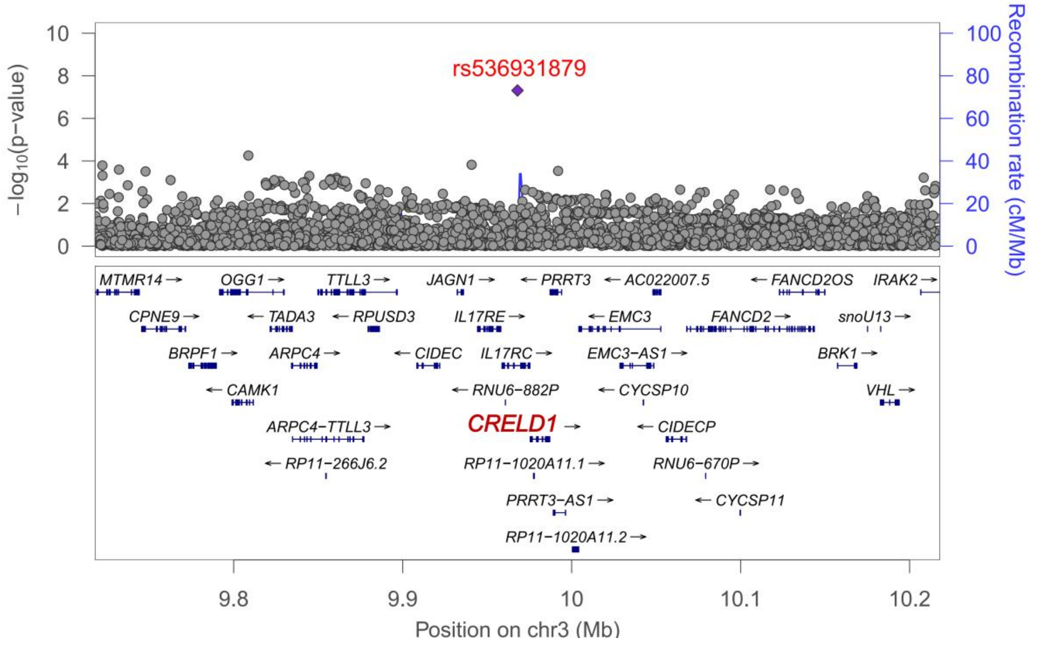

rs536931879
−log10(p−value)
Recombination rate (cM/Mb)
MTMR14
OGG1
TTLL3
JAGN1
PRRT3
AC022007.5
FANCD2OS
IRAK2
CPNE9
TADA3
RPUSD3
IL17RE
EMC3
FANCD2
snoU13
BRPF1
ARPC4
CIDEC
IL17RC
EMC3−AS1
BRK1
CAMK1
RNU6−882P
CYCSP10
VHL
ARPC4−TTLL3
CRELD1
CIDECP
RP11−266J6.2
RP11−1020A11.1
RNU6−670P
PRRT3−AS1
CYCSP11
RP11−1020A11.2
9.8
9.9
10
10.1
10.2
Position on chr3 (Mb)

**H.**

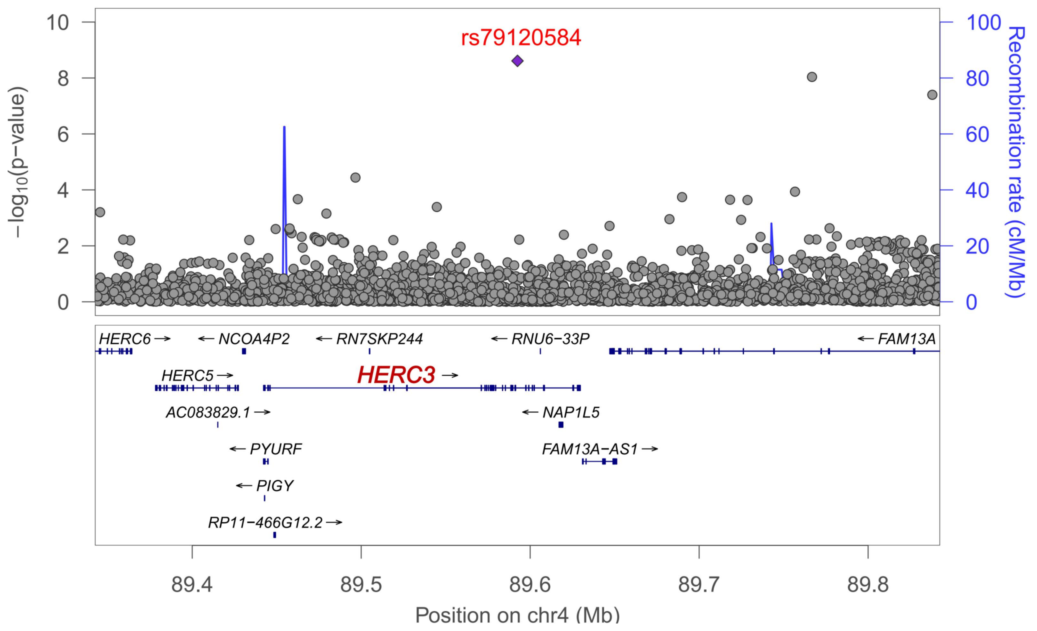

rs79120584
−log10(p−value)
Recombination rate (cM/Mb)
HERC6 →
← NCOA4P2
← RN7SKP244
← RNU6−33P
← FAM13A
HERC5 →
HERC3 →
AC083829.1 →
← NAP1L5
← PYURF
FAM13A−AS1 →
← PIGY
RP11−466G12.2 →
89.4
89.5
89.6
89.7
89.8
Position on chr4 (Mb)


**I.**

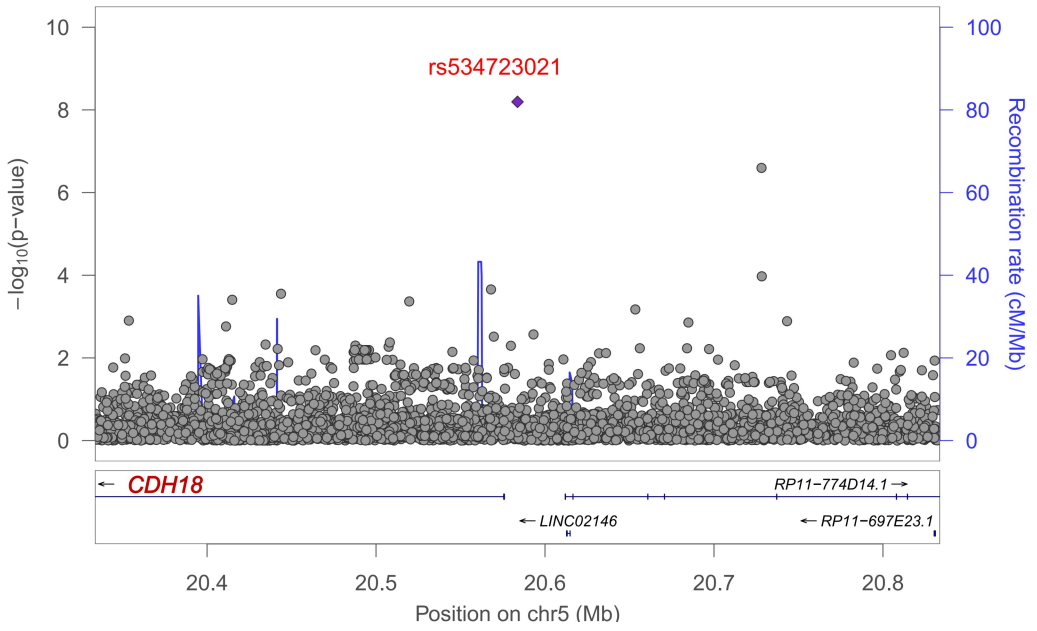

rs534723021
−log10(p−value)
Recombination rate (cM/Mb)
← CDH18
RP11−774D14.1 →
← LINC02146
← RP11−697E23.1
20.4
20.5
20.6
20.7
20.8
Position on chr5 (Mb)

J.

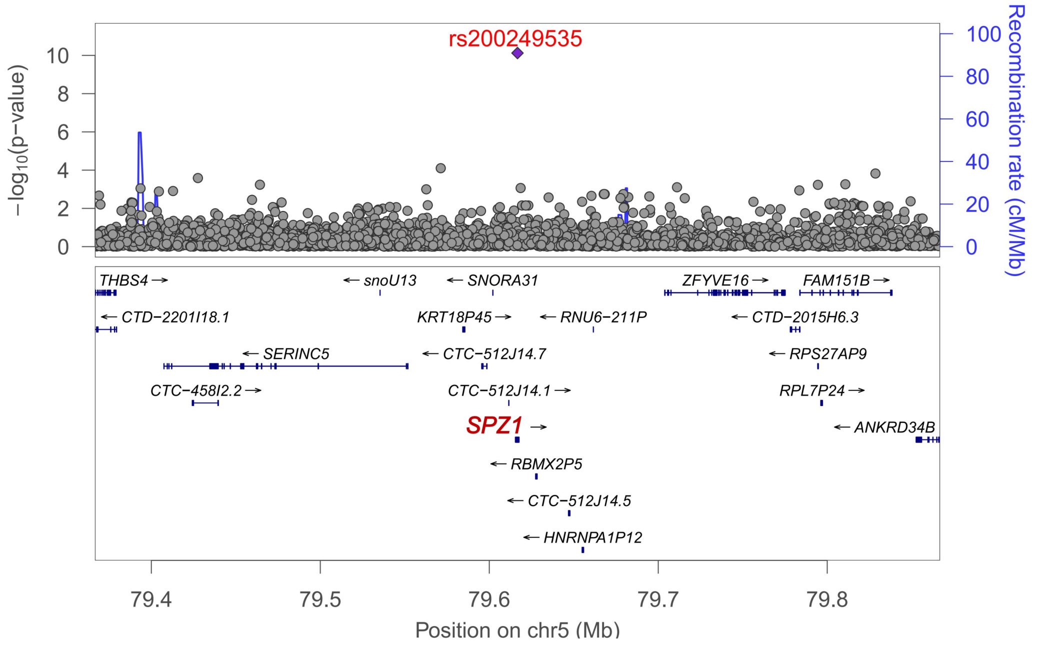
rs200249535
−log10(p−value)
Recombination rate (cM/Mb)
THBS4 →
← snoU13
← SNORA31
ZFYVE16 →
FAM151B →
← CTD−2201I18.1
KRT18P45 →
← RNU6−211P
← CTD−2015H6.3
← SERINC5
← CTC−512J14.7
← RPS27AP9
CTC−458I2.2 →
CTC−512J14.1 →
RPL7P24 →
SPZ1 →
← ANKRD34B
← RBMX2P5
← CTC−512J14.5
← HNRNPA1P12
Position on chr5 (Mb)

K

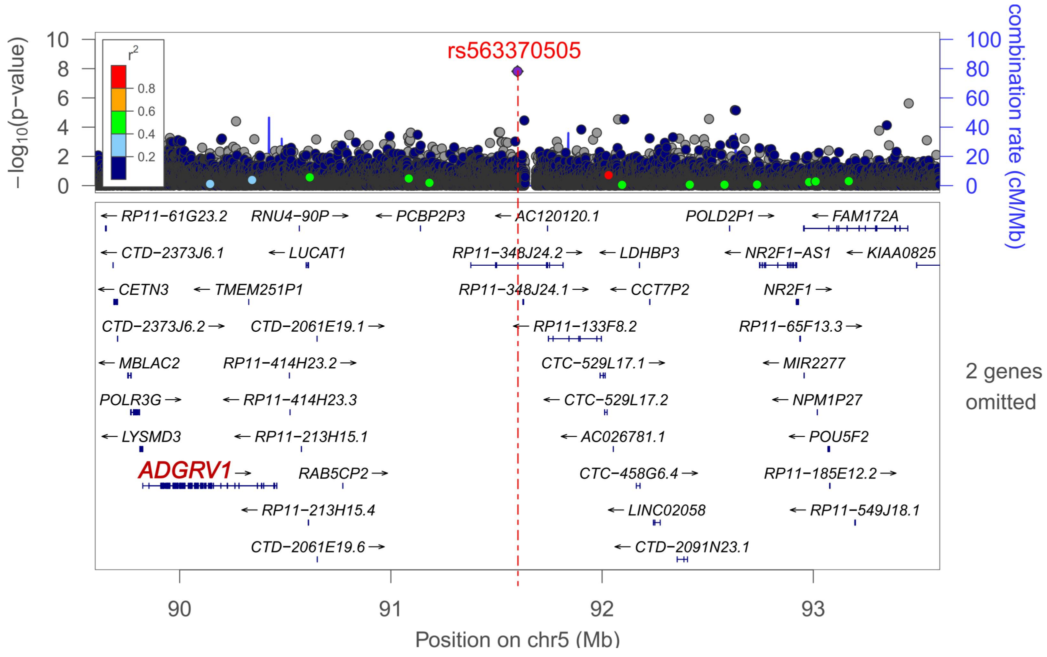
rs563370505
−log10(p−value)
combination rate (cM/Mb)
r2
← RP11−61G23.2
RNU4−90P →
← PCBP2P3
← AC120120.1
POLD2P1 →
← FAM172A
← CTD−2373J6.1
← LUCAT1
RP11−348J24.2 →
← LDHBP3
← NR2F1−AS1
← KIAA0825
← CETN3
← TMEM251P1
RP11−348J24.1 →
← CCT7P2
NR2F1 →
CTD−2373J6.2 →
CTD−2061E19.1 →
← RP11−133F8.2
RP11−65F13.3 →
← MBLAC2
RP11−414H23.2 →
CTC−529L17.1 →
← MIR2277
POLR3G →
← RP11−414H23.3
← CTC−529L17.2
← NPM1P27
← LYSMD3
← RP11−213H15.1
← AC026781.1
← POU5F2
ADGRV1 →
RAB5CP2 →
CTC−458G6.4 →
RP11−185E12.2 →
← RP11−213H15.4
← LINC02058
← RP11−549J18.1
CTD−2061E19.6 →
← CTD−2091N23.1
2 genes omitted
Position on chr5 (Mb)

**L.**

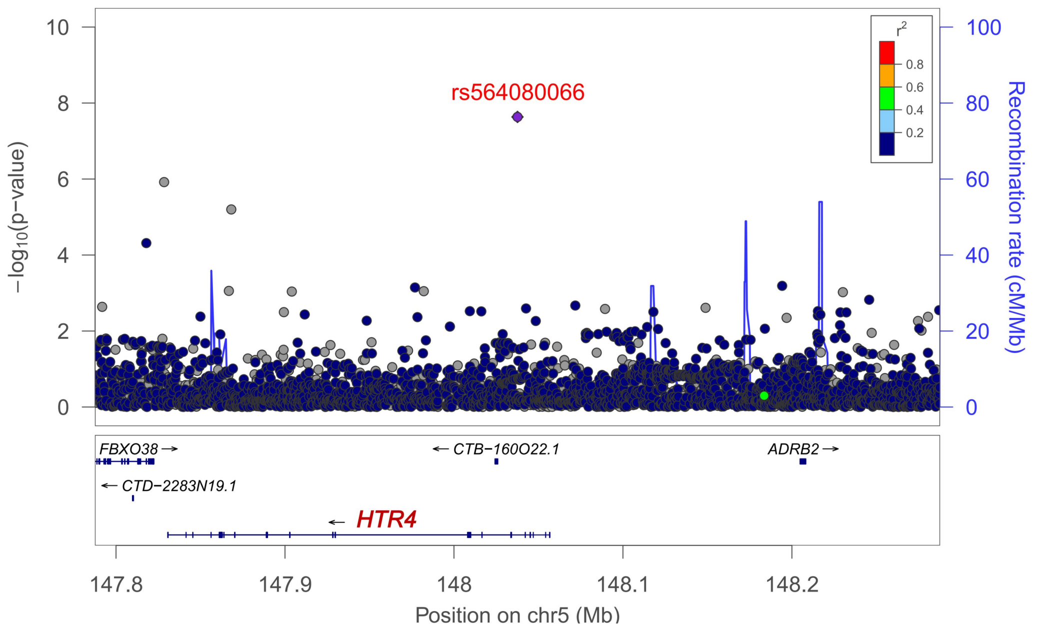

rs564080066
−log10(p−value)
Recombination rate (cM/Mb)
r2
FBXO38
CTD−2283N19.1
CTB−160O22.1
HTR4
ADRB2
147.8
147.9
148
148.1
148.2
Position on chr5 (Mb)


.

**M.**

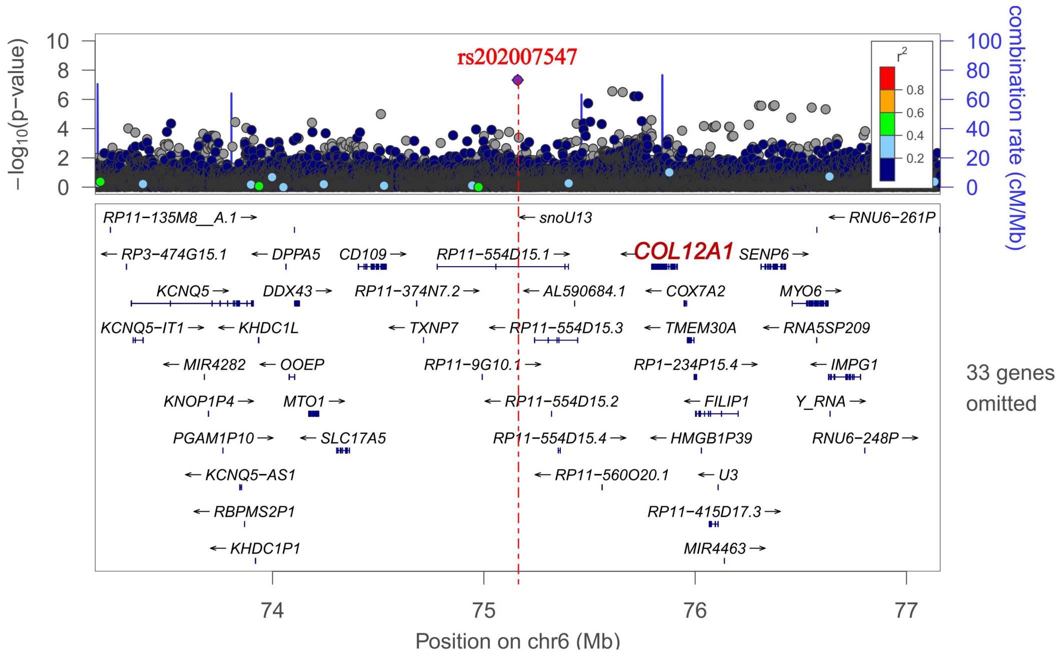

rs202007547
−log10(p−value)
combination rate (cM/Mb)
r2
COL12A1
33 genes omitted
74
75
76
77
Position on chr6 (Mb)

**N.**

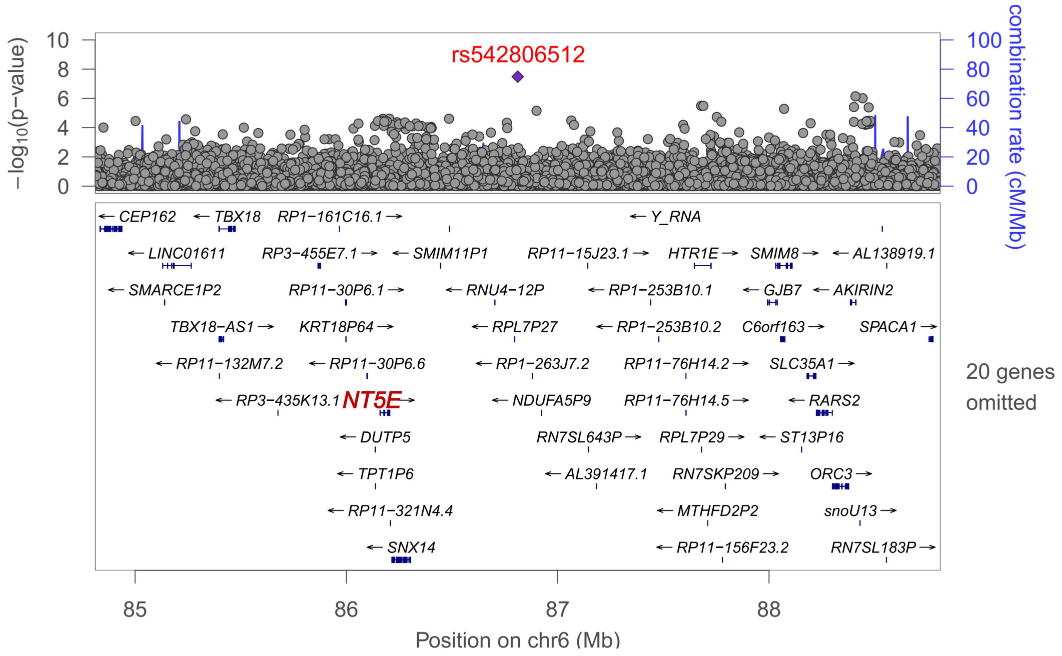
rs542806512
−log10(p−value)
combination rate (cM/Mb)
CEP162
TBX18
RP1−161C16.1
Y_RNA
LINC01611
RP3−455E7.1
SMIM11P1
RP11−15J23.1
HTR1E
SMIM8
AL138919.1
SMARCE1P2
RP11−30P6.1
RNU4−12P
RP1−253B10.1
GJB7
AKIRIN2
TBX18−AS1
KRT18P64
RPL7P27
RP1−253B10.2
C6orf163
SPACA1
RP11−132M7.2
RP11−30P6.6
RP1−263J7.2
RP11−76H14.2
SLC35A1
20 genes omitted
RP3−435K13.1
NT5E
NDUFA5P9
RP11−76H14.5
RARS2
DUTP5
RN7SL643P
RPL7P29
ST13P16
TPT1P6
AL391417.1
RN7SKP209
ORC3
RP11−321N4.4
MTHFD2P2
snoU13
SNX14
RP11−156F23.2
RN7SL183P
Position on chr6 (Mb)

**O.**

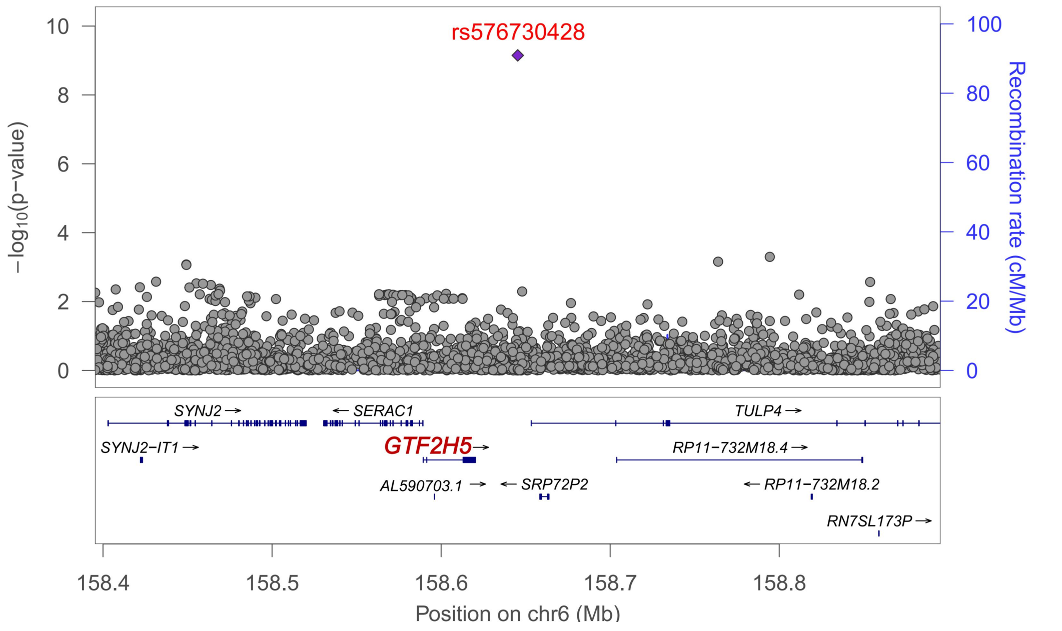
rs576730428
−log10(p−value)
Recombination rate (cM/Mb)
SYNJ2
SERAC1
TULP4
SYNJ2−IT1
GTF2H5
RP11−732M18.4
AL590703.1
SRP72P2
RP11−732M18.2
RN7SL173P
Position on chr6 (Mb)

**P.**

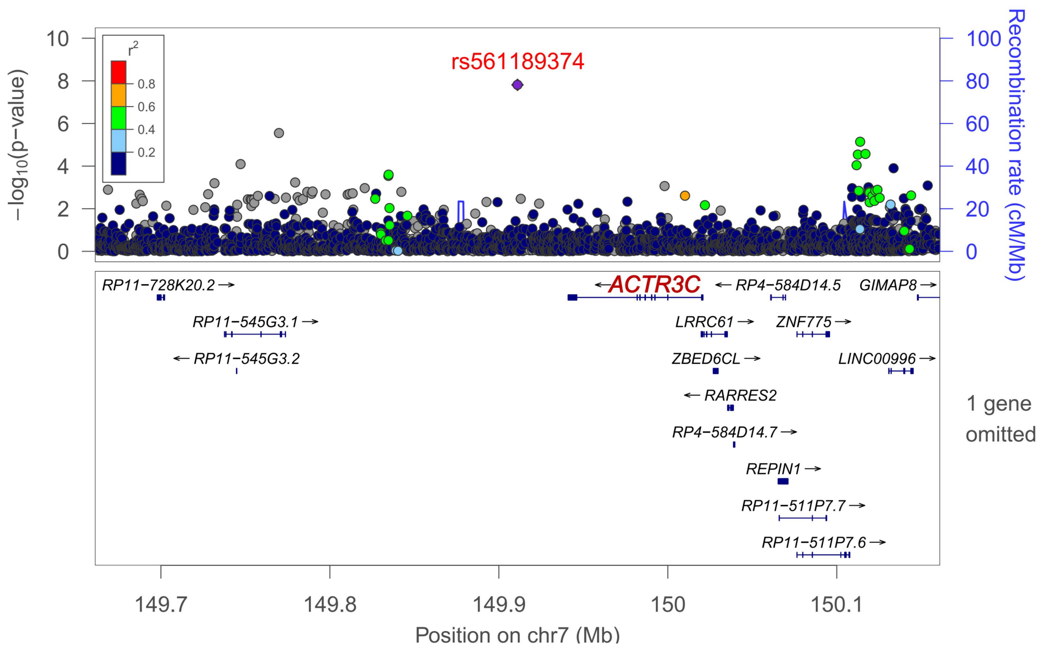
rs561189374
r2
0.8
0.6
0.4
0.2
−log10(p−value)
Recombination rate (cM/Mb)
RP11−728K20.2
ACTR3C
RP4−584D14.5
GIMAP8
RP11−545G3.1
LRRC61
ZNF775
RP11−545G3.2
ZBED6CL
LINC00996
RARRES2
RP4−584D14.7
REPIN1
RP11−511P7.7
RP11−511P7.6
1 gene omitted
149.7
149.8
149.9
150
150.1
Position on chr7 (Mb)

**Q.**

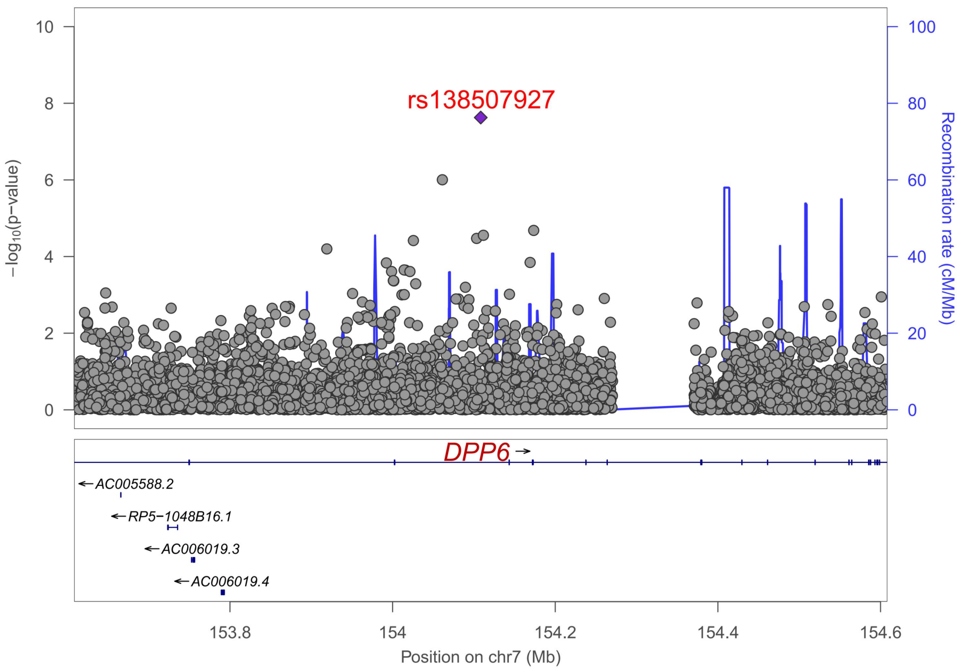
rs138507927
−log10(p−value)
Recombination rate (cM/Mb)
DPP6
AC005588.2
RP5−1048B16.1
AC006019.3
AC006019.4
153.8
154
154.2
154.4
154.6
Position on chr7 (Mb)

**R.**

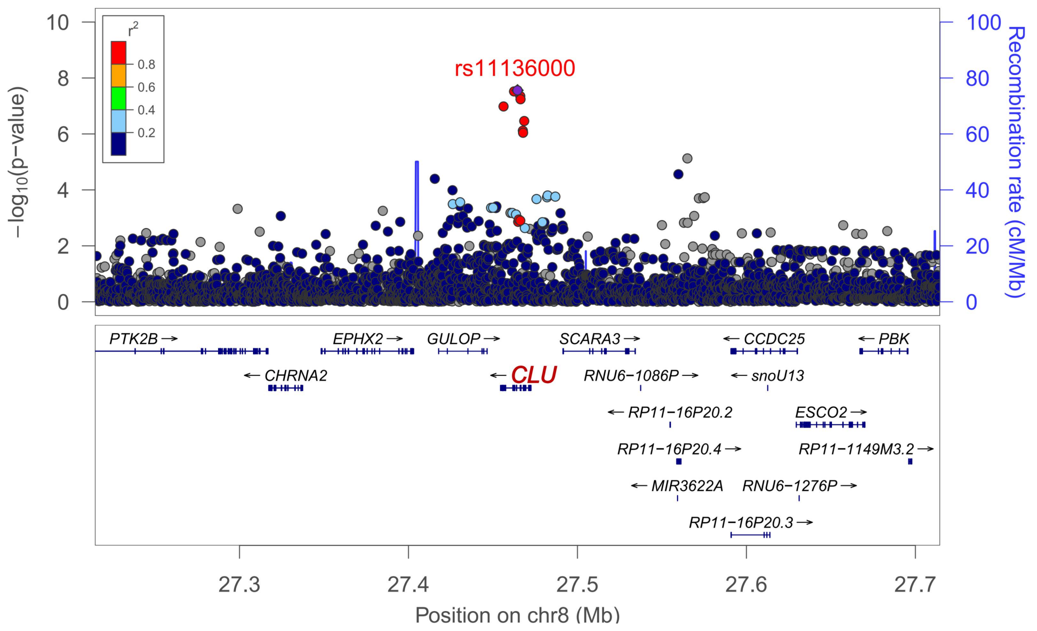

rs11136000
r²
0.8
0.6
0.4
0.2
−log10(p−value)
Recombination rate (cM/Mb)
PTK2B
CHRNA2
EPHX2
GULOP
CLU
SCARA3
RNU6−1086P
RP11−16P20.2
RP11−16P20.4
MIR3622A
RP11−16P20.3
CCDC25
snoU13
RNU6−1276P
ESCO2
RP11−1149M3.2
PBK
27.3
27.4
27.5
27.6
27.7
Position on chr8 (Mb)


**S.**

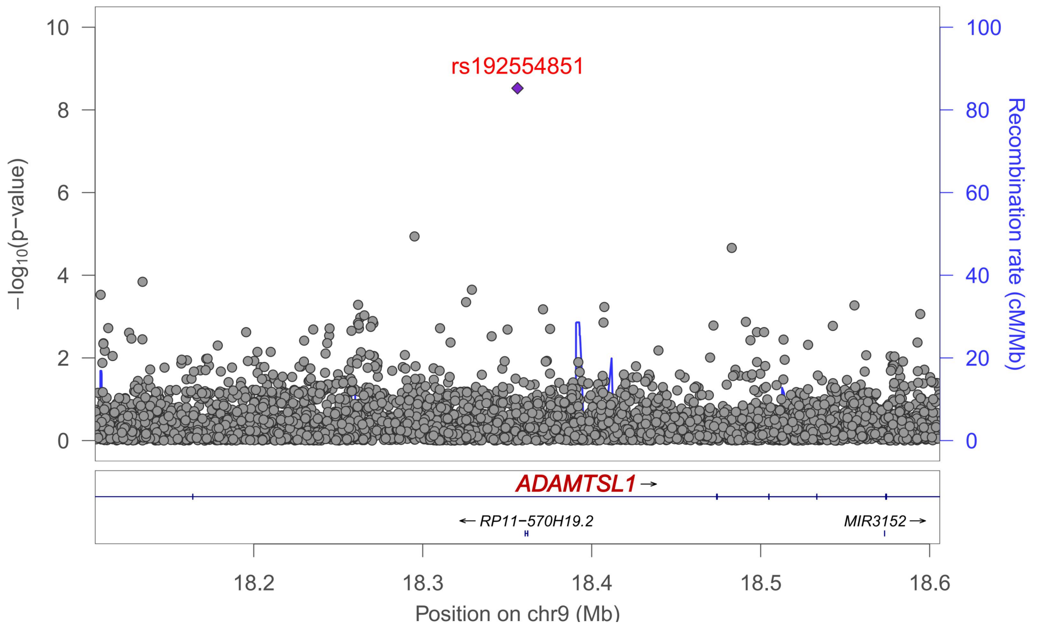

rs192554851
−log10(p−value)
Recombination rate (cM/Mb)
ADAMTSL1
RP11−570H19.2
MIR3152
18.2
18.3
18.4
18.5
18.6
Position on chr9 (Mb)

**T.**

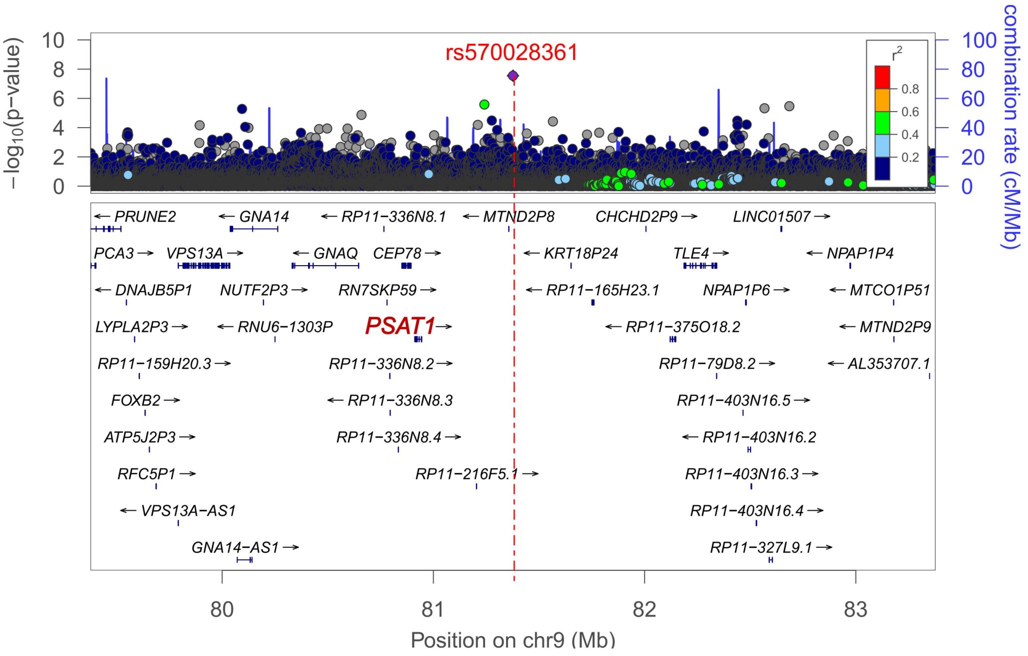

rs570028361
−log10(p−value)
Recombination rate (cM/Mb)
r2
PRUNE2
GNA14
RP11−336N8.1
MTND2P8
CHCHD2P9
LINC01507
PCA3
VPS13A
GNAQ
CEP78
KRT18P24
TLE4
NPAP1P4
DNAJB5P1
NUTF2P3
RN7SKP59
RP11−165H23.1
NPAP1P6
MTCO1P51
LYPLA2P3
RNU6−1303P
PSAT1
RP11−375O18.2
MTND2P9
RP11−159H20.3
RP11−336N8.2
RP11−79D8.2
AL353707.1
FOXB2
RP11−336N8.3
RP11−403N16.5
ATP5J2P3
RP11−336N8.4
RP11−403N16.2
RFC5P1
RP11−216F5.1
RP11−403N16.3
VPS13A−AS1
RP11−403N16.4
GNA14−AS1
RP11−327L9.1
Position on chr9 (Mb)


**U.**

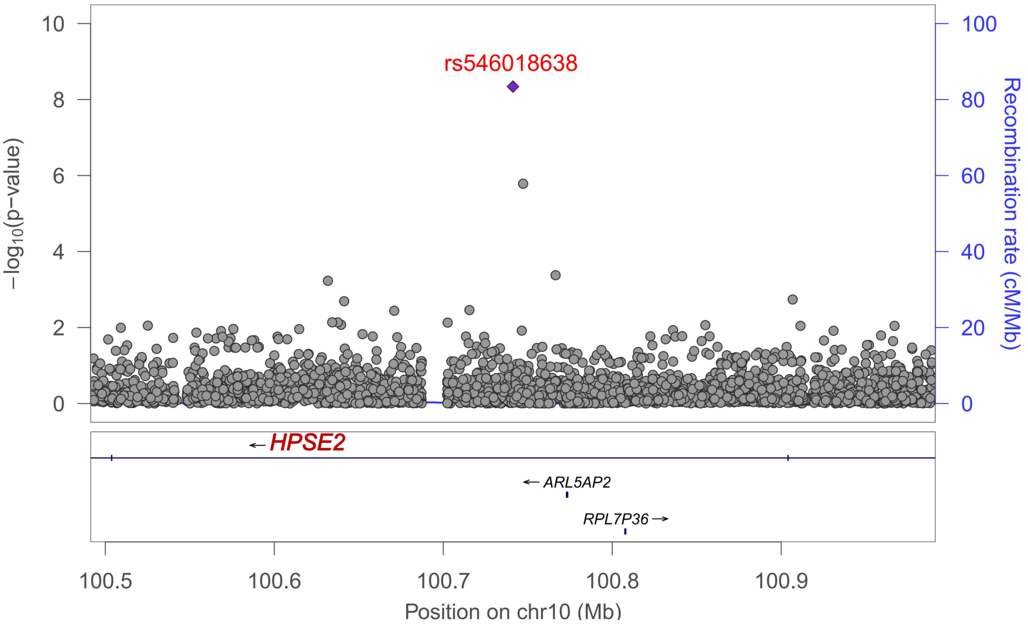

rs546018638
−log10(p−value)
Recombination rate (cM/Mb)
HPSE2
ARL5AP2
RPL7P36
Position on chr10 (Mb)

**V.**

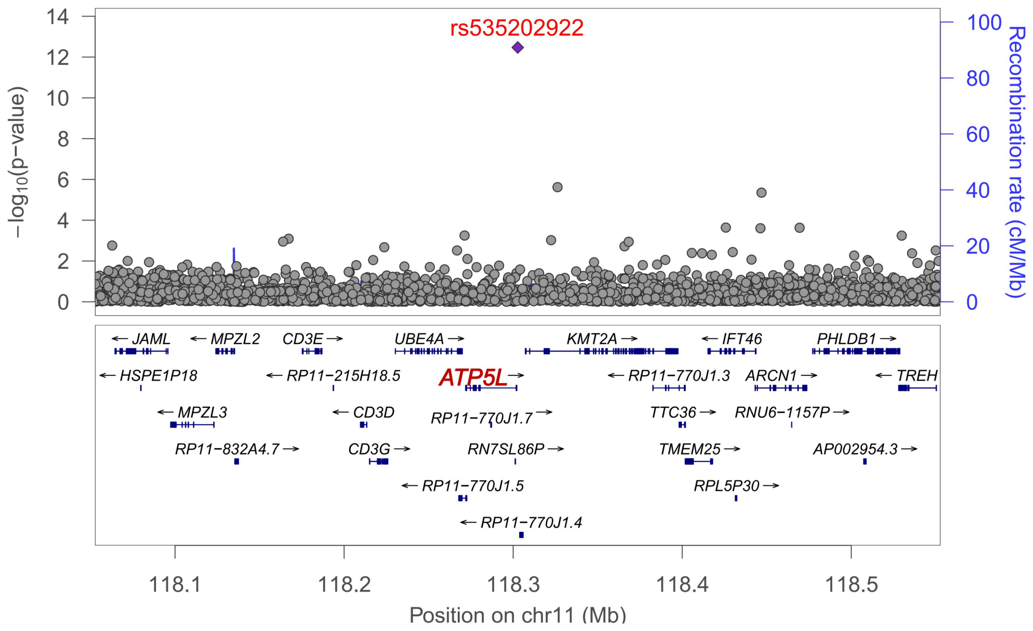
rs535202922
14
12
10
8
6
4
2
0
−log10(p−value)
100
80
60
40
20
0
Recombination rate (cM/Mb)
JAML
MPZL2
CD3E
UBE4A
KMT2A
IFT46
PHLDB1
HSPE1P18
RP11−215H18.5
ATP5L
RP11−770J1.3
ARCN1
TREH
MPZL3
CD3D
RP11−770J1.7
TTC36
RNU6−1157P
RP11−832A4.7
CD3G
RN7SL86P
TMEM25
AP002954.3
RP11−770J1.5
RPL5P30
RP11−770J1.4
118.1
118.2
118.3
118.4
118.5
Position on chr11 (Mb)

**W.**

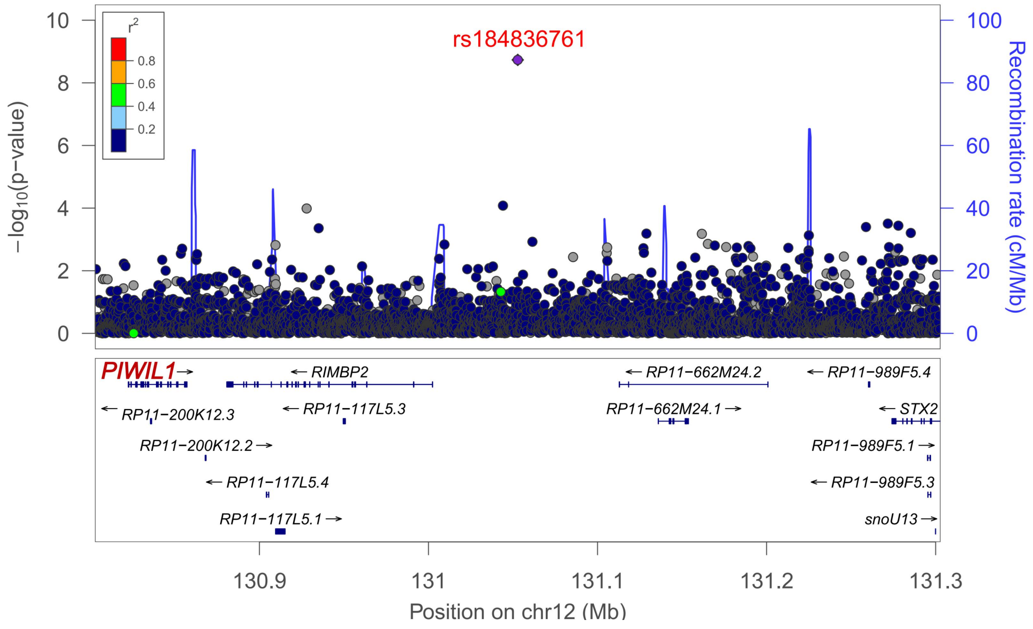
rs184836761
r2
0.8
0.6
0.4
0.2
10
8
6
4
2
0
−log10(p−value)
100
80
60
40
20
0
Recombination rate (cM/Mb)
PIWIL1
RIMBP2
RP11−662M24.2
RP11−989F5.4
RP11−200K12.3
RP11−117L5.3
RP11−662M24.1
STX2
RP11−200K12.2
RP11−989F5.1
RP11−117L5.4
RP11−989F5.3
RP11−117L5.1
snoU13
130.9
131
131.1
131.2
131.3
Position on chr12 (Mb)

**X.**

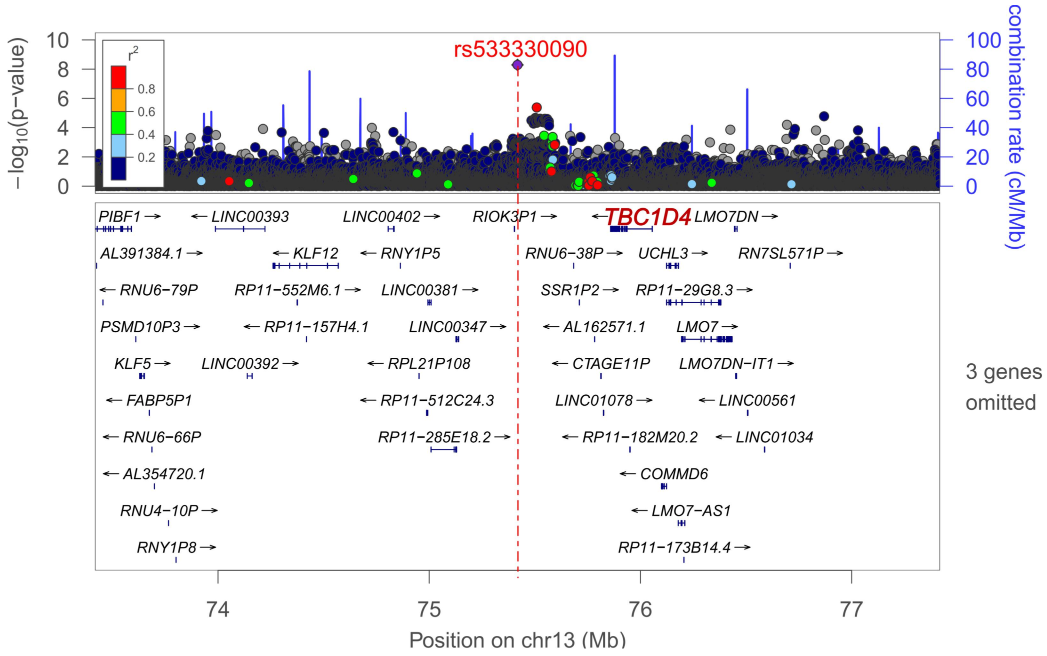

rs533330090
−log10(p-value)
combination rate (cM/Mb)
r2
0.8
0.6
0.4
0.2
PIBF1 LINC00393 LINC00402 RIOK3P1 TBC1D4 LMO7DN
AL391384.1 KLF12 RNY1P5 RNU6−38P UCHL3 RN7SL571P
RNU6−79P RP11−552M6.1 LINC00381 SSR1P2 RP11−29G8.3
PSMD10P3 RP11−157H4.1 LINC00347 AL162571.1 LMO7
KLF5 LINC00392 RPL21P108 CTAGE11P LMO7DN−IT1
FABP5P1 RP11−512C24.3 LINC01078 LINC00561
RNU6−66P RP11−285E18.2 RP11−182M20.2 LINC01034
AL354720.1 COMMD6
RNU4−10P LMO7−AS1
RNY1P8 RP11−173B14.4
3 genes omitted
Position on chr13 (Mb)


**Y.**

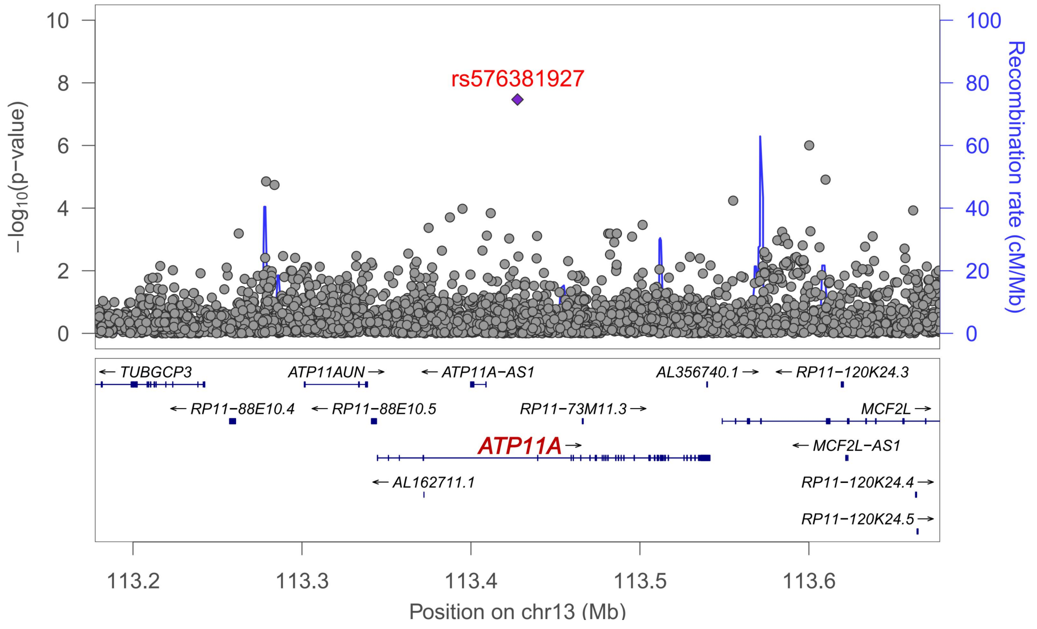

rs576381927
−log10(p-value)
Recombination rate (cM/Mb)
TUBGCP3 ATP11AUN ATP11A−AS1 AL356740.1 RP11−120K24.3
RP11−88E10.4 RP11−88E10.5 RP11−73M11.3 MCF2L
ATP11A MCF2L−AS1
AL162711.1 RP11−120K24.4
RP11−120K24.5
Position on chr13 (Mb)

**Z.**

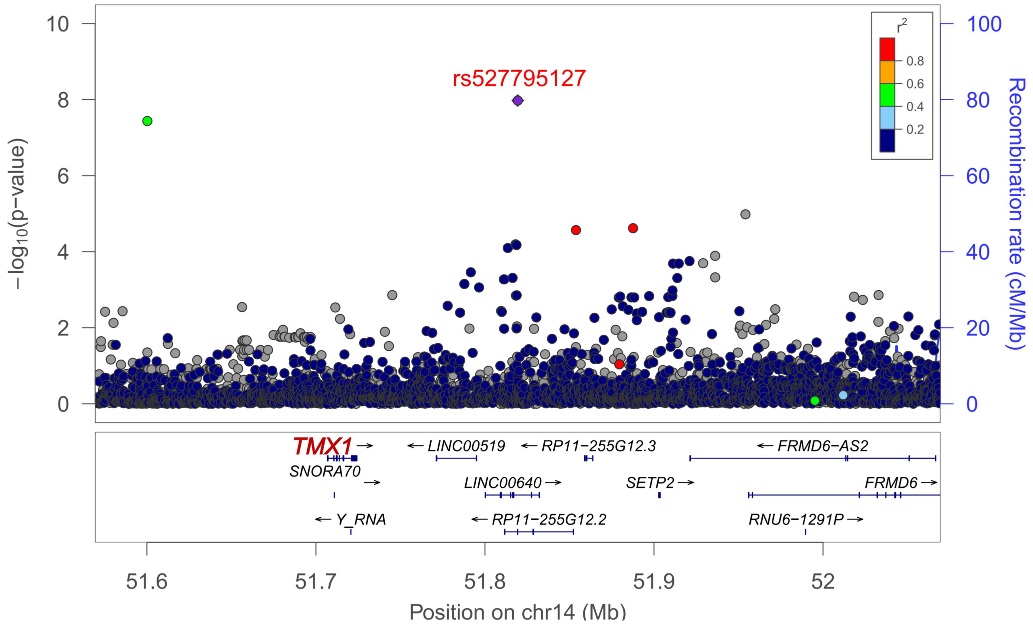
rs527795127
−log10(p−value)
Recombination rate (cM/Mb)
r2
0.8
0.6
0.4
0.2
TMX1
SNORA70
Y_RNA
LINC00519
LINC00640
RP11−255G12.2
RP11−255G12.3
SETP2
FRMD6−AS2
FRMD6
RNU6−1291P
51.6
51.7
51.8
51.9
52
Position on chr14 (Mb)

**AA.**

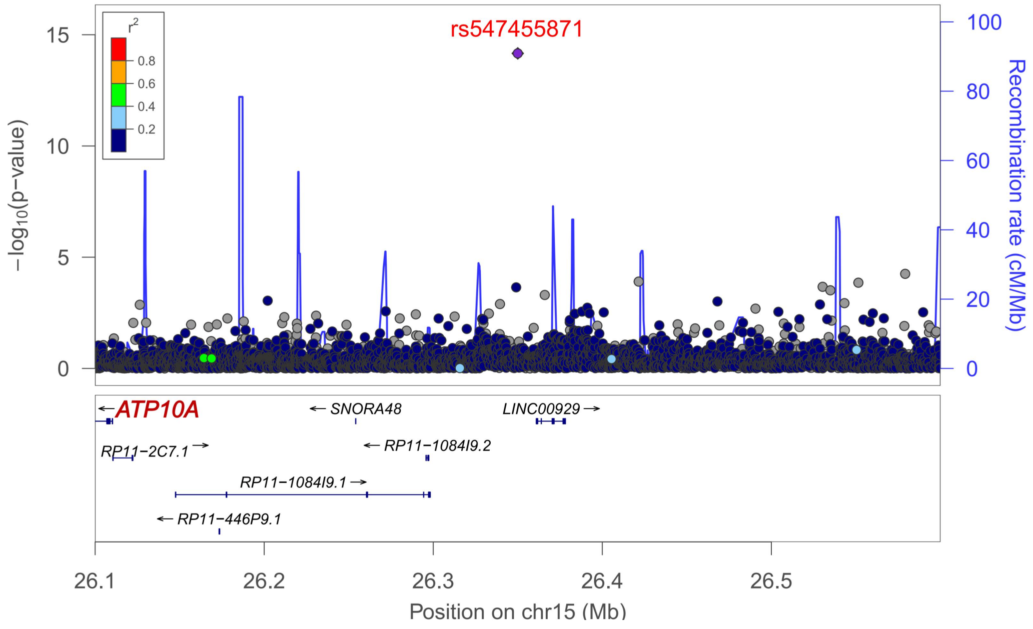
rs547455871
−log10(p−value)
Recombination rate (cM/Mb)
r2
0.8
0.6
0.4
0.2
ATP10A
RP11−2C7.1
RP11−1084I9.1
RP11−446P9.1
SNORA48
RP11−1084I9.2
LINC00929
26.1
26.2
26.3
26.4
26.5
Position on chr15 (Mb)

AB.

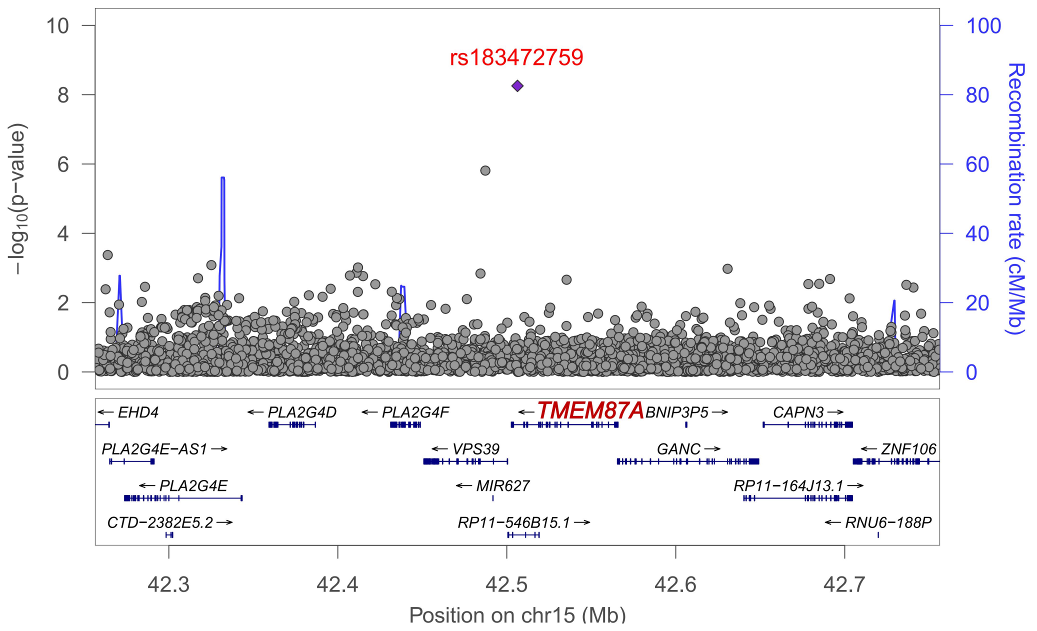
rs183472759
−log10(p−value)
Recombination rate (cM/Mb)
EHD4
PLA2G4D
PLA2G4F
TMEM87A
BNIP3P5
CAPN3
PLA2G4E−AS1
VPS39
GANC
ZNF106
PLA2G4E
MIR627
RP11−164J13.1
CTD−2382E5.2
RP11−546B15.1
RNU6−188P
42.3
42.4
42.5
42.6
42.7
Position on chr15 (Mb)

AC.

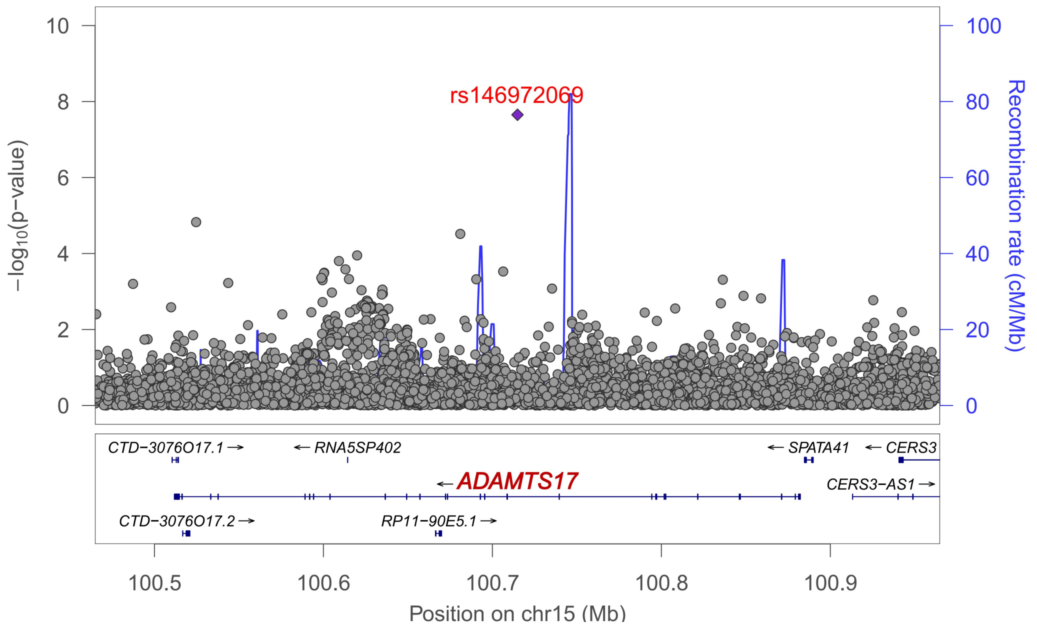
rs146972069
−log10(p−value)
Recombination rate (cM/Mb)
CTD−3076O17.1
RNA5SP402
SPATA41
CERS3
ADAMTS17
CERS3−AS1
CTD−3076O17.2
RP11−90E5.1
100.5
100.6
100.7
100.8
100.9
Position on chr15 (Mb)

**AD.**

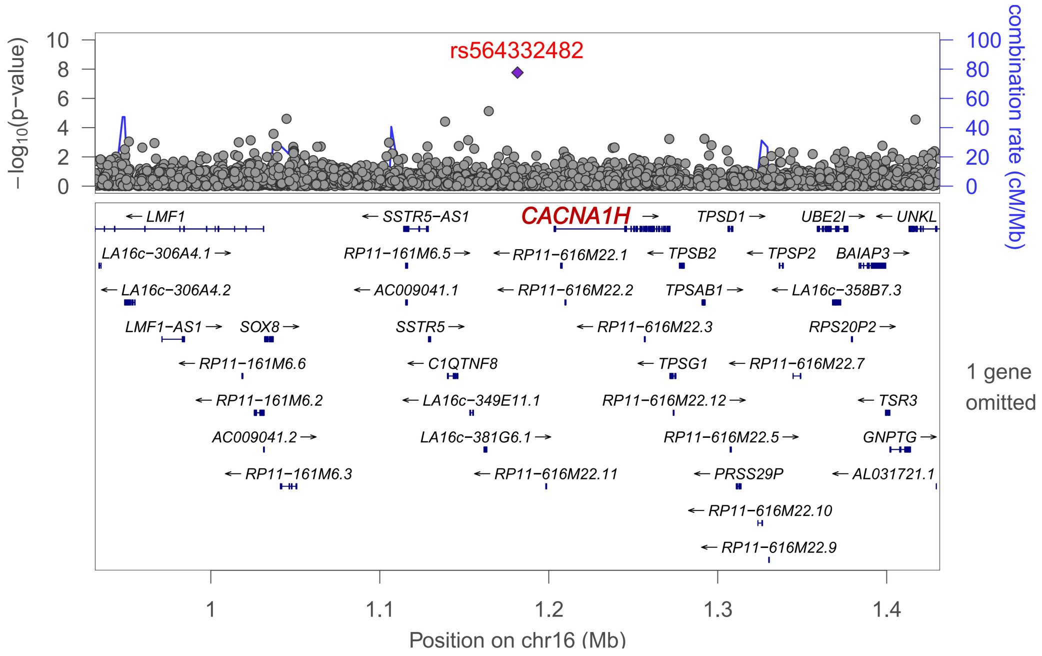
rs564332482
CACNA1H
−log10(p−value)
combination rate (cM/Mb)
1 gene omitted
Position on chr16 (Mb)

**AE.**

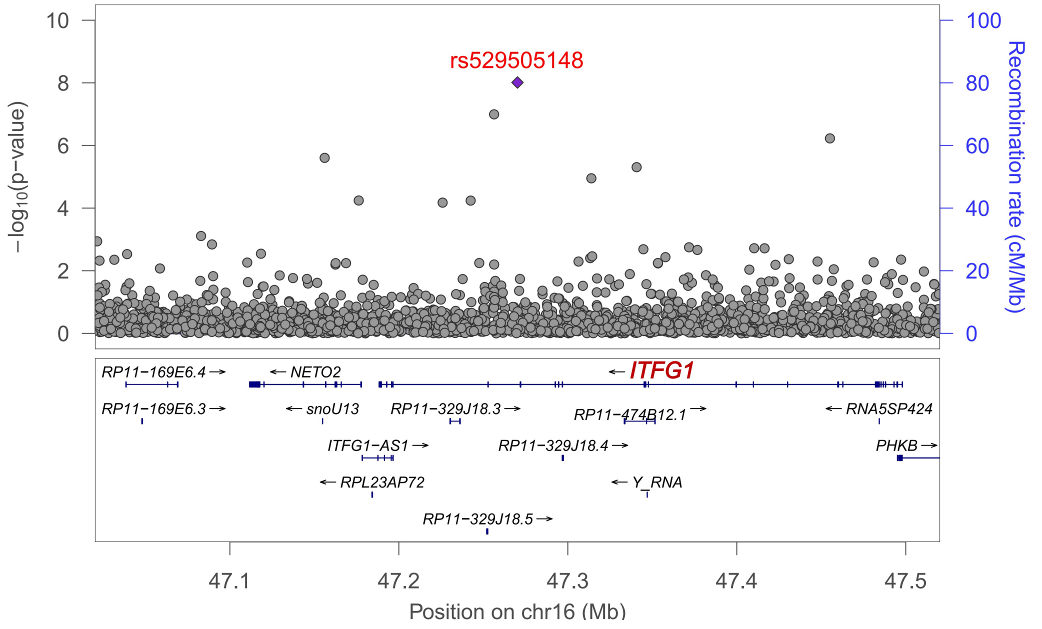
rs529505148
ITFG1
−log10(p−value)
Recombination rate (cM/Mb)
Position on chr16 (Mb)

**AF.**

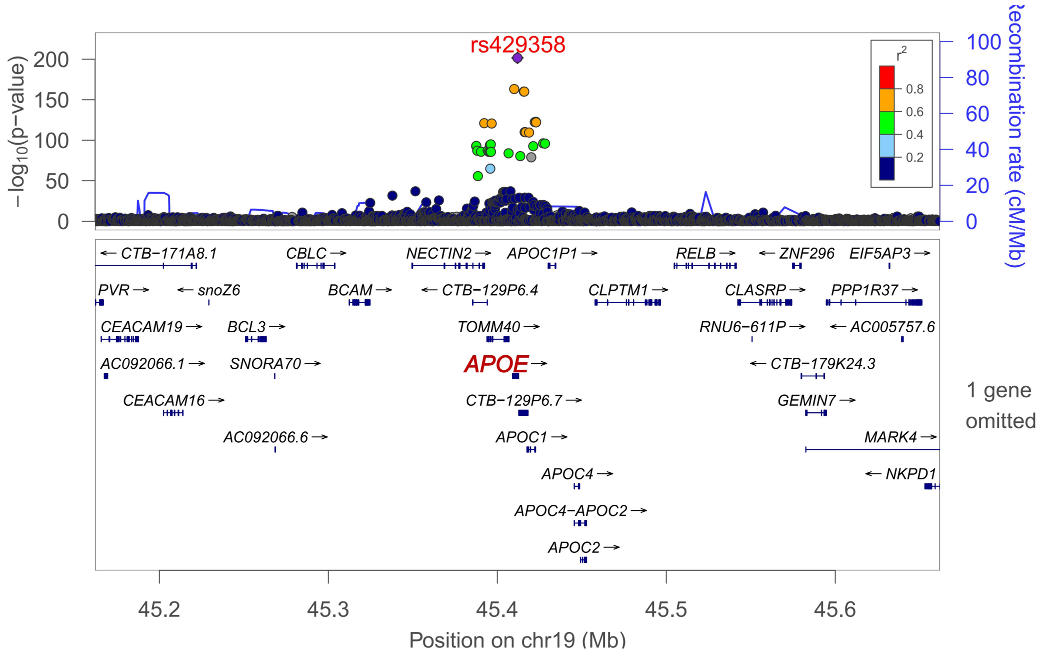
rs429358
−log10(p−value)
Recombination rate (cM/Mb)
r2
APOE
TOMM40
APOC1
APOC4
APOC2
1 gene omitted
Position on chr19 (Mb)

**AG.**

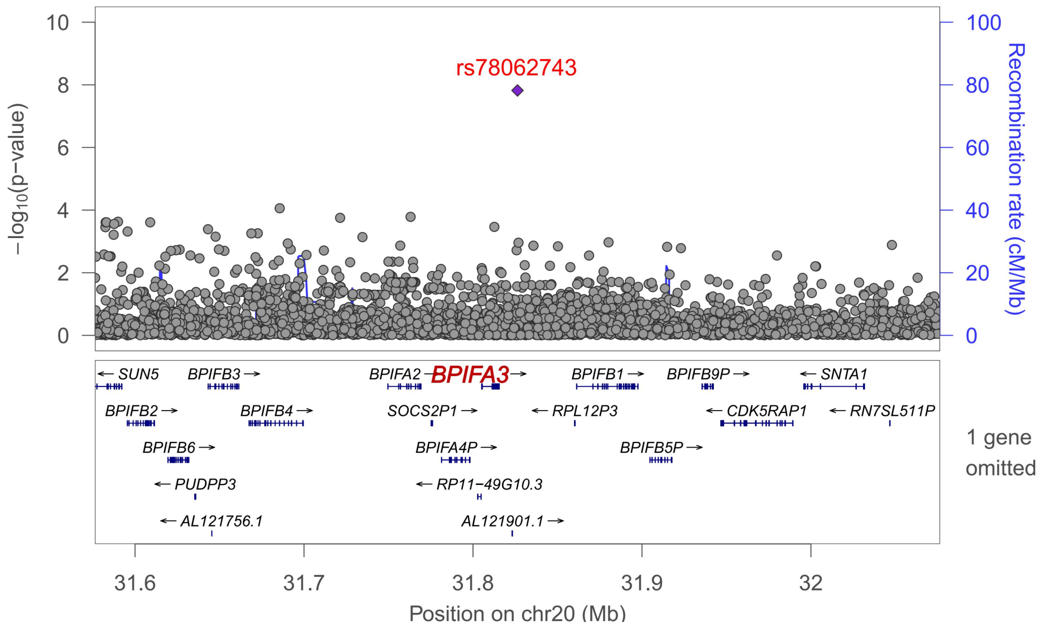
rs78062743
−log10(p−value)
Recombination rate (cM/Mb)
BPIFA3
BPIFA2
BPIFB1
1 gene omitted
Position on chr20 (Mb)

**AH.**

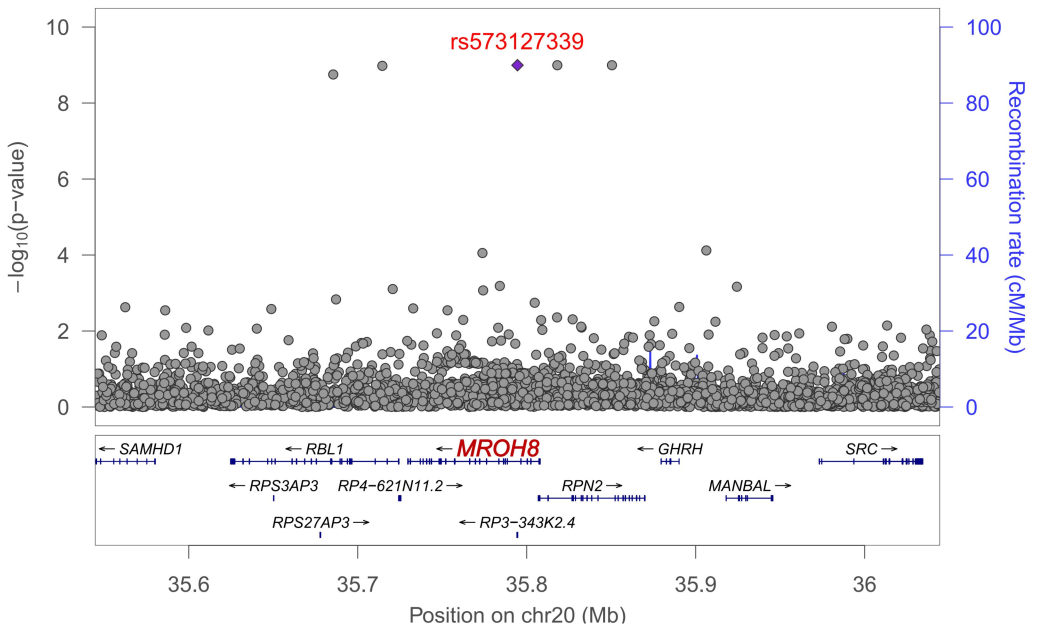

rs573127339
−log10(p−value)
Recombination rate (cM/Mb)
SAMHD1
RBL1
MROH8
GHRH
SRC
RPS3AP3
RP4−621N11.2
RPN2
MANBAL
RPS27AP3
RP3−343K2.4
35.6
35.7
35.8
35.9
36
Position on chr20 (Mb)


**AI.**

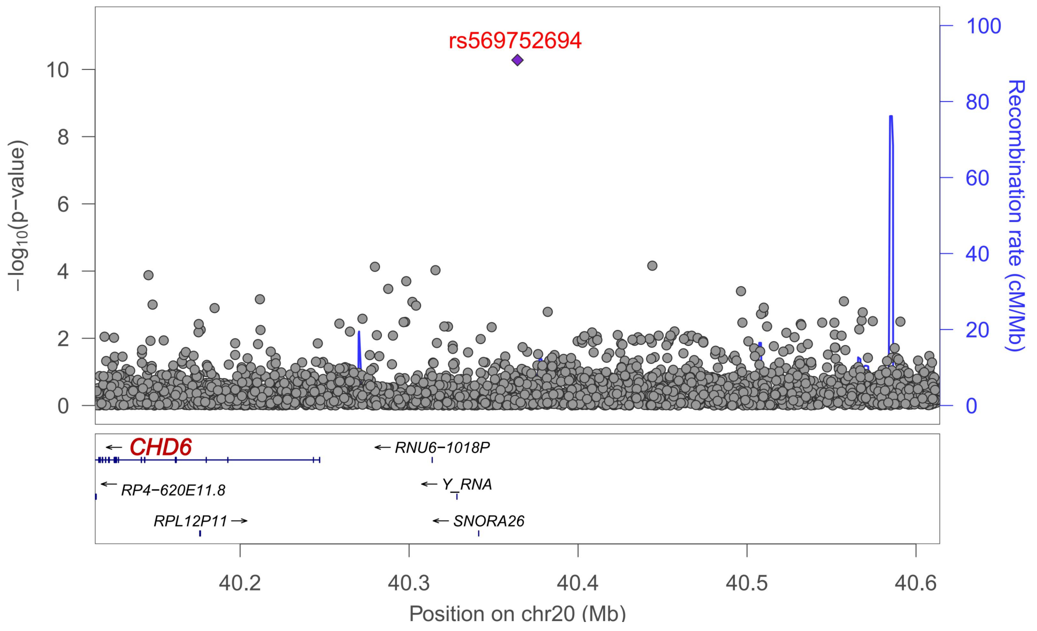

rs569752694
−log10(p−value)
Recombination rate (cM/Mb)
CHD6
RNU6−1018P
RP4−620E11.8
Y_RNA
RPL12P11
SNORA26
40.2
40.3
40.4
40.5
40.6
Position on chr20 (Mb)

**AJ.**

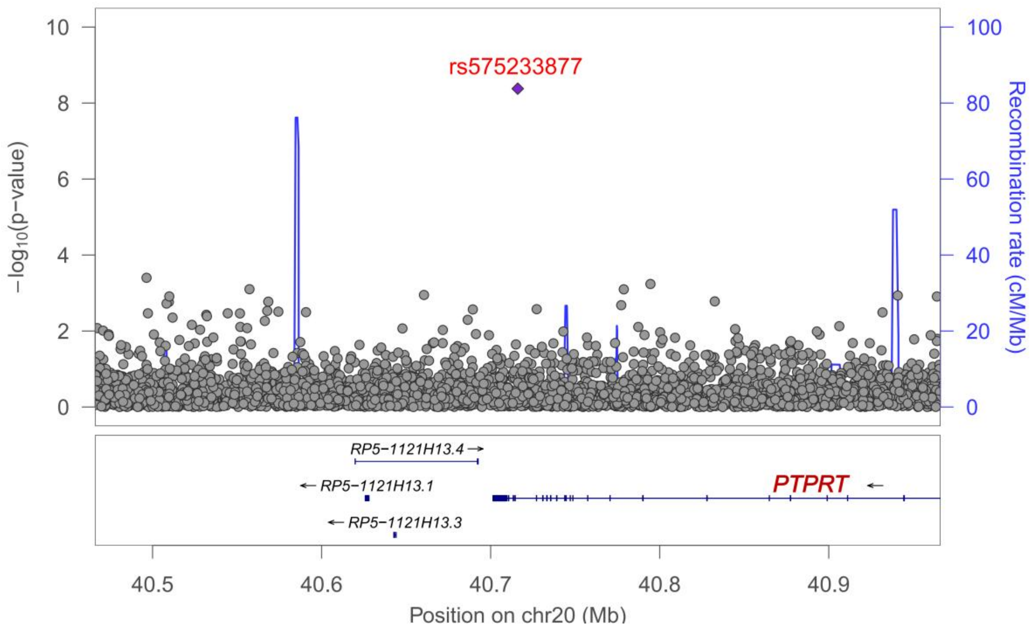

rs575233877
−log10(p−value)
Recombination rate (cM/Mb)
RP5−1121H13.4
RP5−1121H13.1
RP5−1121H13.3
PTPRT
Position on chr20 (Mb)


**AK.**

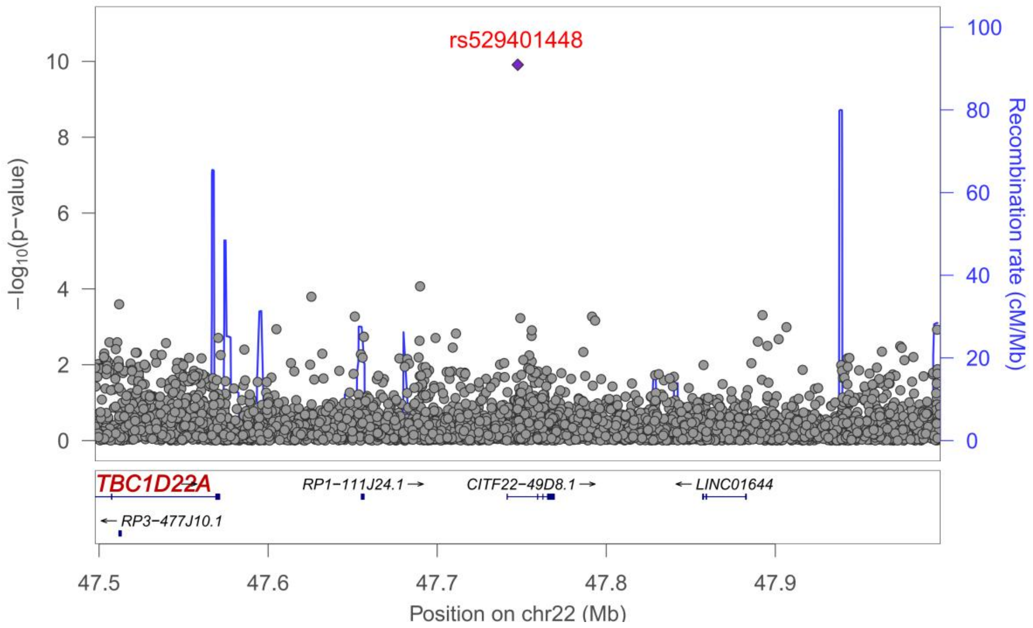

rs529401448
−log10(p−value)
Recombination rate (cM/Mb)
TBC1D22A
RP1−111J24.1
CITF22−49D8.1
LINC01644
RP3−477J10.1
Position on chr22 (Mb)

**Supplementary Figure 4. Box plot of genes from differential expression analysis using GSE282111.**

**A.**

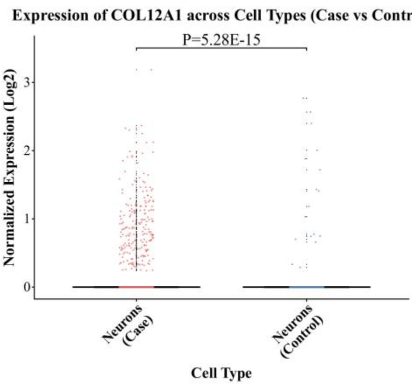


**B.**

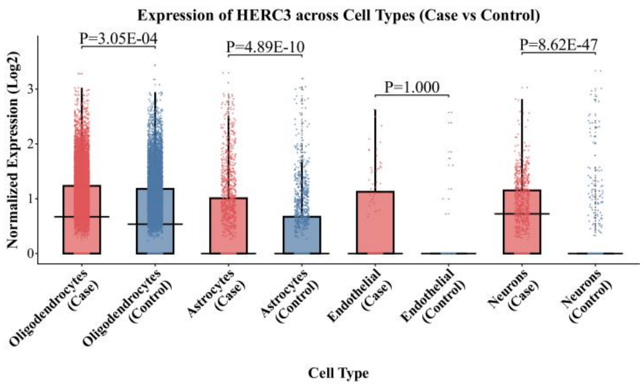


**C.**

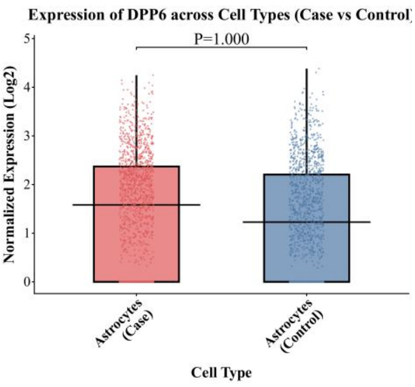


**D.**

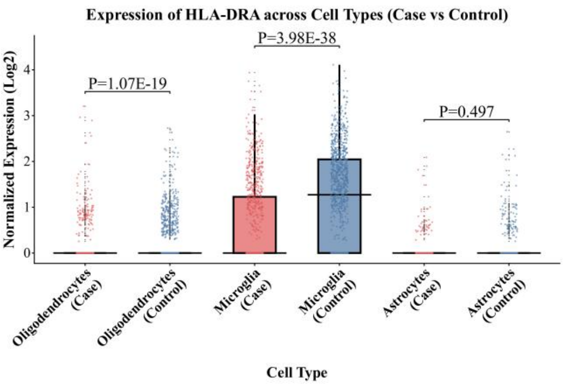


**E.**

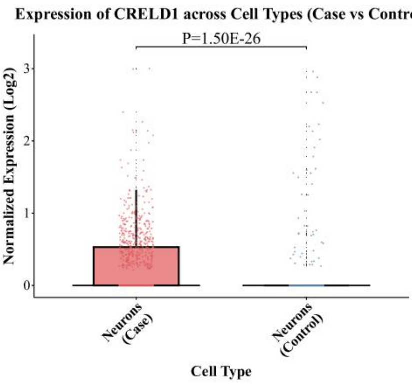


**F.**

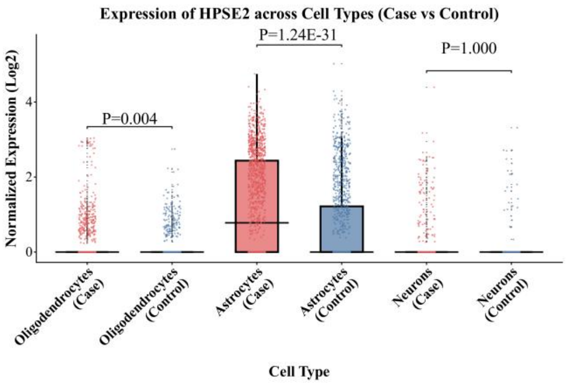


**G.**

**H.**

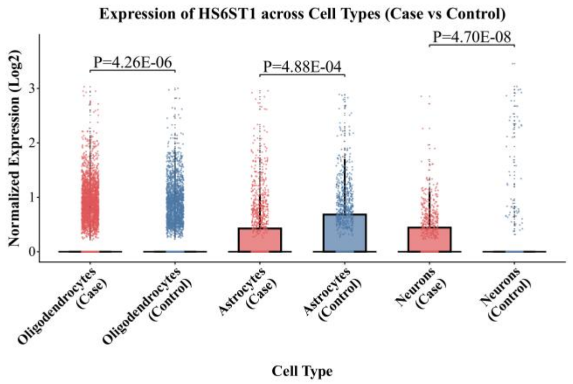
Expression of HS6ST1 across Cell Types (Case vs Control)
P=4.26E-06
P=4.88E-04
P=4.70E-08
Normalized Expression (Log2)
Oligodendrocytes (Case)
Oligodendrocytes (Control)
Astrocytes (Case)
Astrocytes (Control)
Neurons (Case)
Neurons (Control)
Cell Type

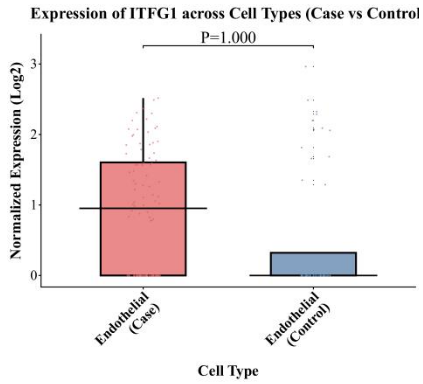
Expression of ITFG1 across Cell Types (Case vs Control
P=1.000
Normalized Expression (Log2)
Endothelial (Case)
Endothelial (Control)
Cell Type

I.

J.

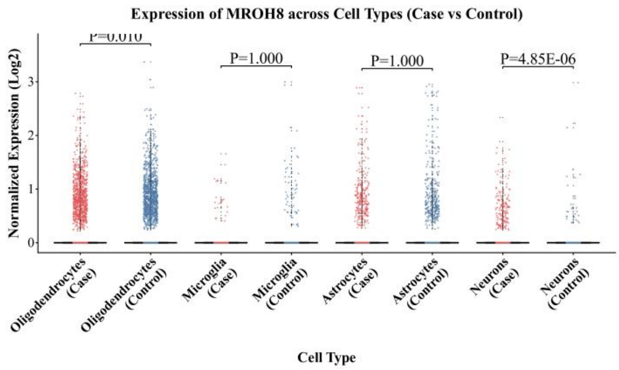
Expression of MROH8 across Cell Types (Case vs Control)
P=0.010
P=1.000
P=1.000
P=4.85E-06
Normalized Expression (Log2)
Oligodendrocytes (Case)
Oligodendrocytes (Control)
Microglia (Case)
Microglia (Control)
Astrocytes (Case)
Astrocytes (Control)
Neurons (Case)
Neurons (Control)
Cell Type

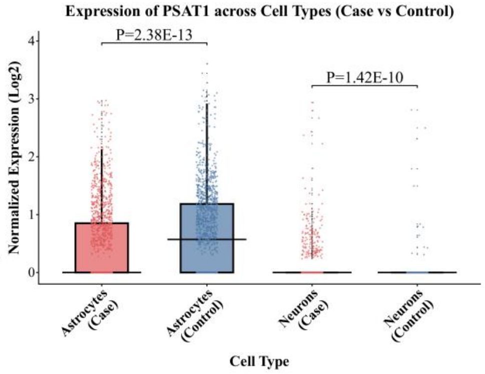
Expression of PSAT1 across Cell Types (Case vs Control)
P=2.38E-13
P=1.42E-10
Normalized Expression (Log2)
Astrocytes (Case)
Astrocytes (Control)
Neurons (Case)
Neurons (Control)
Cell Type

K.

L.

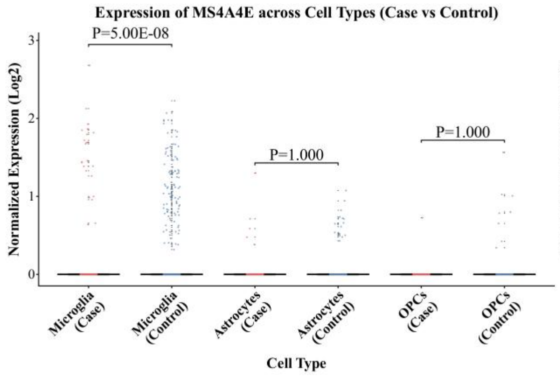
Expression of MS4A4E across Cell Types (Case vs Control)
P=5.00E-08
P=1.000
P=1.000
Normalized Expression (Log2)
Microglia (Case)
Microglia (Control)
Astrocytes (Case)
Astrocytes (Control)
OPCs (Case)
OPCs (Control)
Cell Type

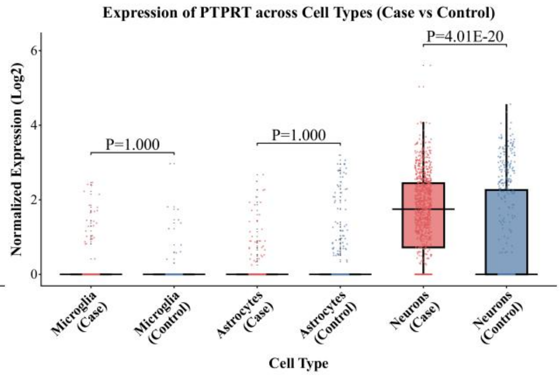
Expression of PTPRT across Cell Types (Case vs Control)
P=1.000
P=1.000
P=4.01E-20
Normalized Expression (Log2)
Microglia (Case)
Microglia (Control)
Astrocytes (Case)
Astrocytes (Control)
Neurons (Case)
Neurons (Control)
Cell Type

L.

N.

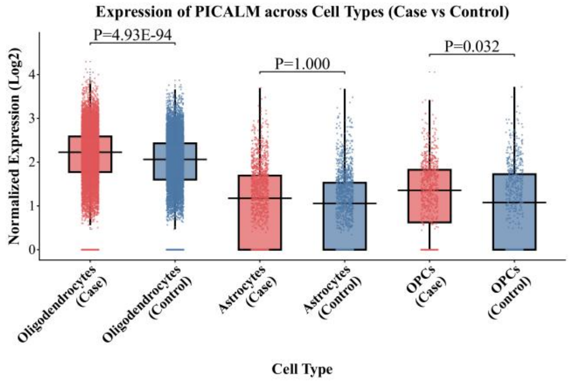
Expression of PICALM across Cell Types (Case vs Control)
P=4.93E-94
P=1.000
P=0.032
Normalized Expression (Log2)
Oligodendrocytes (Case)
Oligodendrocytes (Control)
Astrocytes (Case)
Astrocytes (Control)
OPCs (Case)
OPCs (Control)
Cell Type

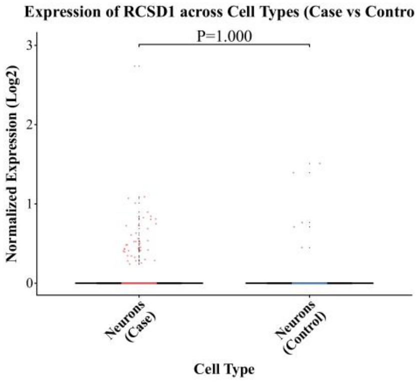
Expression of RCSD1 across Cell Types (Case vs Contro
P=1.000
Normalized Expression (Log2)
Neurons (Case)
Neurons (Control)
Cell Type

O.

P.

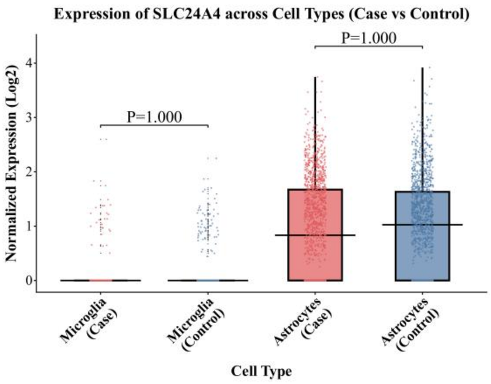

Expression of SLC24A4 across Cell Types (Case vs Control)
P=1.000
P=1.000
Normalized Expression (Log2)
0
1
2
3
4
Microglia (Case)
Microglia (Control)
Astrocytes (Case)
Astrocytes (Control)
Cell Type


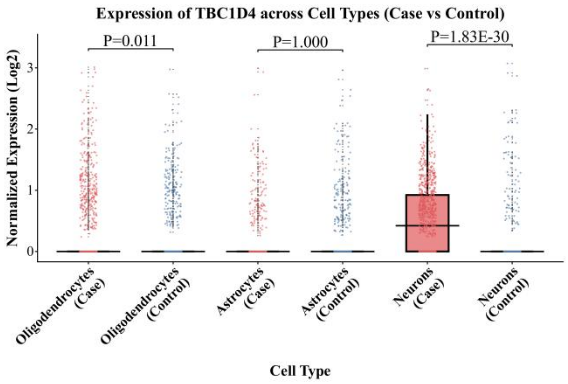

Expression of TBC1D4 across Cell Types (Case vs Control)
P=0.011
P=1.000
P=1.83E-30
Normalized Expression (Log2)
0
1
2
3
Oligodendrocytes (Case)
Oligodendrocytes (Control)
Astrocytes (Case)
Astrocytes (Control)
Neurons (Case)
Neurons (Control)
Cell Type


**P.**

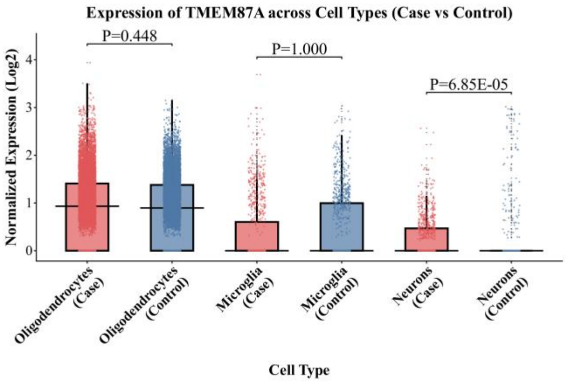

Expression of TMEM87A across Cell Types (Case vs Control)
P=0.448
P=1.000
P=6.85E-05
Normalized Expression (Log2)
0
1
2
3
4
Oligodendrocytes (Case)
Oligodendrocytes (Control)
Microglia (Case)
Microglia (Control)
Neurons (Case)
Neurons (Control)
Cell Type


**R.**

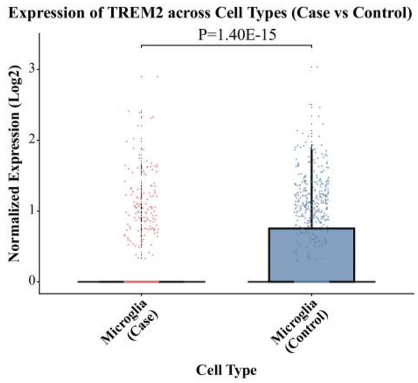

Expression of TREM2 across Cell Types (Case vs Control)
P=1.40E-15
Normalized Expression (Log2)
0
1
2
3
Microglia (Case)
Microglia (Control)
Cell Type


**S.**

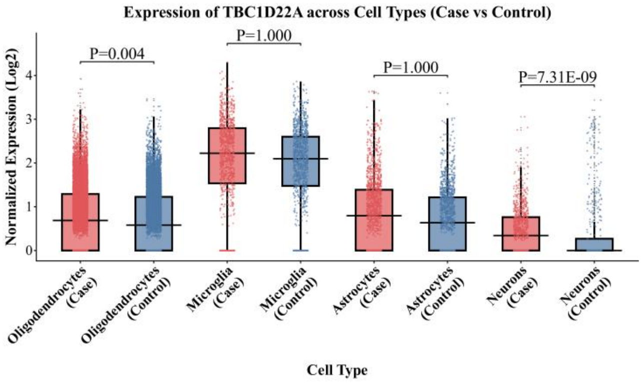

Expression of TBC1D22A across Cell Types (Case vs Control)
P=0.004
P=1.000
P=1.000
P=7.31E-09
Normalized Expression (Log2)
0
1
2
3
4
Oligodendrocytes (Case)
Oligodendrocytes (Control)
Microglia (Case)
Microglia (Control)
Astrocytes (Case)
Astrocytes (Control)
Neurons (Case)
Neurons (Control)
Cell Type
