## Supplemental data1 will be used for the link to the file on the preprint site. for "Genome-wide cross-trait analysis of vascular dementia and Alzheimer’s disease highlights novel loci and lung-brain axis"

### Supplementary Data 1

**Circos plots of chromatin interactions and eQTLs for each chromosome harboring VD risk loci based on the European-specific meta-analysis (Stage 1).** Outer-most layer: Manhattan plot displaying lead SNPs, where additional independent significant SNPs in loci are colored according to linkage disequilibrium ($r^2$; red [$r^2$>0.8], orange [$r^2$>0.6], green [$r^2$>0.4], blue [$r^2$>0.2], grey [$r^2$≤0.2]); $-\log_{10}$ $P$-values for meta-analysis effect estimates for each SNP is indicated on the y-axis. Second layer: chromosome ring with coordinates and genomic risk loci highlighted in blue. Third layer: chromosome ring showing genes mapped by chromatin interactions (orange), eQTL (green), or both (red).

#### Chromosome 1

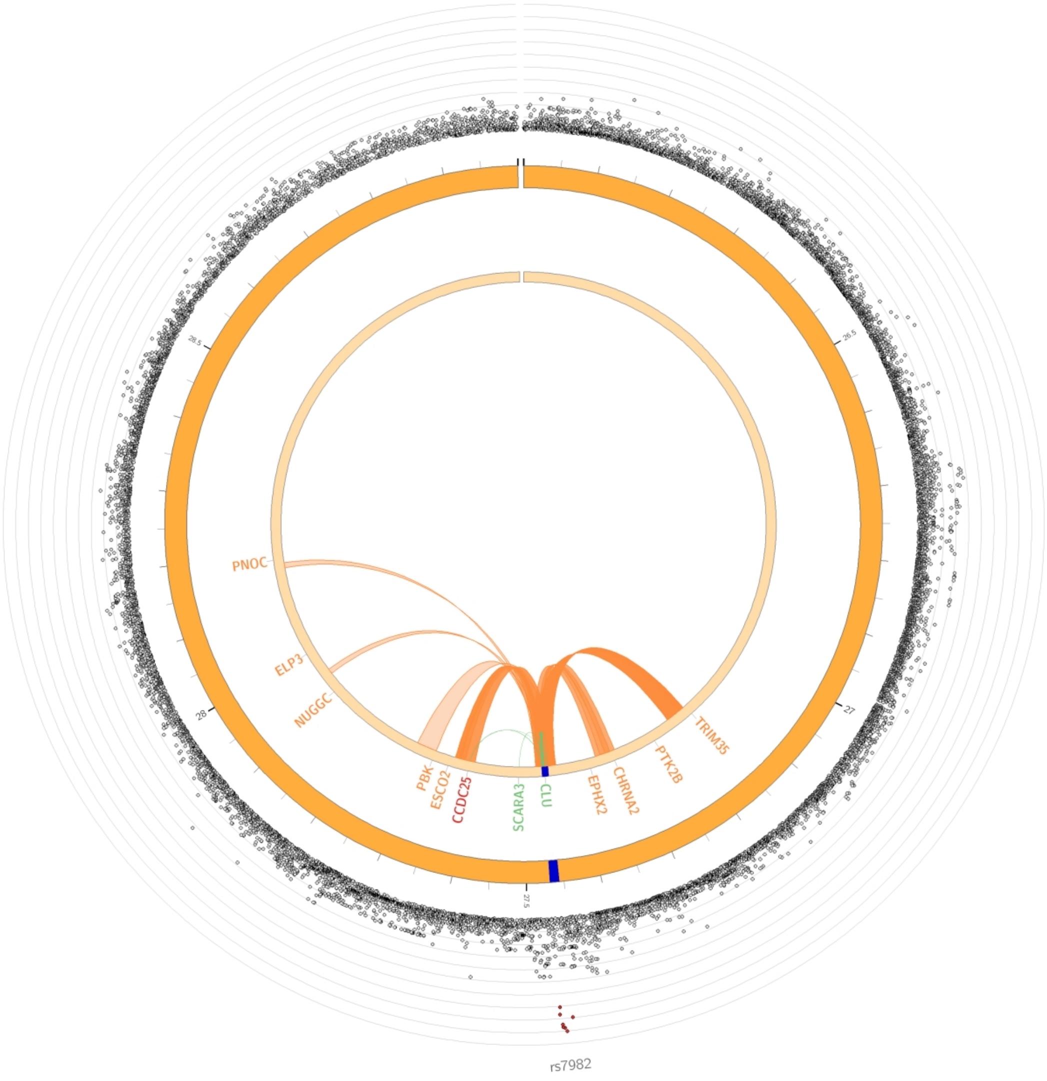

### Chromosome 2

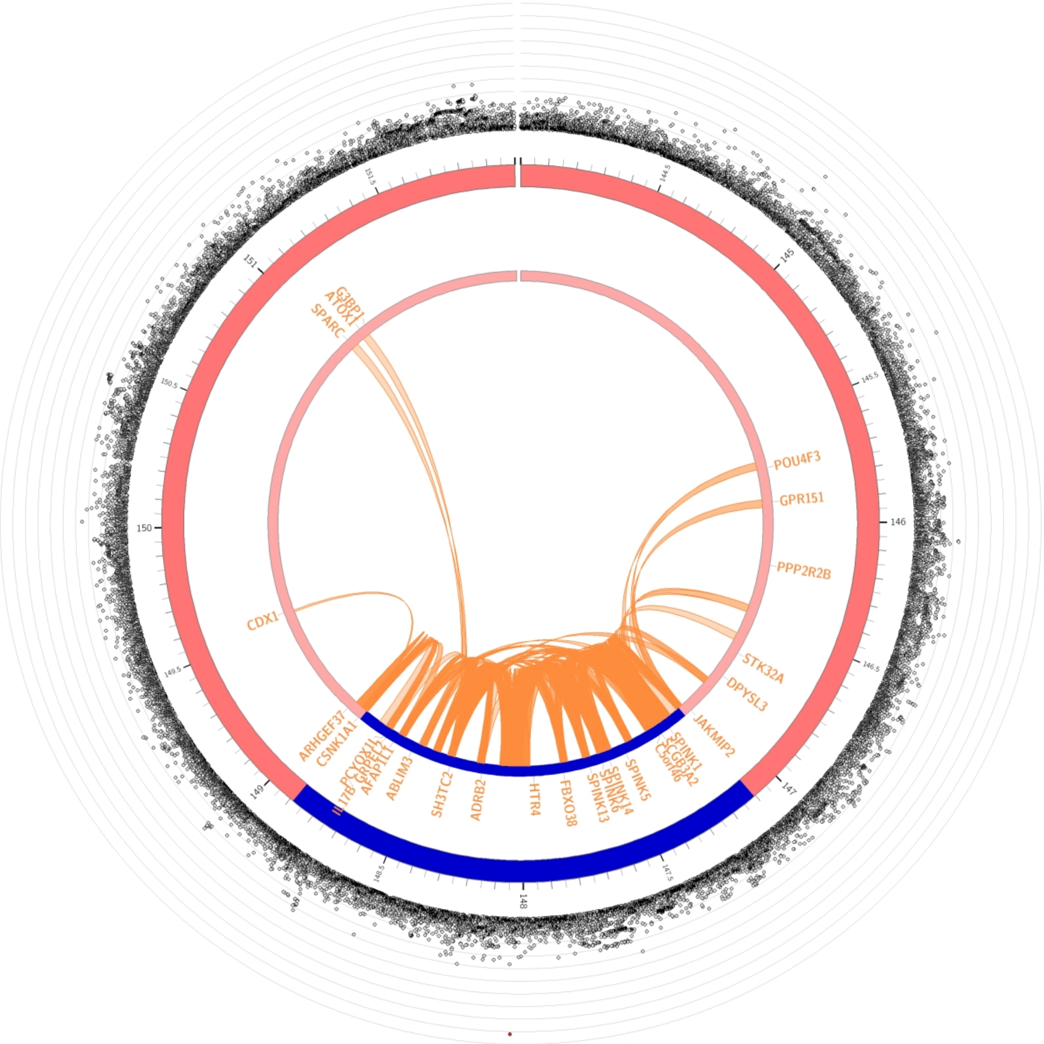

#### Chromosome 3

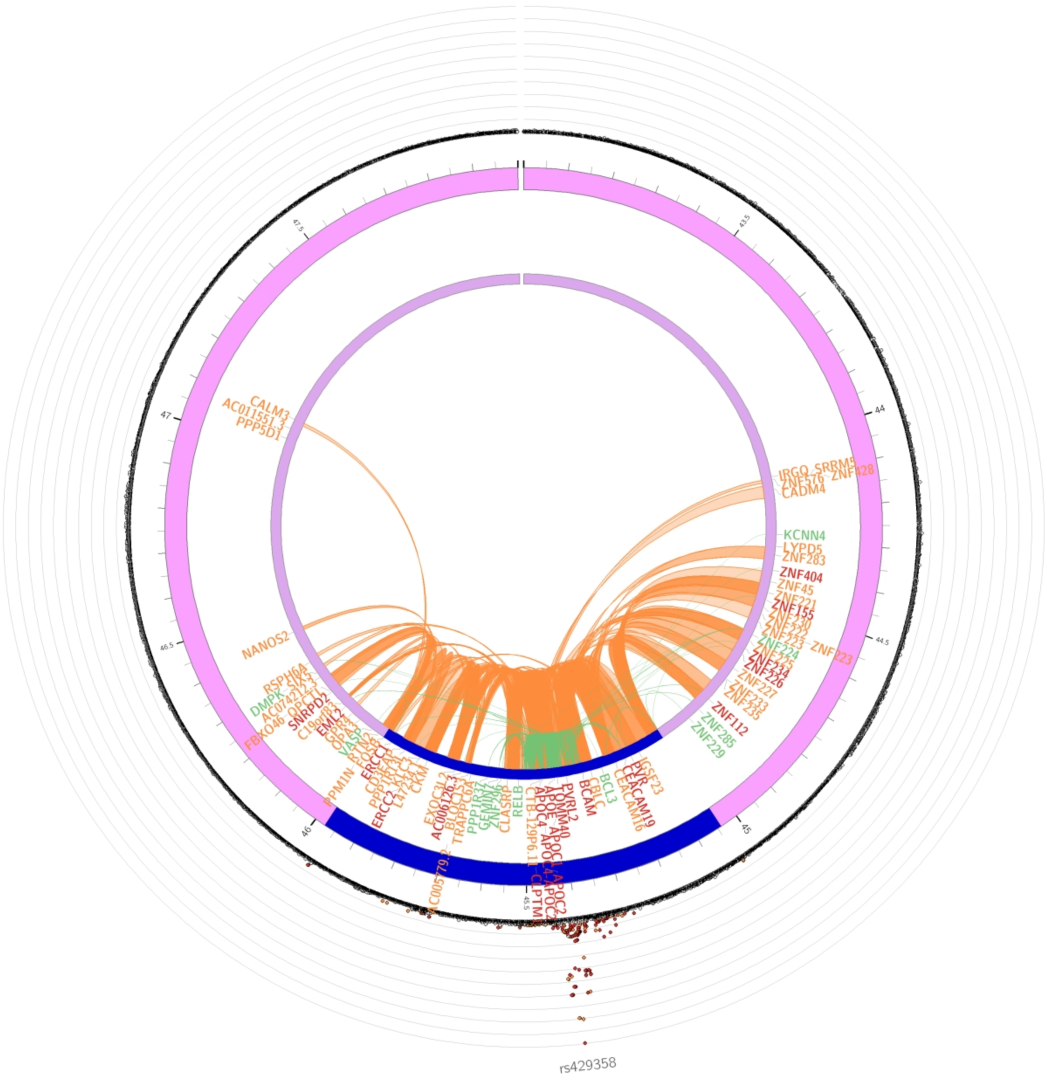
